# A Mechanistic Framework for Modeling Social Gradients in Emerging Infectious Disease Mortality: Evidence from Brazil

**DOI:** 10.1101/2025.04.23.25326307

**Authors:** Jordan D. Klein

**Affiliations:** Max Planck Institute for Demographic Research, Konrad-Zuse-Straße 1, 18057 Rostock, Germany

**Keywords:** COVID-19, Health inequalities, Social epidemiology, Infectious disease modeling, Mechanistic modeling, Fundamental Cause Theory

## Abstract

According to Fundamental Cause Theory, social inequalities in mortality persist even as disease burdens shift. Emerging infectious diseases however, present a particular challenge for studying these inequalities, as protective interventions are introduced while epidemics unfold, making it difficult to disentangle the effects of pre-existing social disadvantage from unequal access to interventions.

To address this challenge, I develop a theory-guided mechanistic modeling framework that embeds pathways to mortality inequalities within a geospatial epidemic model. I apply this framework to the COVID-19 pandemic in Brazil using a susceptible–exposed–infectious–recovered–deceased model. The model incorporates pre-existing social inequalities through disparities in household transmission and infection fatality rates. I simulate non-pharmaceutical interventions (NPIs) and vaccination under nine counterfactual distribution scenarios ranging from no interventions to observed real-world allocation and alternative more equitable strategies.

In the absence of interventions, mortality becomes increasingly concentrated in the most vulnerable municipalities as the epidemic spreads. NPIs adopted following observed, socially stratified patterns accelerate rather than create this gradient, suggesting that pre-existing structural inequalities were the primary drivers of Brazil’s COVID-19 mortality gradient, while unequal intervention uptake reinforced these dynamics.

However, prioritizing the most vulnerable municipalities for vaccination reverses the gradient, an effect further compounded by equal adoption of NPIs, neutralizing the impact of these pre-existing conditions. These findings can help guide more equitable policy responses to future emerging infectious disease crises. This study is, to our knowledge, the first mechanistic model to operationalize Bambra’s pathways framework of inequalities in emerging infectious diseases.

**Highlights:**

- Fundamental cause theory applied to mechanistic epidemiological modeling
- Pre-existing social inequities can drive epidemic mortality disparities
- Intervention distribution can reinforce or reverse mortality inequalities
- National geospatial epidemic model illustrates COVID-19 inequality dynamics in Brazil
- First model to operationalize Bambra’s inequality pathways framework

## 1. Introduction

The fundamental cause theory (FCT) asserts that social conditions are the fundamental causes of disease (Link & Phelan, 1995). Relative advantages and disadvantages, by which those with greater resources can leverage them to gain access to protective factors, avoid risk factors, or receive effective treatments for disease relative to those with fewer resources, are transferable across epidemiological paradigms, including the emergence of novel infectious diseases (EIDs). Thus, while the proximal determinants of disease can change as societies go through health and epidemiological transitions, inequalities in health persist (Donkin, 2014; Phelan et al., 2004). Such inequalities, where populations with higher socioeconomic status (SES) have better health and lower mortality than those with lower SES, in line with the expectations of FCT, are termed *expected* social gradients in health.

Clouston et al. (2016) attempt to explain these dynamics through their theoretical model of the social history of disease, synthesizing FCT with epidemiological transition theory (ETT) proposed by Omran (2001). According to this theory, in the first stage of a disease’s social history, when there is no knowledge about it, nor are preventions or treatments available, populations with higher SES are not able to effectively leverage their greater access to resources to achieve lower mortality from the disease relative to low SES populations, a phenomenon that Clouston et al. (2016) denote “natural mortality.” Following the advent of preventative and curative countermeasures, their unequal diffusion according to SES is what gives rise to social disparities in mortality. However, this model does not account for how pre-existing inequities in access to resources may result in inequities in vulnerability to disease in general (Farmer, 1996), even prior to the advent of such protective interventions.

To address this theoretical gap, I introduce a model that builds upon the pioneering work of Bambra (2022) elucidating distinct pathways through which EID inequities arise, encompassing both pre-existing disease-agnostic inequities and the unequal adoption of disease-specific interventions. According to this model, which can be illustrated using the recent global pandemic of COVID-19, social gradients in mortality can be produced through four pathways, either stemming from disease-agnostic pre-existing inequities or disease-specific interventions which are adopted unequally (Fig. 1). Within these two categories, inequities in mortality can be produced through inequities in exposure or inequities in outcomes. Within disease-agnostic pre-existing inequities, inequities in exposure are encapsulated by the unequal forces of infection that result from disparities in living conditions, while inequities in outcomes are encapsulated by disparities in disease lethality stemming from inequities in health status and healthcare access and quality. Meanwhile, disparities in COVID-specific non-pharmaceutical intervention (NPI) uptake result in inequities in risk of exposure, while disparities in the uptake of COVID-specific pharmaceutical interventions yield inequities in outcomes if exposed to the disease.

**Fig. 1.**
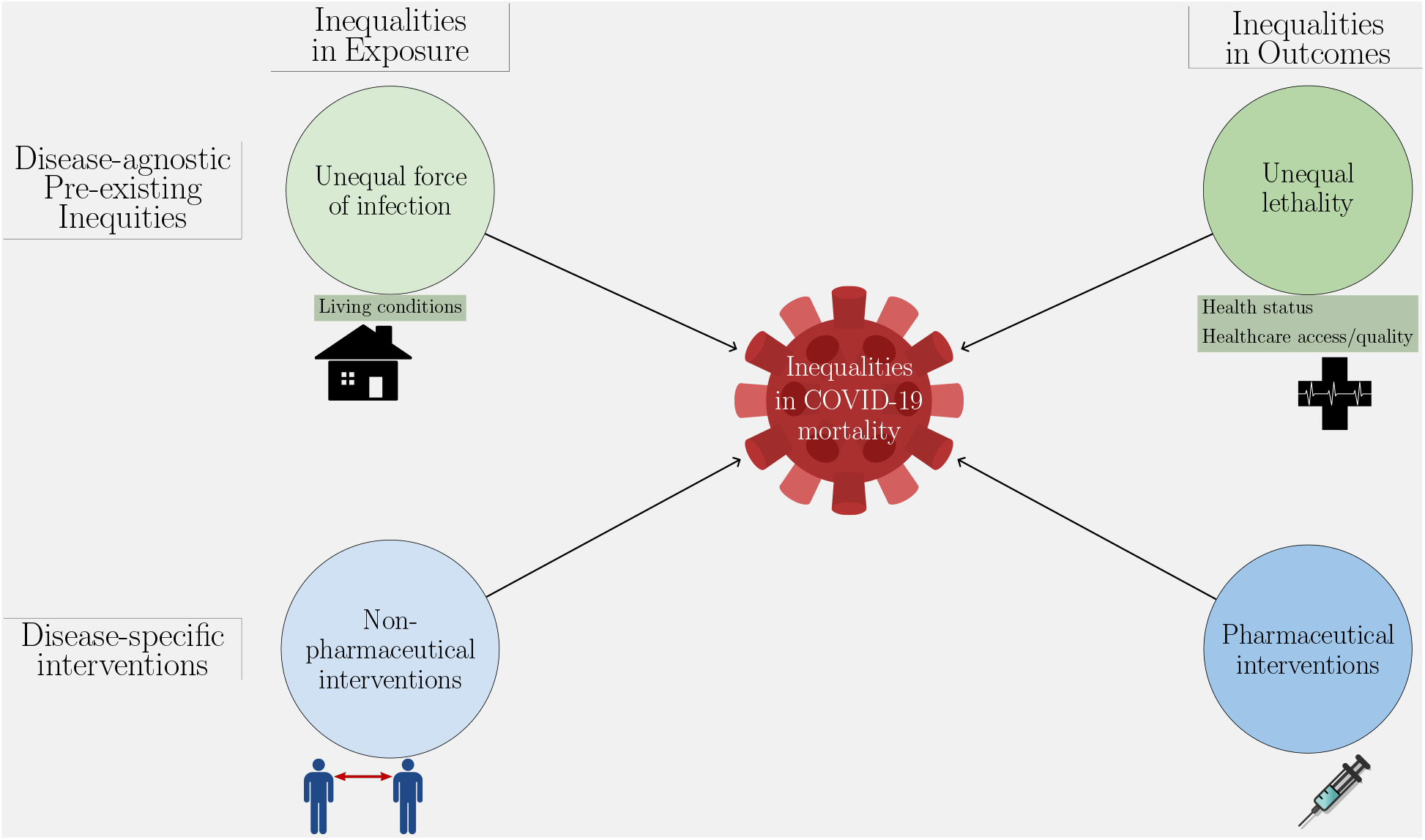
Proposed pathways to COVID-19 mortality gradients (inspired by (Bambra, 2022)).

To illustrate these dynamics empirically, I examine the case of COVID-19 in Brazil, a country characterized by high levels of social and economic inequality, with the 17th highest income inequality in the world according to the World Bank. Brazil has been among the countries most severely impacted by COVID-19, with the 10th highest officially reported mortality rate in the world during 2020 and the first half of 2021, and the highest among countries with a population of 40 million or more (Ritchie et al., 2020).

However, the literature thus far on the temporal dynamics of social gradients in COVID-19 mortality in Brazil has been limited, especially compared to countries in the Global North (Beese et al., 2022; Rhodes et al., 2024), though the evidence does show that social gradients in mortality developed in the months following COVID-19’s emergence, with deaths increasingly concentrating in disadvantaged geographic areas and populations (Andrade et al., 2022; Bermudi et al., 2021; Brizzi et al., 2022; Cajazeiro et al., 2024; Castilho et al., 2023; Chauvin, 2024; Collazos et al., 2023; Raymundo et al., 2023; Razafindrakoto et al., 2022; Santana et al., 2024; Tavares & Betti, 2021). Explaining how these social gradients were produced is complicated by their contemporaneous emergence with the initial roll-out of NPIs, implemented early in the pandemic in Brazil beginning in mid-to-late March 2020 (Faria de Moura Villela et al., 2021; Li et al., 2021), thus making it challenging to disentangle the contributions of disease-specific interventions from disease-agnostic pre-existing inequities.

Multiple pathways likely contribute to producing social gradients in epidemic mortality. Previous work has found higher prevalence of comorbidities and poorer pre-existing health status in lower SES populations, which have been associated with higher rates of mortality (Li et al., 2021; Liu et al., 2022; Nassif Pires et al., 2020; Nunes et al., 2020; Razafindrakoto et al., 2022; Rocha et al., 2021). Other factors include inequities in healthcare access and quality. In the Brazilian case, these inequities are reflected in greater utilization of the public as opposed to the private health system, greater distance to the nearest healthcare facility, and fewer hospital beds, intensive care units (ICUs), and physicians per capita in lower SES geographies, all of which increase the risk of mortality in COVID-19 patients through impeding access to quality healthcare (Boing et al., 2022; Brizzi et al., 2022; Li et al., 2021; Nassif Pires et al., 2020; Razafindrakoto et al., 2022; Rocha et al., 2021; Tavares & Betti, 2021).

Pre-existing disparities in living conditions may increase the risk of exposure to infectious disease in more deprived communities (Bambra, 2022). Evidence from Brazil shows that COVID-19 incidence and mortality rates were higher in lower SES geographies with poorer quality, less sanitary, more crowded, and especially denser housing and living conditions (Chauvin, 2024; Nassif Pires et al., 2020; Nunes et al., 2020; Razafindrakoto et al., 2022; Rocha et al., 2021; Tavares & Betti, 2021).

Once NPIs are introduced during an epidemic, inequities in their adoption may produce social gradients in mortality through disparities in exposure (Bambra, 2022). Frontline, service sector, informal, and other workers in occupations that require in-person contact and/or that do not have paid leave are disproportionately concentrated in lower SES geographies in the Brazilian context (De Negri et al., 2021; Li et al., 2021; Nassif Pires et al., 2020; Nunes et al., 2020; Razafindrakoto et al., 2022; Rocha et al., 2021). Consequently, while mobility sharply fell in the country as a whole following the introduction of social distancing NPIs (COVID-19 Mobility Data Network, 2020; Meta, 2023), lower SES states and municipalities with more frontline, service sector, informal, and other lower-skilled, lower-wage workers reported lower compliance with these measures and saw smaller mobility reductions (Li et al., 2021; Rocha et al., 2021; Tavares & Betti, 2021).

Unequal uptake of pharmaceutical interventions, especially vaccines once they become available, can further contribute to social gradients in mortality. Although vulnerable populations were prioritized in the Brazilian vaccination campaign, lower SES geographies had lower vaccine coverage (Martins et al., 2023; Villar et al., 2024), especially after controlling for political behavior (votes for Jair Bolsonaro in the 2018 and 2022 presidential elections) (Razafindrakoto et al., 2024; Seara-Morais et al., 2023), as a result of inadequate logistics and health systems capacity.

To explore how these pathways generate social gradients in epidemic mortality, I operationalize them in a data-driven geospatially and socially stratified compartmental model. I apply it to the case of COVID-19 in Brazil, designating population classes for each of its 5,565 municipalities. Drawing from real-world data, I parameterize each pathway as depicted in Figure 1. Pre-existing disparities in living conditions across municipalities, represented by household size, are used to capture the unequal force of infection pathway. For the unequal lethality pathway, I estimate infection fatality rates (IFR) before vaccines and treatments became widely available to represent pre-existing disparities in health status and variations in healthcare access and quality. The model incorporates inequities in NPIs by evaluating the proportional change in travel outside of home areas post-introduction of social distancing measures, relative to pre-pandemic patterns. Lastly, disparities in vaccination rates across municipalities are integrated to represent inequities in pharmaceutical interventions.

This comprehensive approach allows me to address a key challenge in studying inequalities during emerging infectious disease epidemics: empirical evidence often does not allow us to isolate the impacts of pre-existing inequities from those of disease-specific interventions on mortality gradients. In essence, we lack concrete evidence to understand how epidemics might unfold if only one of these driving forces of inequity were at play. Models using this framework allow us to gain insights into hypothetical counterfactual scenarios that empirical methods cannot. We can thus examine how social gradients in mortality could develop if no NPIs were introduced, if a vaccine were available at the beginning of an epidemic, and/or if these interventions were distributed more equitably than observed in real-world settings. In this study I construct a model world to explore how gradients in emerging infectious disease mortality would evolve under counterfactual scenarios using data-driven parameters; the goal is not necessarily to fit model dynamics to observed epidemic trajectories.

Some studies have systematically incorporated social vulnerability into mathematical models of infectious disease (Hiraoka et al., 2022; Sajjadi et al., 2025; Tizzoni et al., 2022), but important gaps remain, particularly in Global South contexts such as Brazil (Naidoo et al., 2024). The limited number of studies that have incorporated social vulnerability into mathematical models of COVID-19 in Brazil have focused on cities or metropolitan areas rather than the national level and have typically relied on hypothetical mobility and contact networks rather than real-world data (Barreto et al., 2024; Brotherhood et al., 2022; Cardoso et al., 2020). To our knowledge, this is the first modeling study to explicitly operationalize Bambra (2022)’s pathways framework linking inequality and EIDs. Empirically, it applies this framework through the first national-scale geospatial epidemic model of COVID-19 in Brazil based on real-world mobility network data. This study contributes a mechanistic modeling framework for decomposing structural drivers of mortality inequality during epidemic emergence. By parameterizing key pathways identified in social epidemiology, including exposure, susceptibility, and intervention uptake, we demonstrate how structural inequality can generate dynamic mortality gradients under different policy scenarios.

I posit that unequal adoption of NPIs and vaccination could entrench the effects of pre-existing inequities in the production of social gradients in epidemic mortality. I further hypothesize that more equitable distribution of these interventions, prioritizing the protection of lower SES populations, could counteract or even reverse the impacts of these pre-existing inequities.

## 2. Materials and Methods

### 2.1. The model

I construct a compartmental model governed by the following set of equations.

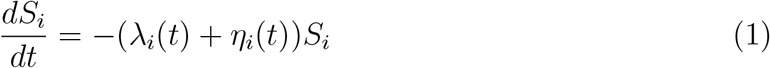

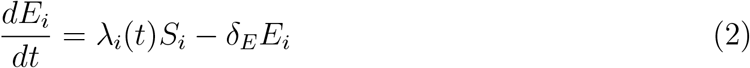

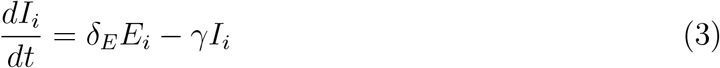

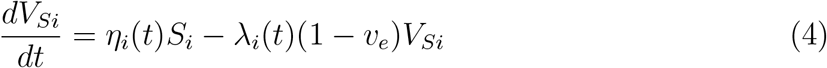

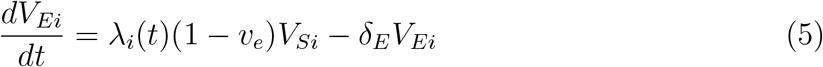

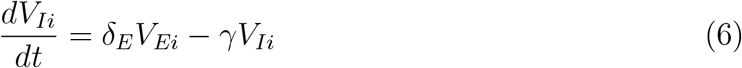

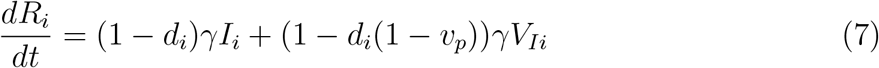

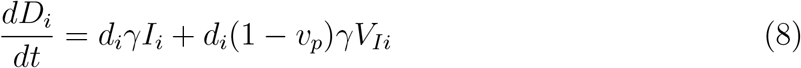

There are eight compartments for a given municipality *i*: unvaccinated susceptible (*S*), exposed (*E*), and infected (*I*); vaccinated susceptible (*V*_*S*_), exposed (*V*_*E*_), and infected (*V*_*I*_); plus recovered (*R*) and deceased (*D*) (Table 1). *λ*_*i*_(*t*) is the force of infection (FOI) (Table 2), which can vary over time *t*. Individuals progress from exposed to infected at rate *δ*_*E*_ and recover from infection at rate *γ* through either full recovery or death, with an IFR of *d*_*i*_. Susceptible individuals can also be vaccinated at a time-varying rate *η*_*i*_(*t*). Vaccine efficacy against infection is *v*_*e*_ and against death post-infection is *v*_*p*_ (see Derivation of vaccination parameters, supplementary materials, for details). Since I am only modeling the first six months after disease introduction, I do not consider loss of immunity from either natural infection nor vaccination.

**Table 1.** Model compartments.

| Compartment | Description |
| --- | --- |
| $S$ | Susceptible (*unvaccinated*) |
| $E$ | Exposed (*unvaccinated*) |
| $I$ | Infected (*unvaccinated*) |
| $V_S$ | Susceptible (*vaccinated*) |
| $V_E$ | Exposed (*vaccinated*) |
| $V_I$ | Infected (*vaccinated*) |
| $R$ | Recovered |
| $D$ | Deceased |

**Table 2.** Model parameters.

| Parameter | Description | Value | Source |
| --- | --- | --- | --- |
| $\lambda_i(t)$ | Force of infection (FOI) | *Municipality-specific* | *Derived* |
| $\delta_E^{-1}$ | Latent period (days) | 2.9 | Pascual-García et al. |
| $\gamma^{-1}$ | Infectious period (days) | 7 | Pascual-García et al. |
| $d_i$ | Infection fatality rate | *Municipality-specific* | Ministério da Saúde<br>Brasil.IO |
| $\eta_i(t)$ | Rate of vaccine<br>administration (per day) | *Municipality-specific* | Cota |
| $v_e$ | Vaccine efficacy | .89 | ACIP |
| $v_p$ | Vaccine protection<br>against death | .509 | ACIP<br>dos Santos et al. |
| $\lambda_i^{HH}$ | Household/<br>mobility-independent FOI | *Municipality-specific* | *Derived* |
| $\lambda_i^M(t)$ | Extra-household/<br>mobility-dependent FOI | *Municipality-specific* | *Derived* |
| $\beta_i^{HH}$ | Household rate of transmission | *Municipality-specific* | *Derived* |
| $\tau_{HH}$ | Daily household<br>probability of transmission | — | *Derived* |
| $c_i^{HH}$ | Household contact rate | *Municipality-specific* | *Derived* |
| $SAR$ | Household secondary<br>attack rate | .189 (.14, .248) | Madewell et al.<br>Thompson et al. |
| $\beta_{ij}^M(t)$ | Inter/intra-municipality<br>mobility-dependent<br>transmission rate | *Municipality-specific* | *Derived* |
| $\tau_M$ | Mobility-dependent<br>probability of transmission | — | *Derived* |
| $c_i^M(t)$ | Mobility-dependent<br>contact rate | *Municipality-specific* | *Derived* |
| $\omega_{ij}(t)$ | Probability of contact with $j$ | *Municipality-specific* | P. Peixoto, Incognia |
| $HH\bar{size}_i$ | Mean household size<br>(residents/household) | *Municipality-specific* | IBGE |
| $R_0$ | Basic reproduction number | 3.1 (2.4, 5.5) | de Souza et al. |
| $R_0^{HH}$ | $R_0$ of household transmission | — | *Derived* |
| $R_0^M$ | $R_0$ of mobility-dependent transmission | — | *Derived* |
| $\kappa_M$ | Intrinsic mobility-dependent<br>transmission rate | — | *Derived* |
| $\Delta M_i(t)$ | Proportional change<br>in mobility | *Municipality-specific* | COVID-19 Mobility<br>Data Network, Meta |

Notes:
*Derived* parameters indicate that they have been calculated by the author using other parameters with known values.

### 2.2. Force of infection

I break down the FOI of COVID-19 into two components: household, or mobility-independent transmission 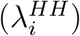, which is time-invariant, and extra-household, or mobility-dependent transmission 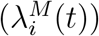, which is time-variant.

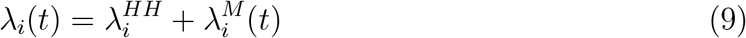

I therefore assume that all mobility-independent transmission occurs within households and hereafter refer to mobility-independent transmission and household transmission interchangeably. I parameterize household transmission with the following equation:

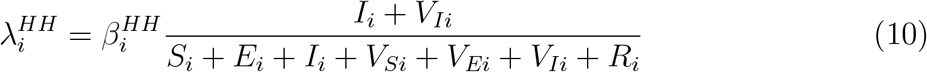

Where 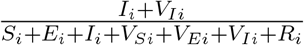 is the proportion of the population in municipality *i* that is infectious. 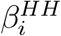 is the rate at which infectious individuals transmit COVID-19 within the household, which can be expressed as:

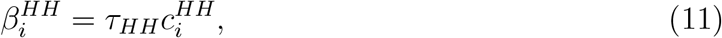

where *τ*_*HH*_ is the daily household-specific probability of transmission given contact and 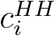 is the number of potentially-infectious household contacts per day. I derive *τ*_*HH*_ from COVID-19 household secondary attack rates (SAR) from meta-analyses by Thompson et al. (2021) (0.166; 95% CI, 0.14-0.193) and Madewell et al. (2020) (0.211; 95% CI, 0.174-0.248). Since household SAR refers to the probability of transmission during the entire duration of the infectious period, and infection events are mutually exclusive, if we assume a uniform probability of infection during each day of the infectious period, *τ*_*HH*_ can be expressed with the following equation:

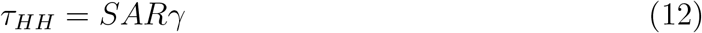

I use the median of the household SAR point estimates obtained by Madewell et al. (2020) and Thompson et al. (2021) and conduct sensitivity analyses using the minimum and maximum values of the 95% confidence intervals of these estimates.

Assuming all transmission of COVID-19 outside of the household is related to mobility, I parameterize this FOI with the following equation:

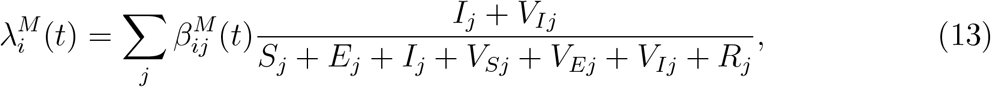

where 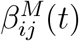 is the mobility-dependent transmission rate between municipalities *i* and *j*, which can be expressed with the following equation:

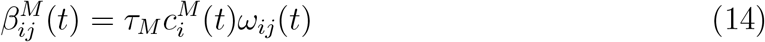

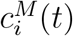 is the number of daily contacts that individuals who live in municipality *i* have outside the home, which can change over time *t*, and *τ*_*M*_ is the probability of infection from these contacts. *ω*_*ij*_(*t*) is the probability that extra-household contacts 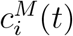 are with individuals from municipality *j*. I thus can re-express the FOI equations of household and mobility-related transmission respectively as the following:

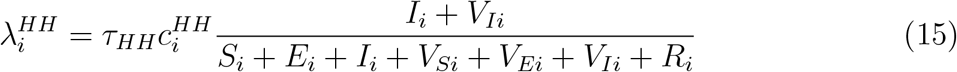

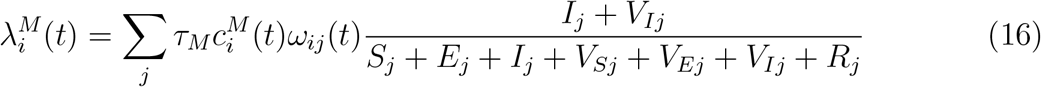

### 2.3. Parameterization of pathways to inequality

I incorporate each of the four pathways to inequality in COVID-19 mortality (Fig. 1) into my model through specific parameters. I incorporate unequal FOIs from pre-existing inequities in living conditions in the parameterization of household FOI (equation 15). Assuming that each individual has on average one potentially infectious contact with each other member of their household per day:

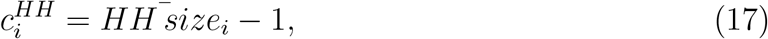

where 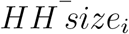 is the mean household size, or number of residents per household, in municipality *i*. This allows equation 15 to be re-expressed as the following.

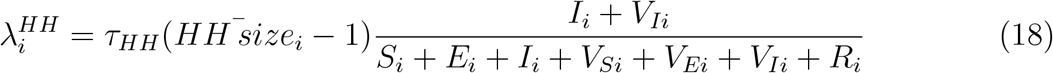

Unequal lethality is captured by the IFR parameter (*d*_*i*_). Since the true number of cases and deaths are latent, I estimate IFR using reported numbers of cases and deaths. I divide cumulative confirmed and suspected COVID-19 deaths for 2020, i.e. deaths through the epidemiological week of 12/27/2020 (the final epidemiological week of 2020), by cumulative confirmed cases of COVID-19 through the epidemiological week of 12/20/2020, one week prior. The IFR estimate for each municipality is therefore the CFR of COVID-19 in 2020 with a one-week lag in cases, since this is the average duration from infection onset to recovery or death. Although I am only modeling the first six months after disease introduction, I estimate IFR using cases and deaths from all of 2020 to minimize the number of municipalities with either missing data or only a very small number of cases, the latter of which are prone to having unusually high or low CFRs due to cases and deaths being discrete events. In addition, no COVID-specific treatments or vaccines were approved nor given emergency use authorization in Brazil until 2021 (Geraque, 2021), suggesting that the IFR of COVID-19 was driven by disease-agnostic pre-existing inequities in lethality from disparities in health status and variations in healthcare access and quality throughout the entire year of 2020. I conduct sensitivity analyses by doubling and halving these IFR estimates.

I introduce disparities in uptake of social distancing NPIs to the model through mobility-related FOI (equation 16). To do so, I first need to derive its constituent parameters. The basic reproduction number (*R*_0_), or the expected number of secondary cases from a single infectious individual in a completely susceptible population, of the wild-type of SARS-CoV-2 in Brazil according to de Souza et al. (2020)’s estimates, is the sum of the basic reproduction numbers specific to both modes of transmission parameterized in this model (equations 15 and 16), such that 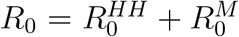 (see Derivation of the basic reproduction number, supplementary materials, for details). I use the point estimate of *R*_0_ obtained by de Souza et al. (2020), and conduct sensitivity analyses using the upper and lower bounds of the 95% credible interval of their estimate.

Given equation 17, 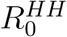 for Brazil as a whole can be expressed as the following (see equation A.16):

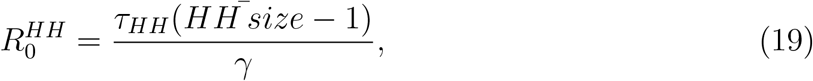

where 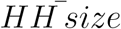 is the mean household size of Brazil as a whole. Assuming frequency-dependent transmission and uniform extra-household contact rates across municipalities prior to the introduction of NPIs (see equation A.17),

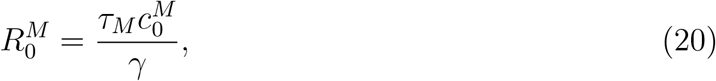

where 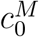 is the null extra-household contact rate at *t* = 0, where no interventions have been implemented. This is justified by previous findings that mobility did not vary by SES in Brazil prior to the introduction of social distancing NPIs (Li et al., 2021; Rocha et al., 2021). Letting the constant 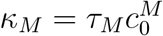, this can be expressed with the following:

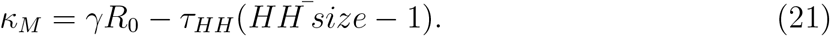

This in turn allows equation 16 to be re-expressed as the following.

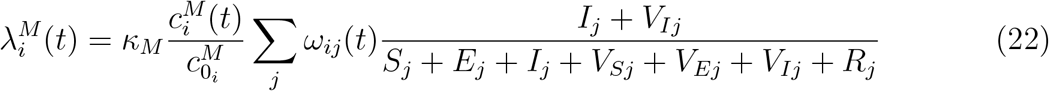

*ω*_*ij*_(*t*) represents the conditional probability that an individual in municipality *i* has contact with individuals from municipality *j*, given that the individual leaves the home. This is estimated using mobility network data provided by P. Peixoto (2019/2022). This data, collected by the company InLoco (now Incognia (2025)), captured the movements of users of mobile applications over distinct 24-hour periods covering over one quarter of the Brazilian population in 2020 (P. S. Peixoto et al., 2020). *i* ≠ *j* thus indicates devices that moved from municipality *i* to municipality *j* within day *t*, while *i* = *j* indicates devices that have moved within their own municipality. Devices that did not move at all were not captured. According to P. S. Peixoto et al. (2020), although this is not necessarily a statistically consistent estimator of the population probabilities of traveling between municipalities, as the same device may be recorded more than once in a day, it provides a good representation of the overall mobility network and its dynamics.

Finally, the uptake of social distancing NPIs is captured in this model by the term 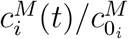 denoting the relative rate of extra-household contacts of residents of municipality *i* at time *t* compared to the null, which can be expressed as:

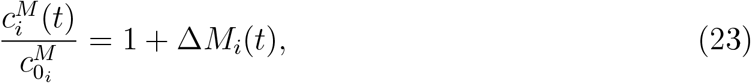

where Δ*M*_*i*_(*t*) is the proportional change in mobility outside the home of individuals living in municipality *i* over time *t* following the introduction of NPIs, compared to a pre-COVID baseline. (In scenarios with no social distancing, 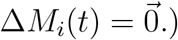 These estimates are obtained from Facebook Data for Good Movement Range Maps, where individuals with an evening location in municipality *i* are considered residents (COVID-19 Mobility Data Network, 2020; Meta, 2023). This parameterization relies on the assumption that changes in overall mobility are proportional to changes in mobility-dependent contacts. For sensitivity analyses, I double and halve the impact of these changes in mobility Δ*M*_*i*_(*t*) on the resultant relative rate of extra-household contacts 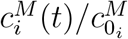. Municipalities without sufficient observations to produce estimates of *ω*_*ij*_(*t*) or Δ*M*_*i*_(*t*) are imputed using the inverse squared distance-weighted means of municipalities without missing data.

Measuring social distancing NPI uptake during the COVID-19 pandemic using changes in mobility from digital trace data is an established practice, including in Brazil (Barreto et al., 2024; Brotherhood et al., 2022; Li et al., 2021; Razafindrakoto et al., 2022; Rocha et al., 2021; Tavares & Betti, 2021). This previous work has found similar mobility levels using these data sources across geographies prior to the introduction of NPIs, succeeded by sharp reductions in mobility following their introduction in mid March. These mobility reductions were also associated with occupational status, with areas having more informal and frontline workers showing smaller reductions than areas with more remote-capable professionals.

Inequities in vaccine uptake are incorporated into the model through the *η*_*i*_(*t*) parameter. (In scenarios with no vaccination 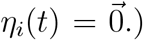 Real-world data on the rate of vaccine administration by municipality is obtained from Cota (2020), which compiles data on the Brazilian COVID-19 vaccination campaign from Ministério da Saúde (2024) beginning on 01/16/2021. For calculating the rate of vaccination uptake, each vaccination event is defined as the time that an individual received their second dose. All vaccines administered are assumed to confer the levels of protection against infection (symptomatic and asymptomatic) and death of BNT162b2 (Pfizer-BioNTech), the most commonly administered COVID-19 vaccine in Brazil (Ministério da Saúde, 2024). For more details on how vaccine efficacy in preventing infection (*v*_*e*_) and vaccine protection against death (*v*_*p*_) are derived, please see Derivation of vaccination parameters, supplementary materials.

A graphical representation of the model specifying the roles of each of these pathways is displayed in Figure 2.

**Fig. 2.**
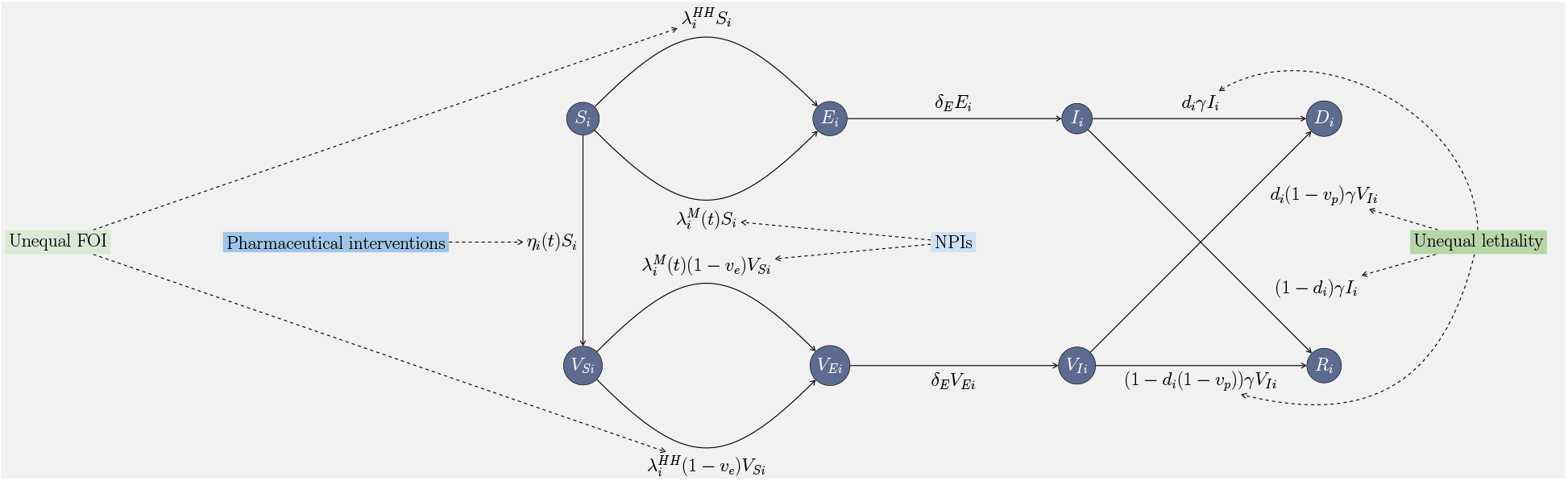
Model diagram.

### 2.4. Model output analysis

I initialize the model setting the whole population of each of the 5,565 municipalities as susceptible (*S*_*i*_ = municipality *i* 2020 midyear population estimate) with 10 exposed cases in the municipality of São Paulo, the location where the actual first cases of COVID-19 in Brazil were detected after being introduced from abroad via air travel (Candido et al., 2020; de Souza et al., 2020; Mee et al., 2022; Nicolelis et al., 2021; Serdan et al., 2020). I run iterations of the model using R (R Core Team, 2021) with the package *deSolve* (Karline Soetaert et al., 2010) for the counterfactual scenarios listed in Table 3.

**Table 3.** Counterfactual model scenarios for different NPI and vaccine distribution/adoption strategies.

| Scenario | Interventions implemented | NPI strategy | Vaccine strategy |
| --- | --- | --- | --- |
| 1 | None (*baseline*) |  |  |
| 2 | NPIs | As in real world |  |
| 3 | Vaccination |  | As in real world |
| 4 | NPIs & Vaccination | As in real world | As in real world |
| 5 | NPIs | Equal<sup>a</sup> |  |
| 6 | Vaccination |  | Equal<sup>a</sup> |
| 7 | Vaccination |  | Equitable<sup>b</sup> |
| 8 | NPIs & Vaccination | Equal<sup>a</sup> | Equal<sup>a</sup> |
| 9 | NPIs & Vaccination | Equal<sup>a</sup> | Equitable<sup>b</sup> |

*Notes:*
<sup>a</sup> All municipalities have the same adoption as the country as a whole.
<sup>b</sup> Lower SES municipalities prioritized for intervention. (Given real-world country-level vaccination rates, municipalities are fully vaccinated in ascending order by SES.)

Each simulation is run for 182 days (26 weeks/6 months, or half a year), from March 1 - August 29, 2020. Contemporaneous data for the time-variant mobility parameters (*ω*_*ij*_(*t*) and Δ*M*_*i*_(*t*)) are used, while the time-variant vaccination parameter (*η*_*i*_(*t*)) uses data from the first six months of the vaccination campaign. I obtain deaths in each municipality, *D*_*i*_ at each day *t*. Each day is part of an epidemiological week *w*, such that *D*_*iw*_ = ∑_*t*∈*w*_ *D*_*it*_, which is rounded to the nearest whole number. The number of person-years in each epidemiological week, *PY*_*iw*_, is calculated by dividing municipality *i*’s midyear 2020 population estimate by 53 since there were 53 epidemiological weeks in the epidemiological year 2020 (Grolemund & Wickham, 2011). For each municipality, I obtain two indicators of SES frequently used in the literature, the social vulnerability index (SVI) and the municipal human development index (MHDI) (ipea- Instituto de Pesquisa Econômica Aplicada, 2022). These indicators go in opposite directions, with a higher SVI indicating greater social vulnerability and thus lower SES, while a higher MHDI indicates greater human development and thus higher SES. See Tables 4 and 5 for how these indicators are derived, respectively.

**Table 4.**
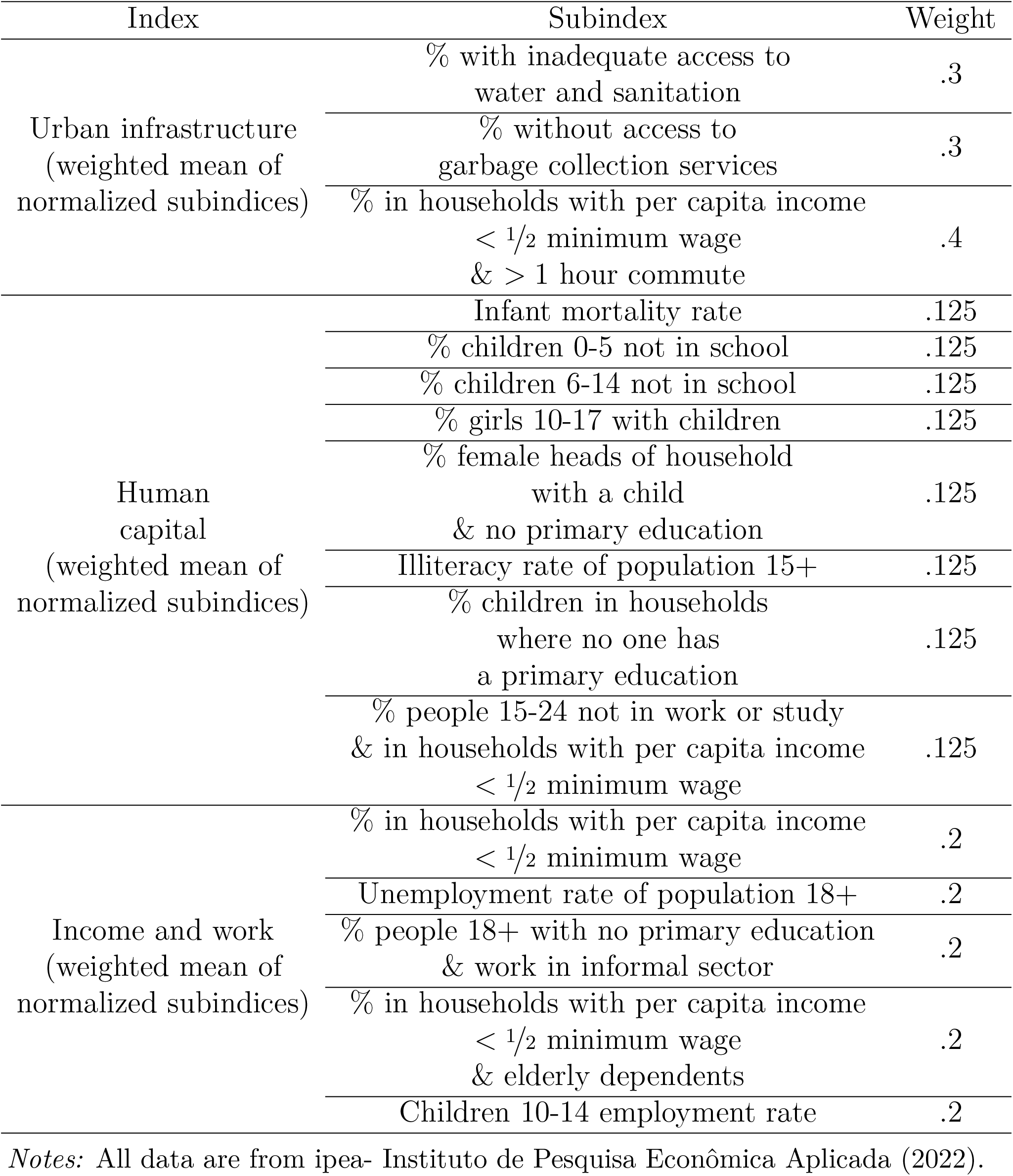
Social Vulnerability Index (SVI)- calculated by the arithmetic mean of its component indices.

**Table 5.**
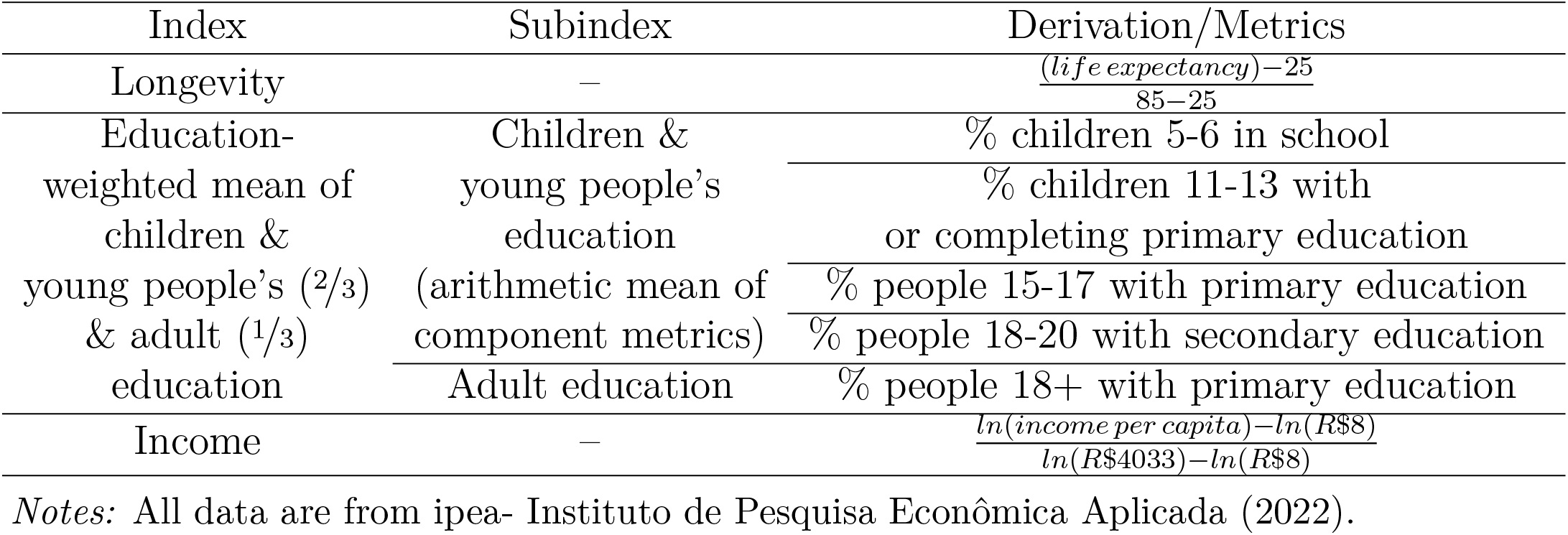
Municipal Human Development Index (MHDI)- calculated by the geometric mean of its component indices.

For the weekly mortality time series produced by each simulation, I analyze how SES gradients in mortality evolve over time by calculating the relative risk (RR) of mortality in municipalities in the 5th compared to the 1st quintile of the (inverse) SVI and the MHDI for each week *w*. I also replicate these analyses using the empirical weekly time series of COVID-19 mortality in Brazil covering the same March 1 - August 29 time period. Since NPIs had been implemented during this period in Brazil but vaccinations had not yet been rolled out, the results should be similar to those obtained in the scenario with NPI adoption following real-world rates (scenario 2, Table 3).

To account for confounders of the relationship between municipality-level SES and weekly mortality, I construct a negative binomial regression model with a log link and an offset for person-years, specified as:

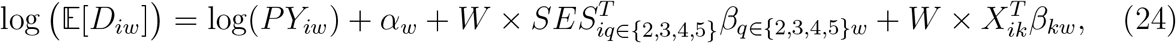

where *SES*_*iq*∈{2,3,4,5}_ is a set of dummy variables for quintiles 2, 3, 4, and 5 of the SVI or MHDI (with quintile 1 as the reference category), interacted with epidemiological week *W*. log(*PY*_*iw*_) is an offset, and *α*_*w*_ denotes week fixed effects. *X*_*ik*_ is a vector of additional municipality-level controls that are also interacted with week *W*. These controls include the age distribution (proportion of the population aged over 50) (Ministério da Saúde, 2021) and intra-municipality income inequality measured by the Gini coefficient (ipea-Instituto de Pesquisa Econômica Aplicada, 2022). I assume any bias in age distribution estimates is proportional across municipalities. As robustness checks, I additionally control for region and state fixed effects and for political behavior, measured by mean municipality-level first-round vote share for President Jair Bolsonaro in the 2018 and 2022 presidential elections (Justiça Eleitoral, 2022). Prior studies have identified political behavior as a confounder of the relationship between SES and COVID-19 mortality, as municipalities with greater support for Bolsonaro tend to have higher SES while also exhibiting lower adoption of protective measures (NPIs and vaccination), ceteris paribus (Rache et al., 2021; Razafindrakoto et al., 2022, 2024; Seara-Morais et al., 2023; Xavier et al., 2022).

I fit these models using the *fixest* package in R (Bergé, 2018). Standard errors are computed using Driscoll–Kraay robust covariance estimators to account for serial correlation over time and cross-sectional dependence across municipalities (Driscoll & Kraay, 1998). Relative risks (RRs) of mortality for municipalities in the least vulnerable quintile (Q5) compared to the most vulnerable quintile (Q1) are obtained by exponentiating the estimated coefficients *β*_*q*=5,*w*_. Ninety-five percent confidence intervals are constructed on the log scale and transformed to the relative-risk scale. An *RR <* 1 indicates higher mortality risk in the most vulnerable municipalities (an expected social gradient), an *RR >* 1 indicates an inverted gradient, and *RR* = 1 indicates no mortality gradient. The modeling architecture is modular, allowing alternative structural indicators, intervention scenarios, and epidemiological parameters to be substituted in other national or disease contexts.

## 3. Results

To contextualize the scenario results, we begin by summarizing the empirical distributions of the model parameters corresponding to the four pathways to inequality (Fig. 1). Pre-existing inequities in exposure are reflected in substantially larger estimated mean household contact rates in the most vulnerable (Q1) compared to the least vulnerable (Q5) municipalities (Fig. 3). Pre-existing inequities in outcomes are captured by higher estimated IFRs in the most vulnerable municipalities (Fig. 4), although these disparities are less pronounced than those observed for household contacts. Some extreme IFR values are observed across all SES groups, reflecting the fact that some municipalities reported only a small number of cases, and that cases and deaths are discrete events.

**Fig. 3.**
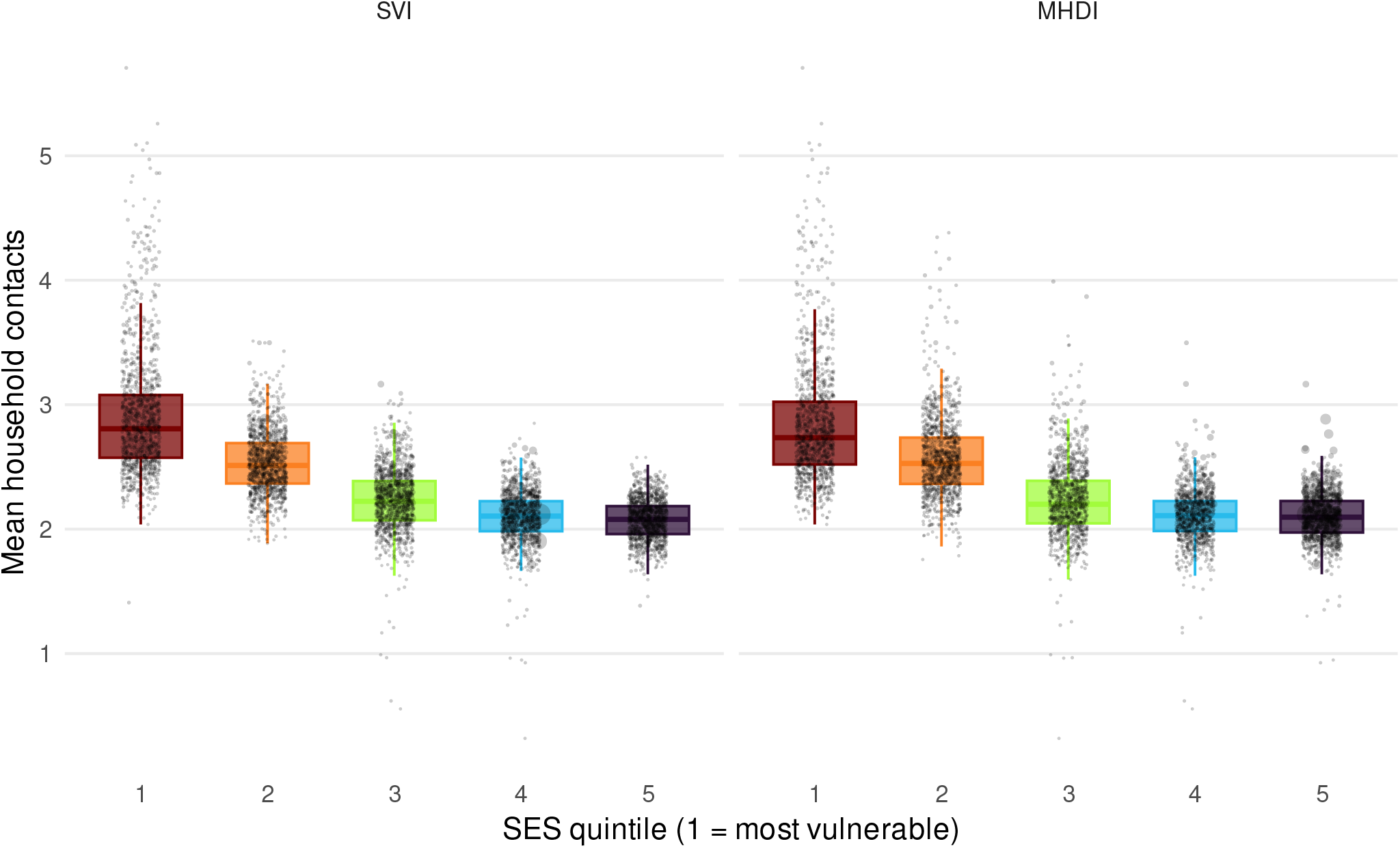
Distribution of estimated mean household contact rates by municipality SES quintile, by inverse SVI and MHDI.

**Fig. 4.**
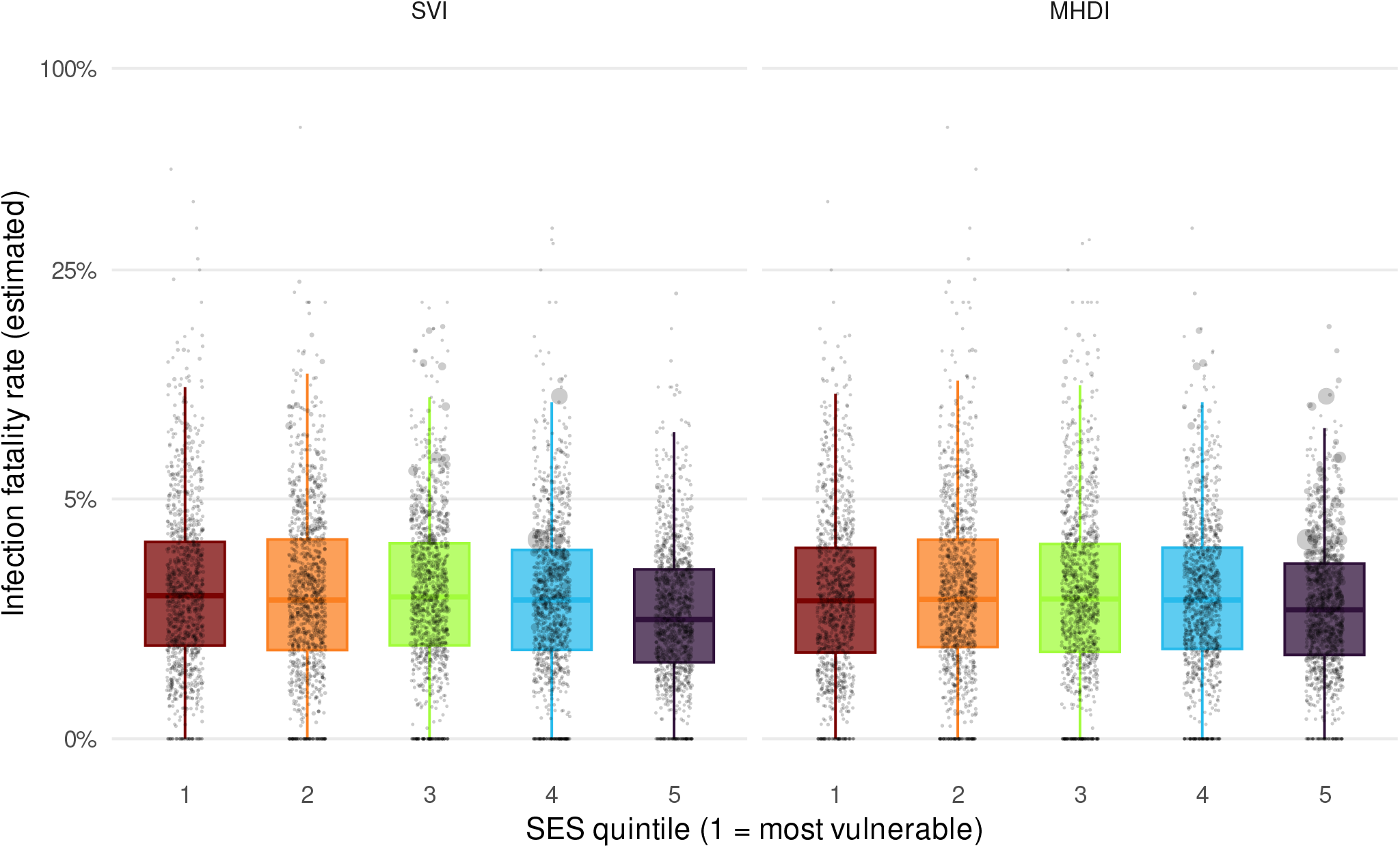
Distribution of estimated IFRs by municipality SES quintile, by inverse SVI and MHDI.

Inequalities in exposure arising from differential uptake of disease-specific interventions are reflected in SES-stratified mobility reductions following the onset of the pandemic (Fig. 5). Residents of the least vulnerable municipalities reduced their mobility more sharply and sustained lower mobility levels for a longer period compared to residents of the most vulnerable municipalities. However, the SES–mobility relationship differs by index: while mobility reductions are monotonic with SES when measured using the MHDI, this pattern is less consistent for the SVI. In particular, mobility in the least vulnerable municipalities rebounded sharply after the initial decline, briefly converging with levels observed in the most vulnerable municipalities, before the latter continued increasing their mobility and returned to pre-pandemic levels by August.

**Fig. 5.**
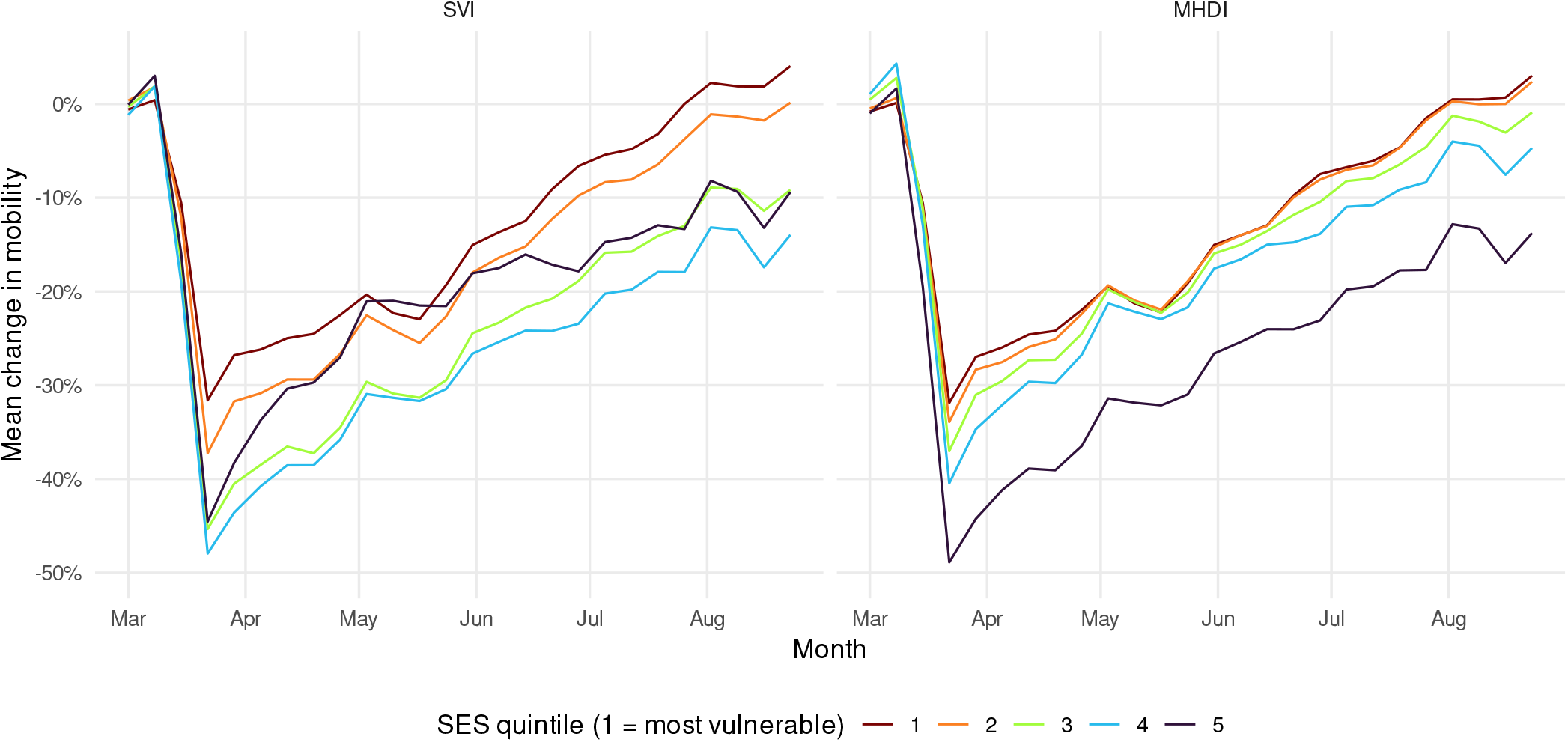
Proportional change in mobility over time following the onset of the COVID-19 pandemic by municipality SES quintile, by inverse SVI and MHDI. Estimated using Facebook Data for Good Movement Range Maps (COVID-19 Mobility Data Network, 2020; Meta, 2023).

Finally, inequalities in outcomes arising from differential uptake of disease-specific interventions are captured by disparities in vaccination uptake (Fig. 6). Cumulative vaccine coverage increases almost monotonically with SES over time. For the purpose of comparing counterfactual scenarios, vaccination is modeled as if it were available from the start of the simulations on March 1, 2020, rather than from January 16, 2021 as in the real-world Brazilian vaccination campaign. Together, these parameter distributions define the initial conditions and intervention heterogeneity that shape the temporal evolution of mortality gradients across empirical and simulated scenarios.

**Fig. 6.**
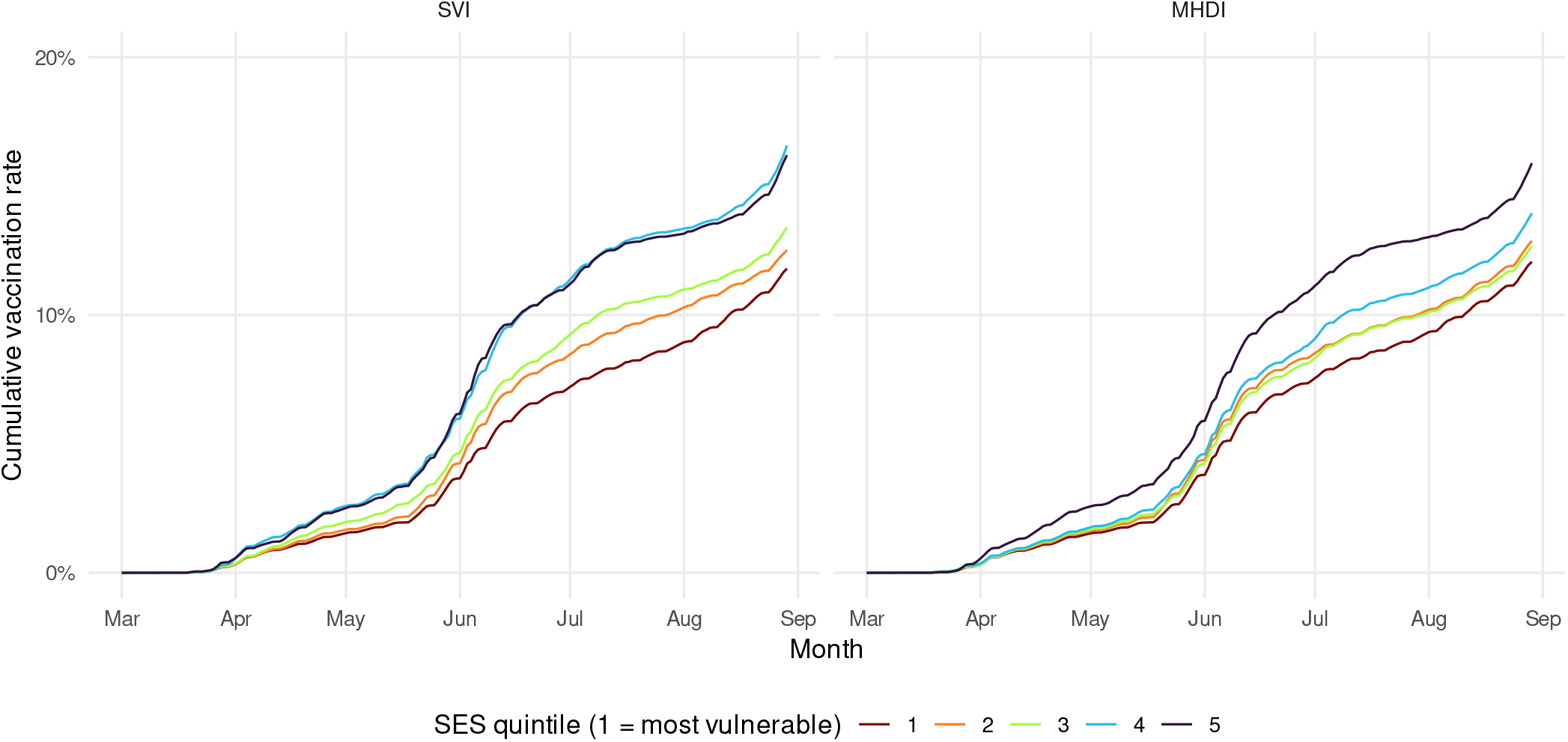
Cumulative COVID-19 vaccination uptake by municipality SES quintile, by inverse SVI and MHDI. Vaccination uptake is temporally transposed to align with the counterfactual scenario in which the vaccination campaign started on March 1, 2020 rather than January 16, 2021.

Based on the empirical mortality data, controlling for age distribution and intra-municipality inequality, the earliest phase of the COVID-19 pandemic in Brazil was characterized by a higher risk of mortality in the least vulnerable municipalities compared to the most vulnerable, as measured by the SVI (*RR >* 1; Fig. 7), indicating an initially inverted mortality gradient. As the outbreak progressed, the estimated relative risk fluctuated between inverted (*RR >* 1) and expected (*RR <* 1) gradients, before stabilizing in the expected direction by approximately the seventh epidemiological week. After this point, the relative risk gradually increased toward parity, crossing RR = 1 again by week 16. This pattern reflects a common dynamic in emerging epidemics, where early transmission occurs through highly connected populations before diffusing to socially vulnerable populations.

**Fig. 7.**
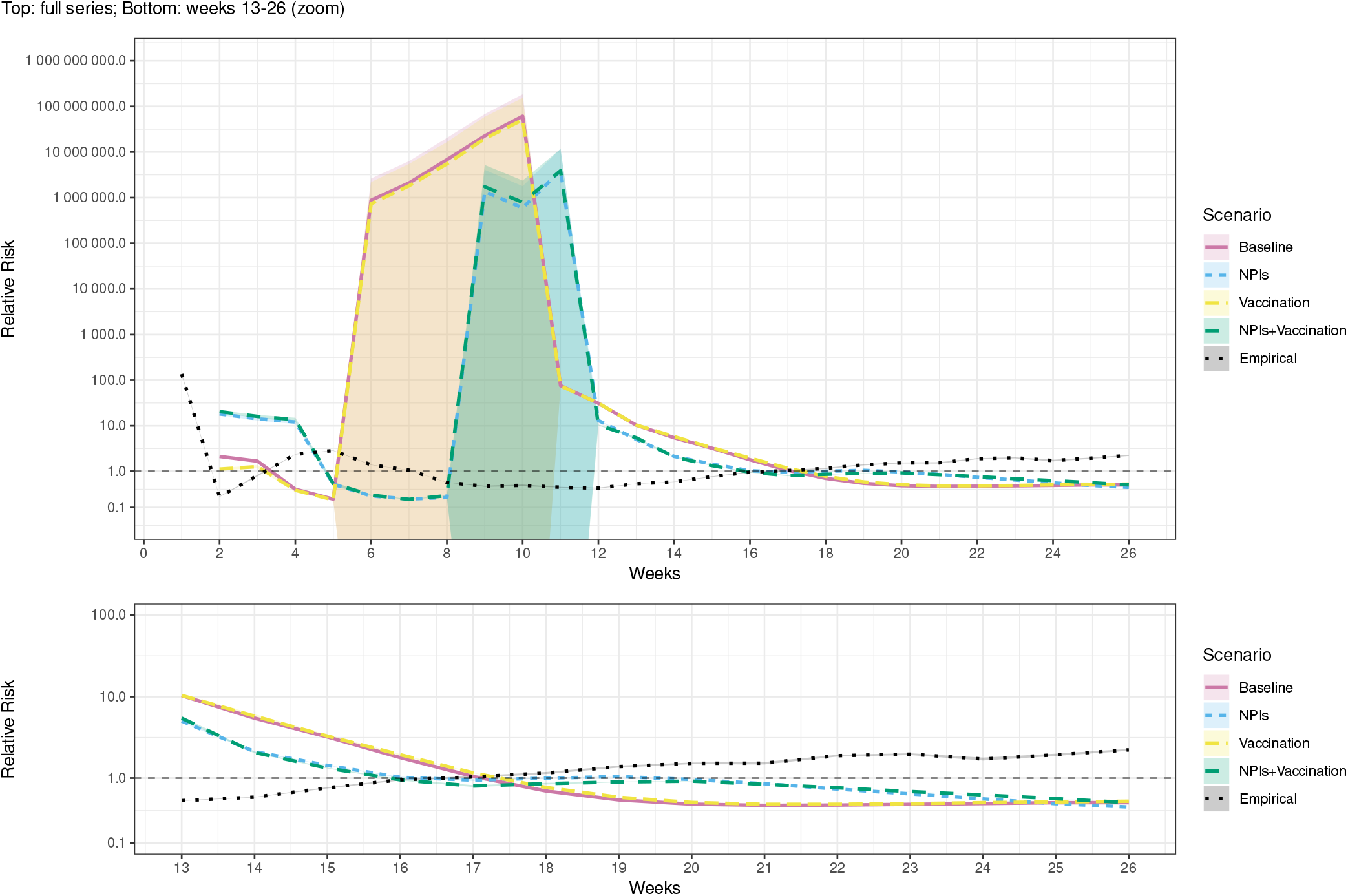
Relative risk of mortality in the least vulnerable (Q5) compared to the most vulnerable (Q1) municipalities by SVI during the first 26 weeks of an outbreak in the real world (*Empirical*) and simulations of scenarios 1-4 (Table 3). Shaded bands indicate 95% confidence intervals.

In the earliest weeks of the simulations, deaths are rare (Figs. A.1-A.18). For the first several weeks, the most vulnerable group records zero deaths while the least vulnerable group records nonzero deaths, yielding weakly identified RR estimates and very wide uncertainty. Early-week estimates are interpreted primarily in terms of the direction of the gradient, while comparisons of magnitude and inter-scenario differences are not interpretable until mortality is observed in both groups. Across scenarios, inflection points in RR largely reflect the transition from early seeding in highly connected, less vulnerable municipalities to broader diffusion into more vulnerable municipalities. This diffusion process, combined with weak early identification, explains the pronounced fluctuations observed in the simulated RR curves during the initial phase of the pandemic. Accordingly, all simulation figures include zoomed panels focusing on the second half of the simulation period (weeks 13–26), where relative risk estimates are more stable and suitable for inter-scenario comparison.

Scenario 1 (*Baseline*, no NPIs nor vaccination, Table 3) thus begins with an inverted mortality gradient. By weeks 11–12 (around the 3rd month), the ratio falls toward 1, marking a shift towards an expected gradient. Once this crossover occurs (by the 4th month), the model sustains an expected gradient through the remainder of the six-month simulation. This transition is accelerated in scenarios with NPIs (scenarios 2, *NPIs* and 4, *NPIs + Vaccination*), but after faster initial declines their RR curves plateau around 1 for several weeks as NPIs impede the transmission and geographic diffusion of COVID, before settling into expected mortality gradients. Conversely, vaccination alone does not appear to have much of an effect (scenario 3, *Vaccination*), and only very slightly compounds the impact of NPIs when the two interventions are combined (scenario 4).

Despite surface-level similarities, the scenario with NPIs adopted according to observed patterns (scenario 2) does not fully reproduce the empirical RR trajectory. While it does capture the overall shape of the empirical mortality gradient once it becomes well-defined, it lags real-world dynamics by several weeks and does not exhibit the gradual return toward parity observed in the empirical data. In the simulations, once expected mortality gradients emerge, they remain largely stable across scenarios, in contrast to the empirical RR curve, which continues to trend upward over time. These divergences likely reflect earlier community transmission prior to the simulation start date, as well as features of the real-world pandemic not fully captured by mobility-based proxies for NPI uptake, including shifts in contact patterns over time and the effects of unmodeled interventions such as masking. Waning immunity, which is not represented in the model, may also have contributed to the attenuation or partial re-inversion of empirical gradients later in the study period. Measurement limitations in early mortality surveillance and residual confounding related to political behavior may further contribute to these differences. We return to these considerations in the Discussion.

Comparable patterns are observed when analyzing the mortality gradients of these same scenarios using MHDI instead of inverse SVI (Fig. 8), but with the NPI scenarios remaining plateaued around 1 for longer periods of time. However, the RR curve of the NPI scenario differs even further from the empirical RR curve, as the latter never falls below 1. This implies a consistently inverted social gradient in real-world mortality with respect to MHDI during the first six months of the pandemic, even after accounting for age structure and intra-municipality inequality. This discrepancy highlights that empirical COVID-19 mortality gradients observed in Brazil can depend on the SES index employed.

**Fig. 8.**
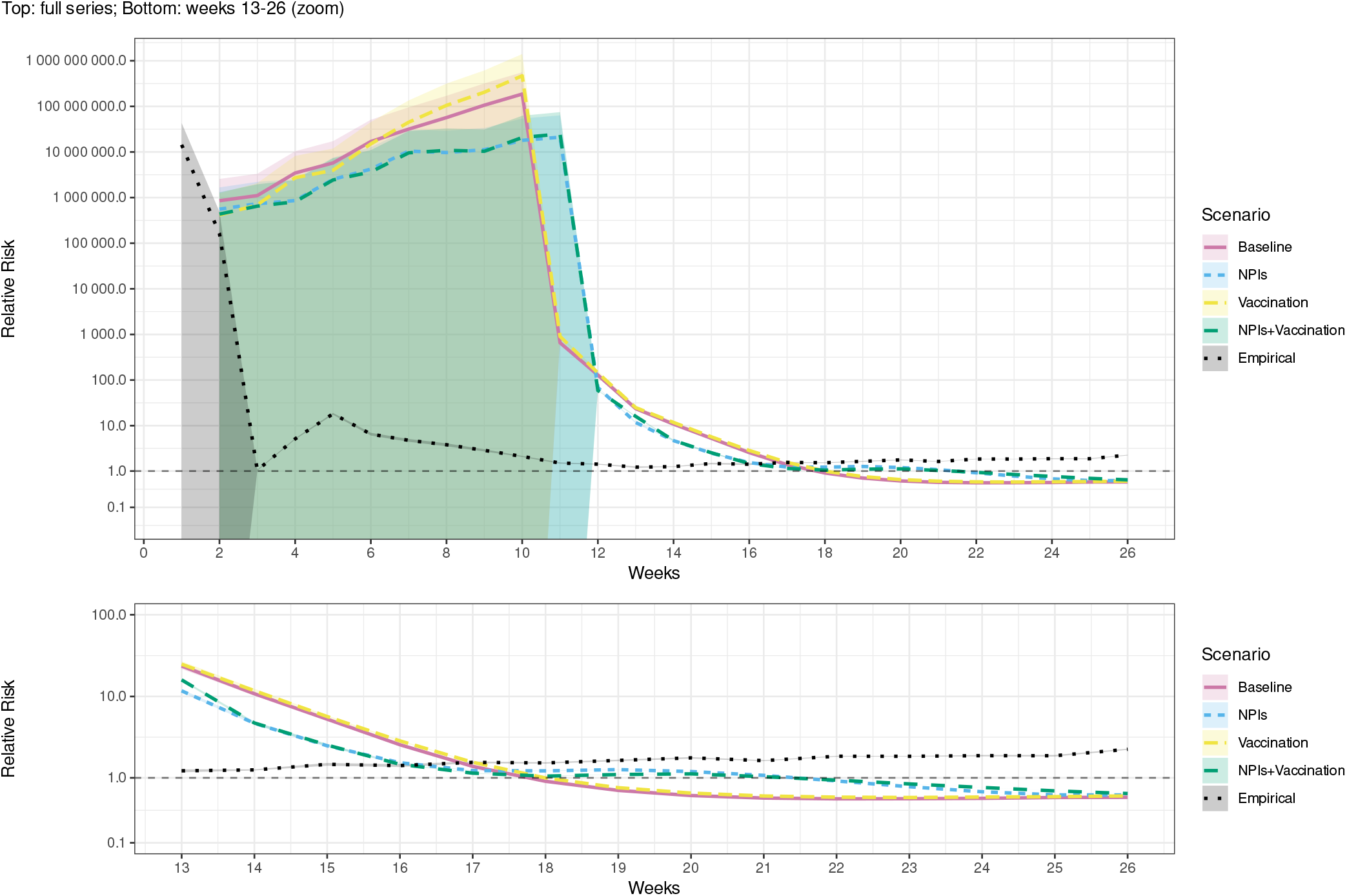
Relative risk of mortality in the least vulnerable (Q5) compared to the most vulnerable (Q1) municipalities by MHDI during the first 26 weeks of an outbreak in the real world (*Empirical*) and simulations of scenarios 1-4 (Table 3). Shaded bands indicate 95% confidence intervals.

Similar dynamics are observed between the SVI and MHDI RR curves for the counter-factual scenarios with alternative intervention distribution strategies (scenarios 5-9, Table 3), but with those of the MHDI shifted up and to the right compared to SVI (Fig. 9 and 10). Equal adoption of NPIs (scenarios 5, *NPIs-equal* and 8, *NPIs + Vaccination-equal*), whereby all municipalities reduce their mobility by the same amount (Fig. A.19), shifts the RR curve to the right compared to the baseline scenario while largely maintaining its shape, pushing back the time until a stable expected mortality gradient of similar magnitude is achieved by several weeks. In contrast, the implementation of equal vaccination (scenarios 6, *Vaccination-equal* and 8), whereby all municipalities are subject to the same rates of vaccine uptake (Fig. A.20), only slightly diminishes the magnitude of the expected mortality gradient. However, prioritizing the lowest SES municipalities for vaccination (scenario 7, *Vaccination-equitable*), where municipalities are fully vaccinated in descending order according to vulnerability given the national rate of vaccine administration on each given date (Fig. A.21), shifts the RR curve up, yielding a more strongly inverted mortality gradient for most of the duration of the simulation, though it does drop below 1 in the final weeks.

**Fig. 9.**
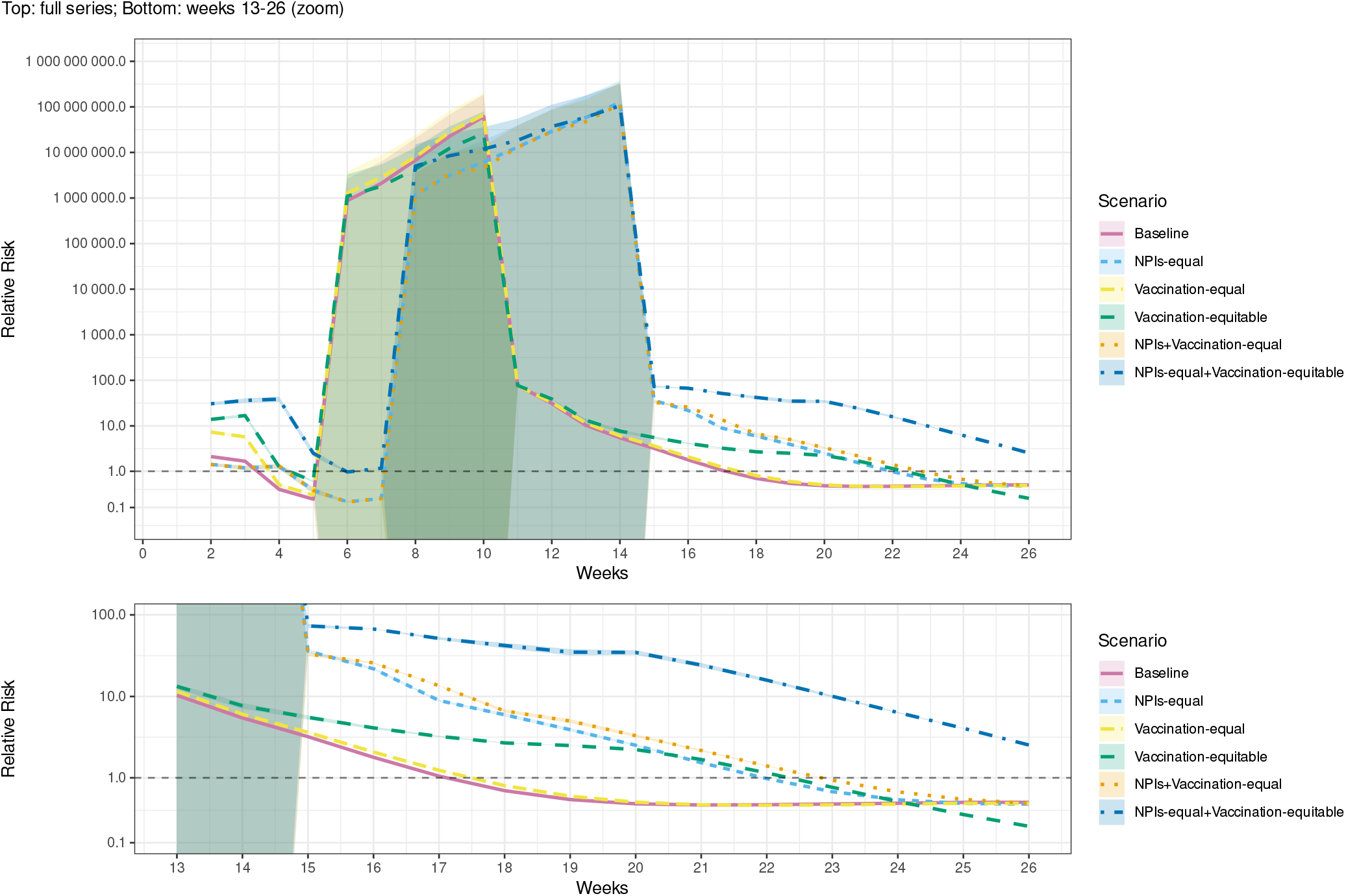
Relative risk of mortality in the least vulnerable (Q5) compared to the most vulnerable (Q1) municipalities by SVI during the first 26 weeks of an outbreak in simulations of scenarios 1 and 5-9 (Table 3). Shaded bands indicate 95% confidence intervals.

**Fig. 10.**
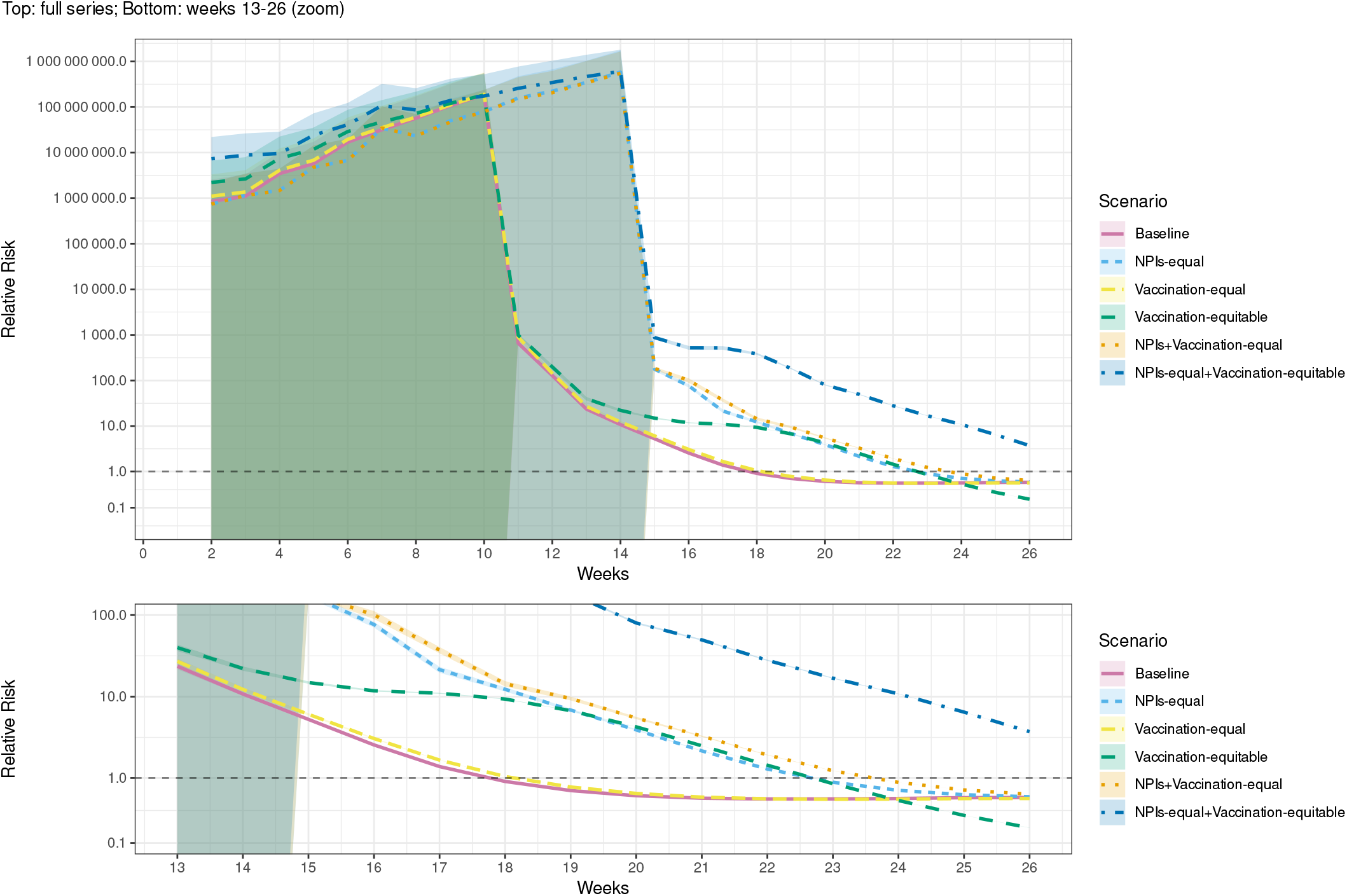
Relative risk of mortality in the least vulnerable (Q5) compared to the most vulnerable (Q1) municipalities by MHDI during the first 26 weeks of an outbreak in simulations of scenarios 1 and 5-9 (Table 3). Shaded bands indicate 95% confidence intervals.

The scenario that sees the greatest inversion of the mortality gradient is when both equal NPIs and equitable vaccination are implemented (scenario 9, *NPIs-equal + Vaccination-equitable*). The effects of these interventions reinforce each other, shifting the RR curve dramatically upwards and to the right, avoiding the emergence of the expected mortality gradient observed in all other scenarios. However, even with these highly redistributive strategies, the mortality gradient still begins converging back towards the expected direction by the end of the simulation. This highlights the difficulty of avoiding expected social gradients in mortality if underlying pre-existing inequities remain unchanged. Qualitatively similar results, especially the equal NPI-equitable vaccination scenario having the most-inverted mortality gradient, are largely observed across the sensitivity analyses and robustness checks, though with a greater degree of uncertainty in the RR estimates, particularly in the latter (Figs. A.22-A.73).

## 4. Discussion

These results suggest that expected social gradients in mortality during emerging infectious disease epidemics arise primarily from disease-agnostic pre-existing inequities in living conditions, health status, and healthcare access and quality. The Brazilian COVID-19 case provides a real-world illustration of how these structural inequalities can shape the development of mortality gradients even before disease-specific interventions are introduced. In this case, the expected mortality gradient would most likely have developed even in the absence of unequally adopted disease-specific interventions; the NPIs implemented contemporaneously with disease introduction were likely not the source of this expected mortality gradient, but merely reinforced it and accelerated its development. Any initially observed inverted social gradient in COVID-19 mortality was thus likely an artifact of earlier introduction of the disease to highly connected, high-SES urban centers (Albuquerque et al., 2021; Candido et al., 2020; Castro et al., 2021; da Silva et al., 2022; de Souza et al., 2020; Fortaleza et al., 2020; Mee et al., 2022; Nicolelis et al., 2021; Serdan et al., 2020). These findings help fill a gap in the empirical literature, providing evidence contrary to COVID-19 having been subject to “natural mortality” before the introduction of social distancing NPIs (Clouston et al., 2016, 2021), and evidence supporting all pathways having played a role in producing the expected mortality gradient (Bambra, 2022) (Fig. 1). More broadly, these findings suggest that the emergence of expected mortality gradients during epidemic outbreaks may often be driven primarily by structural inequalities that shape exposure and vulnerability, while the unequal adoption of disease-specific interventions acts to reinforce or accelerate these dynamics rather than initiate them.

However, the emergence of expected mortality gradients during epidemics may be delayed or even reversed through intervention strategies that prioritize the most socioeconomically vulnerable populations. Although NPIs alone are unlikely to reverse the emergence of expected mortality gradients, they can impede this process by reducing exposure and slowing transmission, thereby delaying the diffusion of infection down the socioeconomic gradient through socially stratified mobility network structures (Licciardi & Monteiro, 2022). This delaying effect is most clearly produced if NPIs are adopted equally, buying time for other interventions that protect or improve the outcomes of those exposed to the disease to be distributed to vulnerable populations. In the Brazilian COVID-19 case examined here, this role is played by vaccination. However, merely distributing vaccines equally across populations is not sufficient; to meaningfully alter mortality gradients they must be targeted toward the most socioeconomically vulnerable populations. Even then, however, such equitable intervention strategies cannot fully overcome deep-rooted structural inequities embedded in living conditions, healthcare availability, and baseline health status.

Nevertheless, real-world empirical trends sometimes diverged from our simulation results. Several factors may help explain why. For instance, the empirical expected mortality gradient for the SVI re-inverting in the fifth month of the outbreak could reflect waning immunity, a phenomenon omitted in the model. It is also possible that unobserved factors not captured by the regression controls caused mortality risk to systematically shift to higher SES municipalities at this particular point in time. Indeed, previous studies have found that the social gradient in COVID-19 mortality in Brazil diminished or inverted from July/August, 2020 (Razafindrakoto et al., 2022; Rocha et al., 2021). This can, at least in part, be attributed to decreasing compliance with NPIs in higher SES municipalities over time, which is already captured in the simulations. This in turn can partially be explained by support for Jair Bolsonaro confounding the relationship between SES and the adoption of protective interventions (Rache et al., 2021; Razafindrakoto et al., 2024; Seara-Morais et al., 2023). Controlling for Bolsonaro vote share in the regression model robustness checks does shift the empirical RR curve down, as well as the RR curves of the simulations of scenarios with interventions adopted according to real-world patterns, indicating a more concentrated risk of death in the lowest SES municipalities. While this pushes back the time until the empirical mortality gradient re-inverts by several weeks, it does not entirely prevent this re-inversion from occurring during the final weeks of the study period. Further investigating the role of political behavior as a confounder of the relationship between SES and infectious disease mortality could be a valuable direction for future research.

By contrast, the persistent inversion of the empirical gradient when measured using the MHDI may reflect its components being less salient social determinants of COVID-19 mortality than those comprising the SVI. In fact, previous work has paradoxically found simultaneous positive associations between both higher municipal human development (da Silva et al., 2022; Fortaleza et al., 2020) and greater social vulnerability and mortality (Albuquerque et al., 2021; Andrade et al., 2022; Martins-Filho et al., 2021). Positive associations between the specific components of SVI and mortality have been found, especially with respect to lack of basic education (illiteracy and deficiency of primary education), poor environmental and housing conditions (poor access to water, sanitation, and garbage collection), and poverty (income relative to the minimum wage) (Passos et al., 2021; K. B. Ribeiro et al., 2021; Silva & Ribeiro-Alves, 2021). Simultaneously, Razafindrakoto et al. (2022) and Baggio et al. (2021) have found total absolute income and the education sub-index of MHDI (which takes primary and secondary education levels but not literacy into account) to be associated with higher mortality, even while poverty, the human capital and urban infrastructure sub-indices of SVI (Table 4), and poor health, living, and working conditions remained risk factors. Nassif Pires et al. (2020) further demonstrate that while lower income and lack of educational attainment above the primary level are associated with poorer living, working, and health conditions, it is ultimately these latter conditions that are the proximal drivers of mortality. Consequently, the body of evidence suggests that it is not overall income and education levels per se, but rather their absolute deprivation, as well as associated absolute deprivation with respect to housing, environmental and working conditions, that are the most salient drivers of inequities in the risk of exposure to and severe outcomes of COVID-19 in Brazil. All of these factors are captured by the SVI, but not by the MHDI (Table 5). In future work, more comprehensive, theory-driven measurements of SES composed of the most relevant social determinants of mortality could be explored, inspired by previous research using principal component analysis, Alkire-Foster, and fuzzy-set approaches (Bermudi et al., 2021; Rocha et al., 2021; Tavares & Betti, 2021).

The simulated mortality gradient of the NPI scenario for the SVI lagging behind the empirical mortality gradient could be explained by Brazil’s real-world outbreak having started markedly earlier than the March 1st date used in the simulations. With limited testing, the first COVID-19 cases in Brazil were probably not reported (de Souza et al., 2020; Mee et al., 2022), and according to phylogenetic analyses could have been introduced as far back as February 17th (Candido et al., 2020). By early March, community transmission was likely already well-established.

Additionally, although based on real-world data, the parameterization of NPIs in my model suffers from limitations hindering its ability to capture real-world dynamics. Since I only have data on proportional changes in mobility by municipality (COVID-19 Mobility Data Network, 2020; Meta, 2023) compared to a pre-pandemic baseline where mobility levels were similar across socioeconomic groups (Brotherhood et al., 2022; Li et al., 2021; Rocha et al., 2021), I assume a uniform baseline rate of potentially infectious extra-household contacts across municipalities, that any changes to this contact rate are directly proportional to changes in mobility, and that the probability of infection from these contacts is constant. However, this fails to take into account how both rates of contact outside the household and probability of infection could vary across municipalities independent of mobility and how they may also be affected by unequally adopted NPIs.

For example, previous literature has found positive associations between living in more densely populated municipalities, urban areas, and favelas, and COVID-19 incidence and resulting mortality, suggesting higher rates of potentially infectious contacts in the community, that are independent of mobility and housing conditions (Albuquerque et al., 2021; Brotherhood et al., 2022; Fortaleza et al., 2020; Mee et al., 2022; Razafindrakoto et al., 2022; Xavier et al., 2022). Furthermore, though populations with different SES may have had similar overall levels of mobility pre-pandemic, the modes of transportation by which this mobility occurred, with different associated contact rates, varied. Lower SES populations were more likely to walk, cycle, or use public transportation (Sá et al., 2016), while higher SES populations were more likely to use private automobiles or fly (Candido et al., 2020; G. R. Ribeiro et al., 2014). The risk of infection from contacts potentially also varied across these populations. Of particular relevance is evidence from the US that lower income workers were more likely to work in occupations with high personal proximity to others, even before the pandemic (Mongey & Weinberg, 2020).

NPIs implemented in Brazil at the beginning of the pandemic independent of mobility included, but were not limited to, the use of face masks and other personal protective equipment, and physical distancing guidelines (e.g. 1.5-2m) (de Souza Santos et al., 2021; Faria de Moura Villela et al., 2021; Razafindrakoto et al., 2022; Rocha et al., 2021). These interventions aimed to reduce the probability of infection from each contact and are not captured by the mobility reduction parameter in my model. While there are cross-sectional data estimating compliance with these measures, in which an educational gradient is identified (Faria de Moura Villela et al., 2021), unfortunately no longitudinal municipality-level data exist to parameterize them.

Challenges in parameterizing other pathways to COVID-19 mortality gradients should also be noted, particularly unequal lethality via the IFR parameter. Estimation of municipality-level IFRs is sensitive to small-population municipalities where random fluctuations can inflate or deflate estimates since cases and deaths are discrete events. Additionally, measurement error in case and death data could introduce further uncertainties in IFR estimates that differentially affect municipalities. These potential sources of error may thus bias the IFR parameter estimates, though I attempt to mitigate them through aggregating data throughout all of 2020 and conducting sensitivity analyses. While aggregating smaller municipalities for IFR estimation could potentially further address some of these sources of error, this would come at the expense of using as much municipality-level real-world data as possible to parameterize my model.

Although this study focuses on the COVID-19 pandemic in Brazil, the framework developed here is intended to be applicable across epidemic contexts in which structural inequality shapes exposure and intervention uptake. Ultimately, if unequal distribution and adoption of social distancing and other interventions reinforce the effects of disease-agnostic pre-existing inequities and further entrench expected mortality gradients, then alternative intervention distribution and uptake strategies to counteract their effects should be considered. The uptake of the social distancing NPIs modeled in this study, where individuals reduce their mobility and rates of contact, is nonrivalrous in that their uptake by one person does not impact another’s ability to do so (Feachem et al., 2002). Therefore a strategy encouraging their equal adoption by everyone would be most just in terms of fairness (Rawls, 1958). However, sometimes a strategy pursuing equality is not enough, especially when pre-existing social determinants of mortality are already highly unequal, as in the Brazilian context and many other settings characterized by deep socioeconomic disparities. Since vaccines are a rivalrous good, prioritizing some populations over others in the face of limited supply in pursuit of a specific public health goal could be justified. The same is true of other rivalrous public health interventions not modeled in this study, including mobility-independent NPIs, such as the distribution of masks and other personal protective equipment, testing, and ventilation upgrades, and treatments. Thus, a strategy pursuing equitable distribution of rivalrous interventions like vaccines, where the lowest SES municipalities are prioritized, in combination with equal adoption of nonrivalrous interventions like social distancing NPIs, would not only be the most effective in achieving the goal of mitigating the emergence of expected mortality gradients in EID epidemics, but also the most just. In future EID epidemics, to effectively minimize mortality inequalities, policymakers should devise equitable public health intervention strategies on the national and global level that prioritize the most disadvantaged and at-risk populations.

## Data Availability

All data produced are available online at

https://github.com/jordan-klein/covid19-mortality-gradients-brazil

https://doi.org/10.5281/zenodo.15272578

## Declaration of generative AI and AI-assisted technologies in the manuscript preparation process

During the preparation of this work the author used ChatGPT in order to review, proofread, and improve the clarity, conciseness, and flow of the writing. After using this tool/service, the author reviewed and edited the content as needed and takes full responsibility for the content of the published article.

## Acknowledgments

I would like to thank Marcia Castro, Cassio Turra, Sabrina Li, Amac Herdagdelen, Ridhi Kashyap, Doug Leasure, Sam Abbott, Carl Pearson, Brianna Rock, Pascal Klamser, Dirk Brockmann, Auss Aboud, and especially Filiz Garip, Jessica Metcalf, Deborah Balk, and Noreen Goldman, as well as the members of the Digital and Computational Demography Department at the Max Planck Institute for Demographic Research, and the IUSSP Epidemics and Contagious Diseases: The Legacy of the Past Workshop, University of São Paulo BIOMAT Seminar, Latin American Association of Population Conference, and University of Washington Computational Demography Working Group Seminar participants for all of their valuable input, advice, and feedback. I would also like to thank Pedro Peixoto for sharing the InLoco mobility data.

