## Supplement for "A Mechanistic Framework for Modeling Social Gradients in Emerging Infectious Disease Mortality: Evidence from Brazil"

#### A.1. Derivation of vaccination parameters

Individuals are considered vaccinated once they have had two vaccine doses. I assume everyone is given BNT162b2 (Pfizer-BioNTech).

The relative risk of all infection, both symptomatic and asymptomatic, in the vaccinated compared to the unvaccinated is .11 (Advisory Committee on Immunization Practices (ACIP), 2023). Accordingly:

$$v_e = 1 - .11 = .89 \tag{A.1}$$

Vaccine effectiveness against death in Brazil ages 20-59 is .945 and ages 60+ is .989 (dos Santos et al., 2023). The weighted average effectiveness based on the proportion of vaccinated individuals in Brazil in the 20-59 and 60+ age groups is:

$$\frac{.945(9,412,804 + 7,975,906 + 6,573,000 + 4,053,906) + .989(354,692 + 58,870 + 37,474)}{9,412,804 + 7,975,906 + 6,573,000 + 4,053,906 + 354,692 + 58,870 + 37,474} = .946 \tag{A.2}$$

Converting this into a relative risk is:

$$RR = 1 - .946 = .054 \tag{A.3}$$

To calculate vaccine protection against death in vaccinated individuals, $v_p$, I use Bayes' Theorem:

$$v_p = 1 - \frac{P(death \mid infection, vaccinated)}{P(death \mid infection, \neg vaccinated)} = 1 - \frac{\frac{P(infection|death,vaccinated)P(death|vaccinated)}{P(infection|vaccinated)}}{\frac{P(infection|death,\neg vaccinated)P(death|\neg vaccinated)}{P(infection|\neg vaccinated)}} \tag{A.4}$$

Since $1 = P(infection \mid death, vaccinated) = P(infection \mid death, \neg vaccinated)$, I can simplify the above to:

$$v_p = 1 - \frac{P(death|vaccinated) / P(death|\neg vaccinated)}{P(infection|vaccinated) / P(infection|\neg vaccinated)} \tag{A.5}$$

Substituting in the relative risks from (Advisory Committee on Immunization Practices (ACIP), 2023) and (dos Santos et al., 2023), I calculate $v_p$:

$$v_p = 1 - \frac{.054}{.11} = .509 \tag{A.6}$$

### A.2. Derivation of the basic reproduction number

I start with the infected subsystem of the model without vaccination.

$$\frac{dE_i}{dt} = S_i \tau_{HH} c_i^{HH} \frac{I_i}{S_i + E_i + I_i + R_i} + S_i \sum_j \tau_M c_i^M(t) \omega_{ij}(t) \frac{I_i}{S_i + E_i + I_i + R_i} - \delta_E E_i \quad \text{(A.7)}$$

$$\frac{dI_i}{dt} = \delta_E E_i - \gamma I_i \quad \text{(A.8)}$$

These equations can be re-expressed as transmission matrix $\mathscr{F}$, which defines how new infections enter the system, and transition matrix $\mathscr{V}$, which defines how individuals in the infected compartments transition between different states.

$$\mathscr{F} = \begin{bmatrix} diag(\mathbf{S})\tau_{HH} diag(c^{HH}) diag^{-1}(\mathbf{S}+\mathbf{E}+\mathbf{I}+\mathbf{R})\mathbf{I} + diag(\mathbf{S})\tau_M diag(c^M(t))\mathbf{\Omega}(t) diag^{-1}(\mathbf{S}+\mathbf{E}+\mathbf{I}+\mathbf{R})\mathbf{I} \\ 0 \end{bmatrix} \quad \text{(A.9)}$$

$$\mathscr{V} = \begin{bmatrix} diag(\delta_E)\mathbf{E} \\ diag(\gamma)\mathbf{I} - diag(\delta_E)\mathbf{E} \end{bmatrix} \quad \text{(A.10)}$$

I next derive the Jacobian matrices of $\mathscr{F}$ and $\mathscr{V}$ at the disease-free equilibrium for the null model, in which $t = 0$, no interventions are implemented, and contact rates are at their baseline levels.

$$F = \begin{bmatrix} 0 & \tau_{HH} diag(\mathbf{N}) diag(c^{HH}) diag^{-1}(\mathbf{N}) + \tau_M diag(\mathbf{N}) diag(c_0^M)\mathbf{\Omega}_0 diag^{-1}(\mathbf{N}) \\ 0 & 0 \end{bmatrix} \quad \text{(A.11)}$$

$$V = \begin{bmatrix} diag(\delta_E) & 0 \\ -diag(\delta_E) & diag(\gamma) \end{bmatrix} \quad \text{(A.12)}$$

The inverse of $V$ is the following:

$$V^{-1} = \begin{bmatrix} diag^{-1}(\delta_E) & 0 \\ diag^{-1}(\gamma) & diag^{-1}(\gamma) \end{bmatrix} \quad \text{(A.13)}$$

Finally, I calculate the next generation matrix $FV^{-1}$:

$$FV^{-1} = \begin{bmatrix} \mathbf{HH} diag^{-1}(\gamma) + \mathbf{M} diag^{-1}(\gamma) & \mathbf{HH} diag^{-1}(\gamma) + \mathbf{M} diag^{-1}(\gamma) \\ 0 & 0 \end{bmatrix} \quad \text{(A.14)}$$

where $\mathbf{HH} = \tau_{HH} diag(\mathbf{N}) diag(c^{HH}) diag^{-1}(\mathbf{N})$ and $\mathbf{M} = \tau_M diag(\mathbf{N}) diag(c_0^M)\mathbf{\Omega}_0 diag^{-1}(\mathbf{N})$. The basic reproduction number is the spectral radius of $FV^{-1}$.

$$R_0 = \rho(\mathbf{HH} diag^{-1}(\gamma) + \mathbf{M} diag^{-1}(\gamma)) \quad \text{(A.15)}$$

This can be split up into basic reproduction numbers for household and mobility-related transmission respectively, such that $R_0 = R_0^{HH} + R_0^M$.

$$R_0^{HH} = \frac{\tau_{HH}}{\gamma} \rho(diag(\mathbf{N}) diag(c^{HH}) diag^{-1}(\mathbf{N})) \tag{A.16}$$

$$R_0^{M} = \frac{\tau_{M}}{\gamma} \rho(diag(\mathbf{N}) diag(c_0^{M}) \mathbf{\Omega}_0 diag^{-1}(\mathbf{N})) \tag{A.17}$$

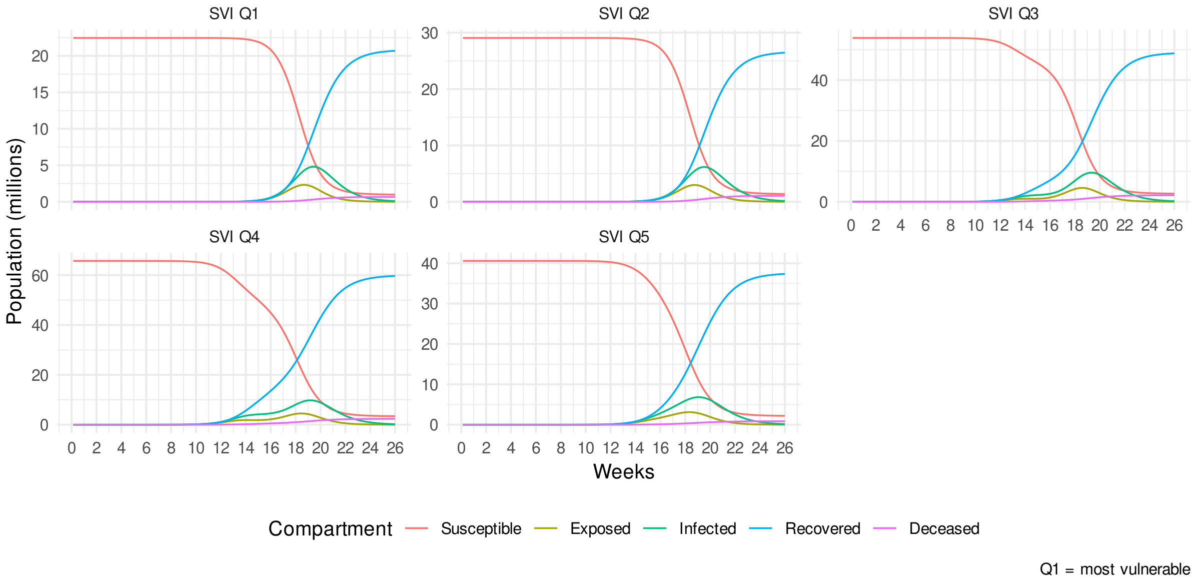


Fig. A.1. SIR dynamics for *Baseline* scenario with no interventions (Scenario 1, Table 3), stratified by SES quintile according to inverse SVI.

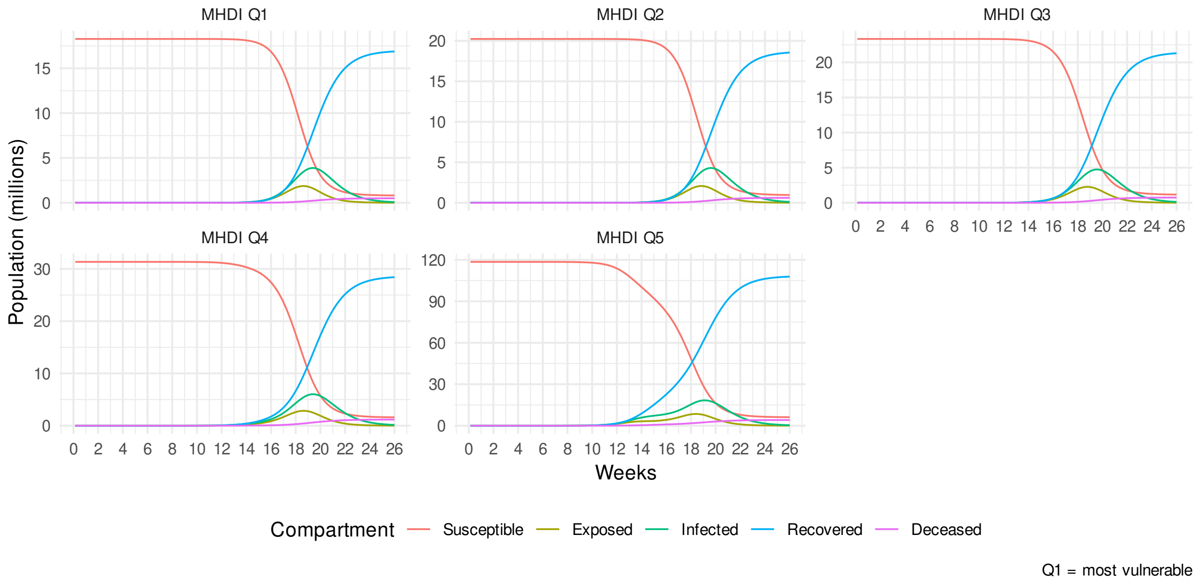


Fig. A.2. SIR dynamics for *Baseline* scenario with no interventions (Scenario 1, Table 3), stratified by SES quintile according to MHDI.

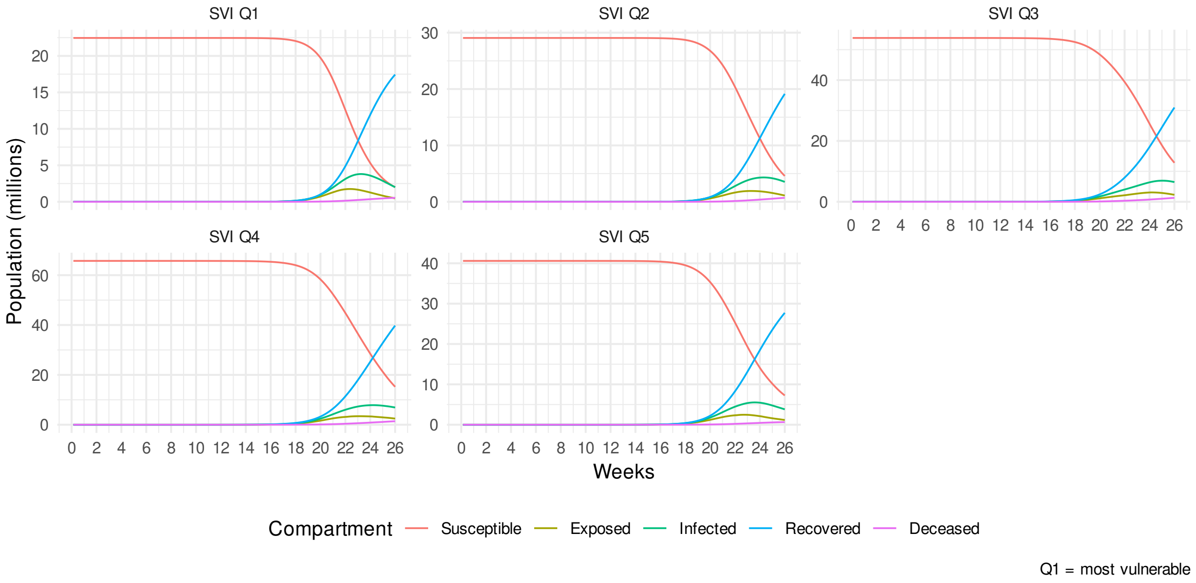


Fig. A.3. SIR dynamics for *NPIs* only scenario with uptake following real-world patterns (Scenario 2, Table 3), stratified by SES quintile according to inverse SVI.

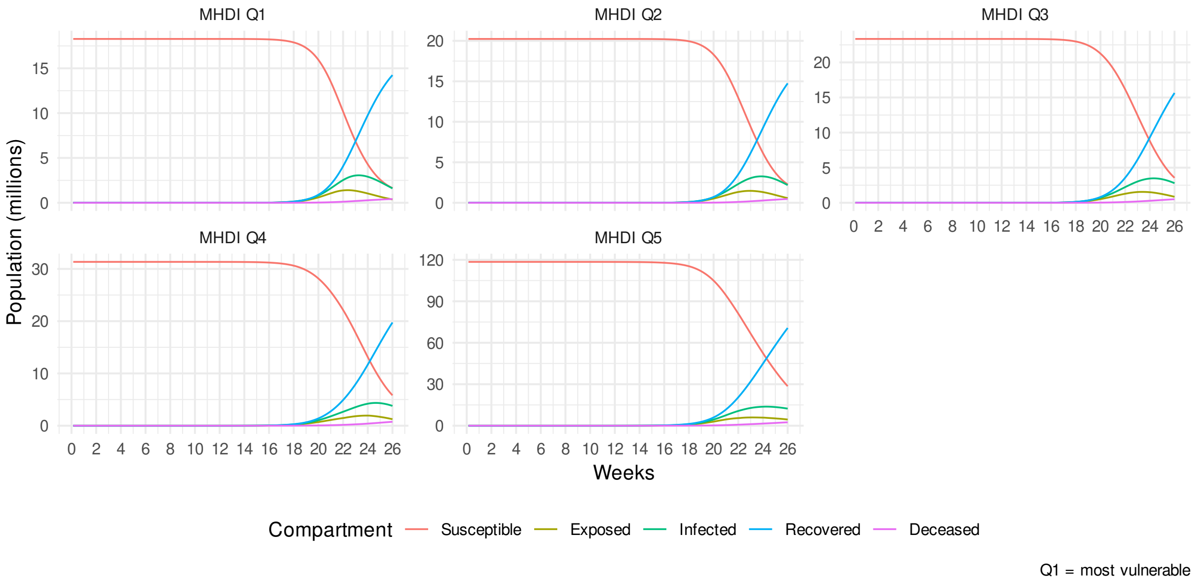


Fig. A.4. SIR dynamics for *NPIs* only scenario with uptake following real-world patterns (Scenario 2, Table 3), stratified by SES quintile according to MHDI.

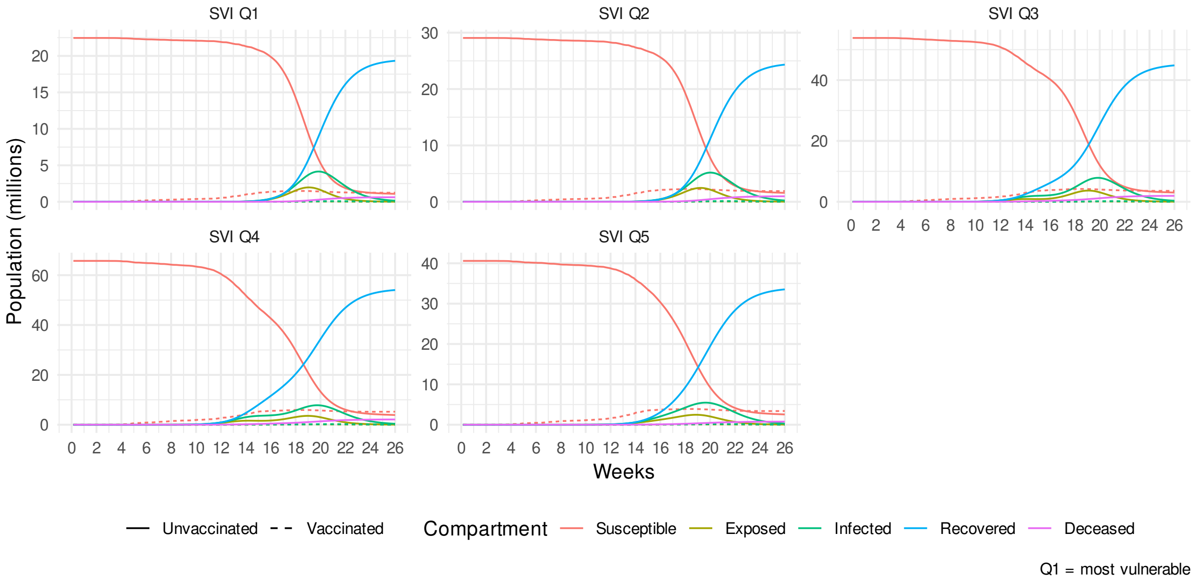


Fig. A.5. SIR dynamics for *Vaccination* only scenario with uptake following real-world patterns (Scenario 3, Table 3), stratified by SES quintile according to inverse SVI.

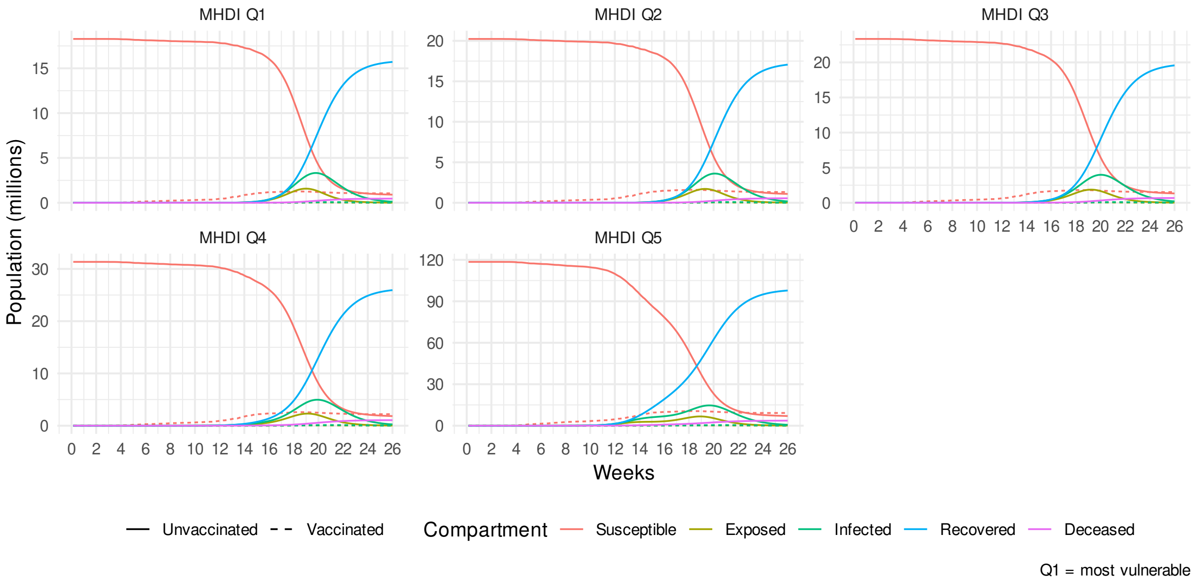


Fig. A.6. SIR dynamics for *Vaccination* only scenario with uptake following real-world patterns (Scenario 3, Table 3), stratified by SES quintile according to MHDI.

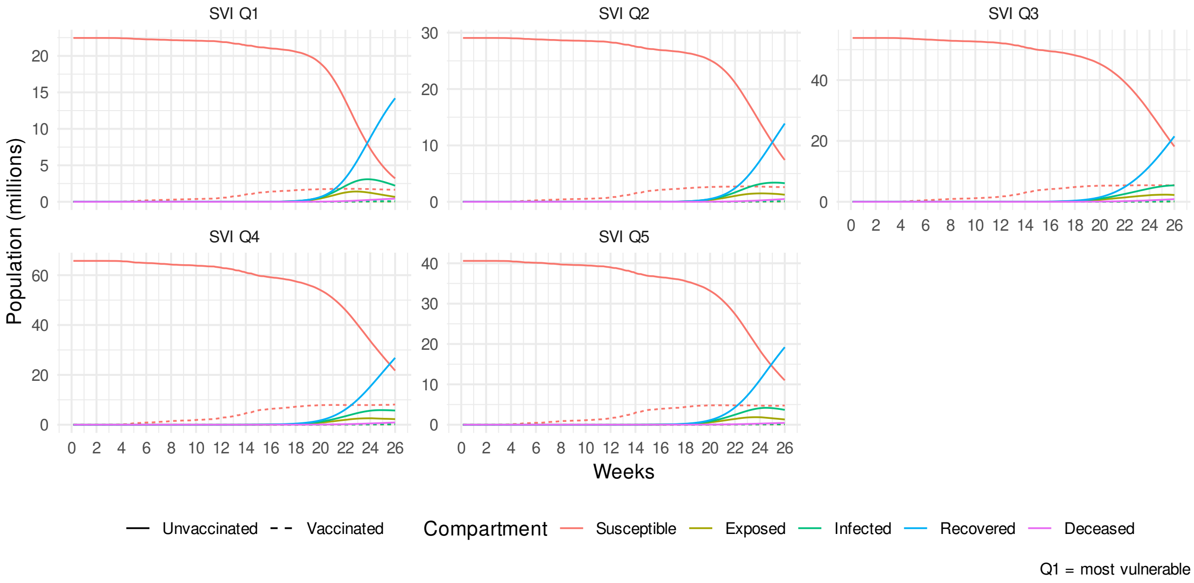


Fig. A.7. SIR dynamics for *NPIs + Vaccination* scenario with uptake following real-world patterns (Scenario 4, Table 3), stratified by SES quintile according to inverse SVI.

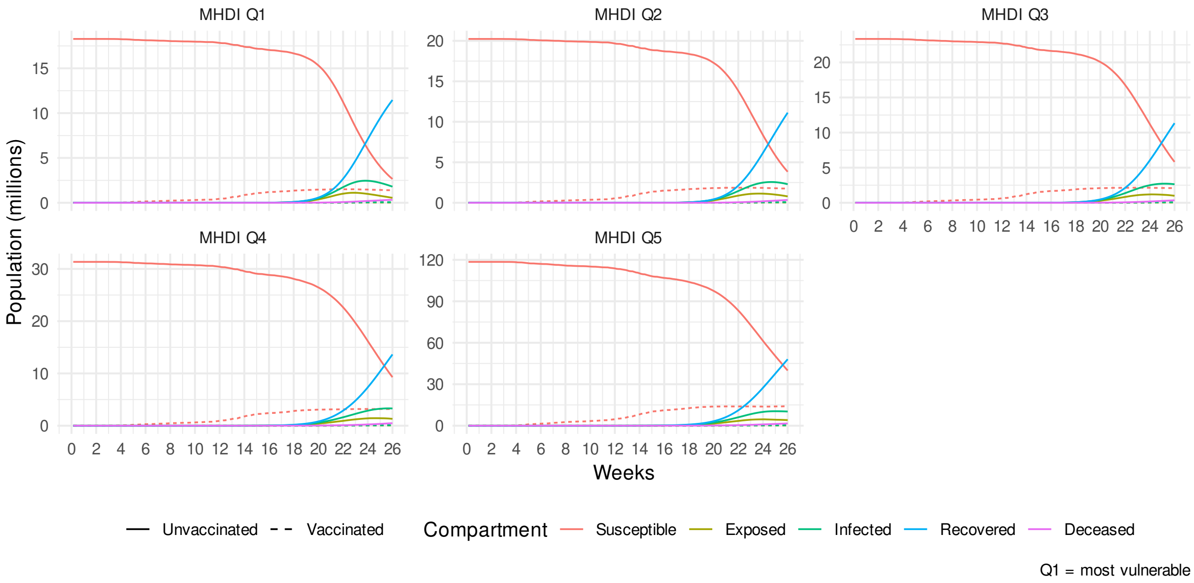


Fig. A.8. SIR dynamics for *NPIs + Vaccination* scenario with uptake following real-world patterns (Scenario 4, Table 3), stratified by SES quintile according to MHDI.

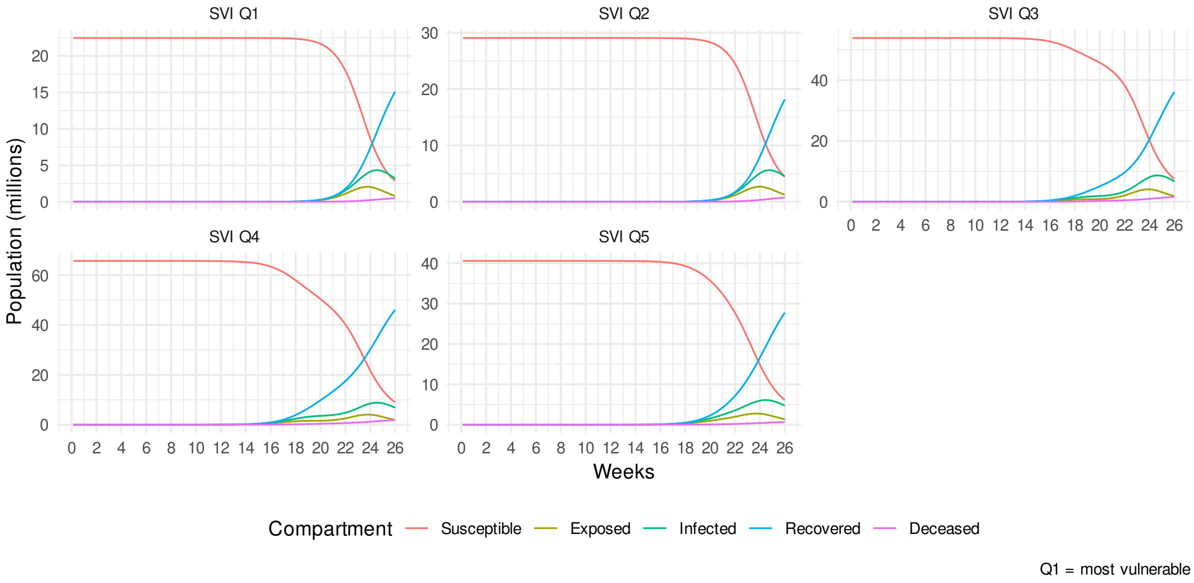


Fig. A.9. SIR dynamics for *NPIs-equal* scenario with all municipalities reducing their mobility by the same amount and no vaccination (Scenario 5, Table 3), stratified by SES quintile according to inverse SVI.

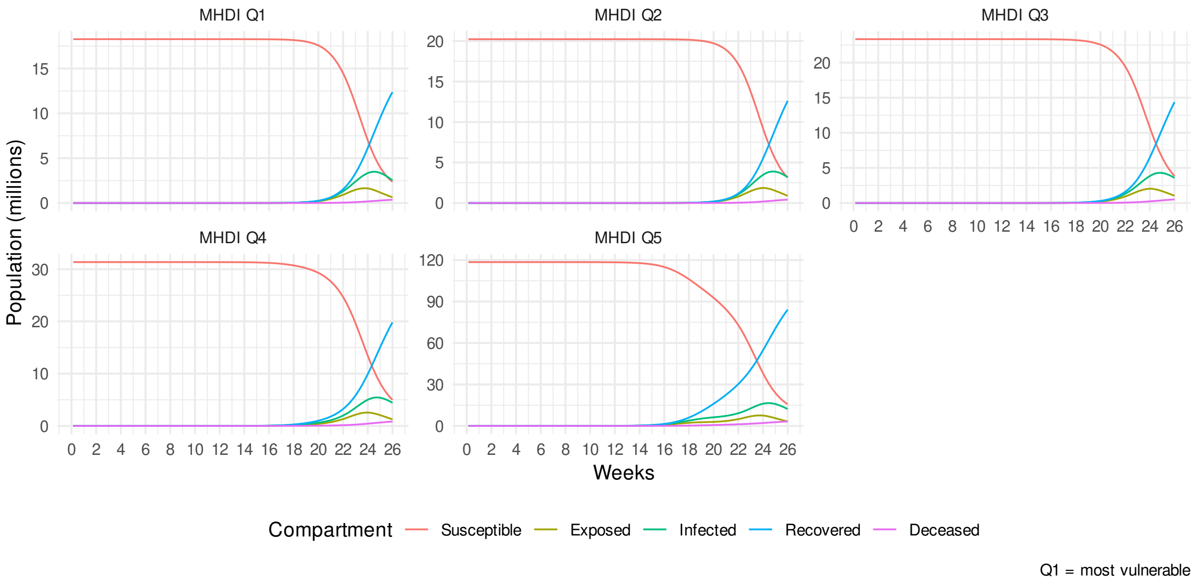


Fig. A.10. SIR dynamics for *NPIs-equal* scenario with all municipalities reducing their mobility by the same amount and no vaccination (Scenario 5, Table 3), stratified by SES quintile according to MHDI.

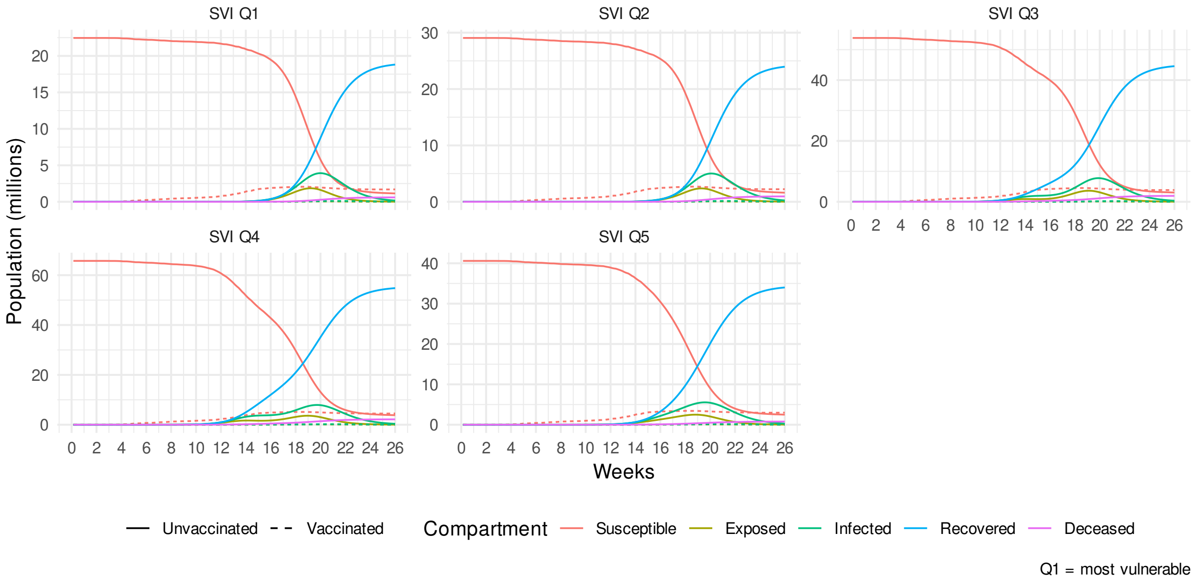


Fig. A.11. SIR dynamics for *Vaccination-equal* scenario with all municipalities vaccinating at the same rate and no NPIs (Scenario 6, Table 3), stratified by SES quintile according to inverse SVI.

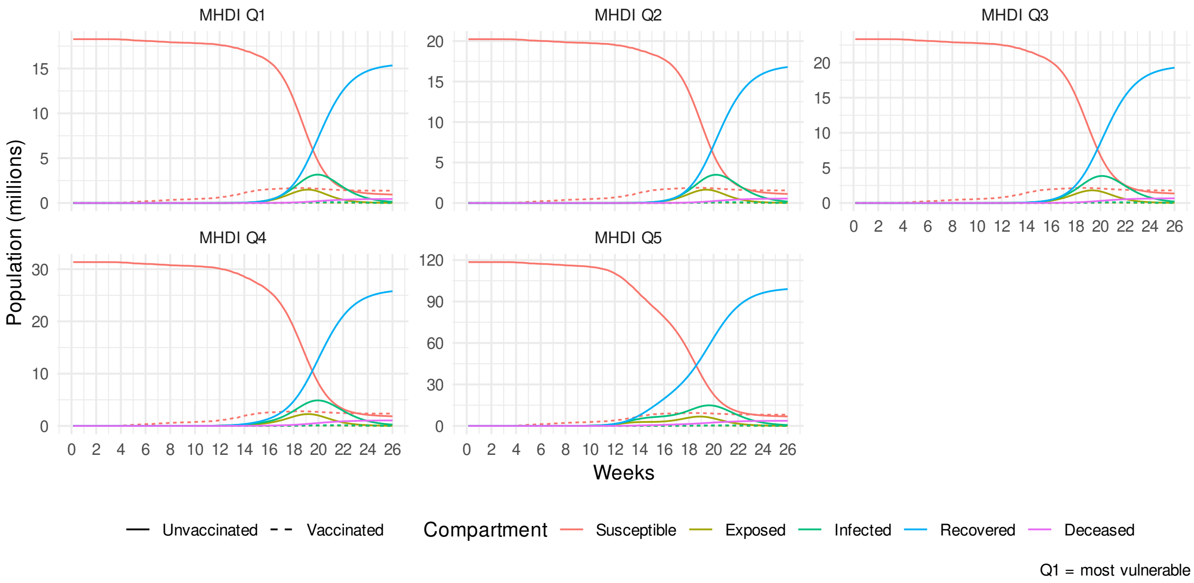


Fig. A.12. SIR dynamics for *Vaccination-equal* scenario with all municipalities vaccinating at the same rate and no NPIs (Scenario 6, Table 3), stratified by SES quintile according to MHDI.

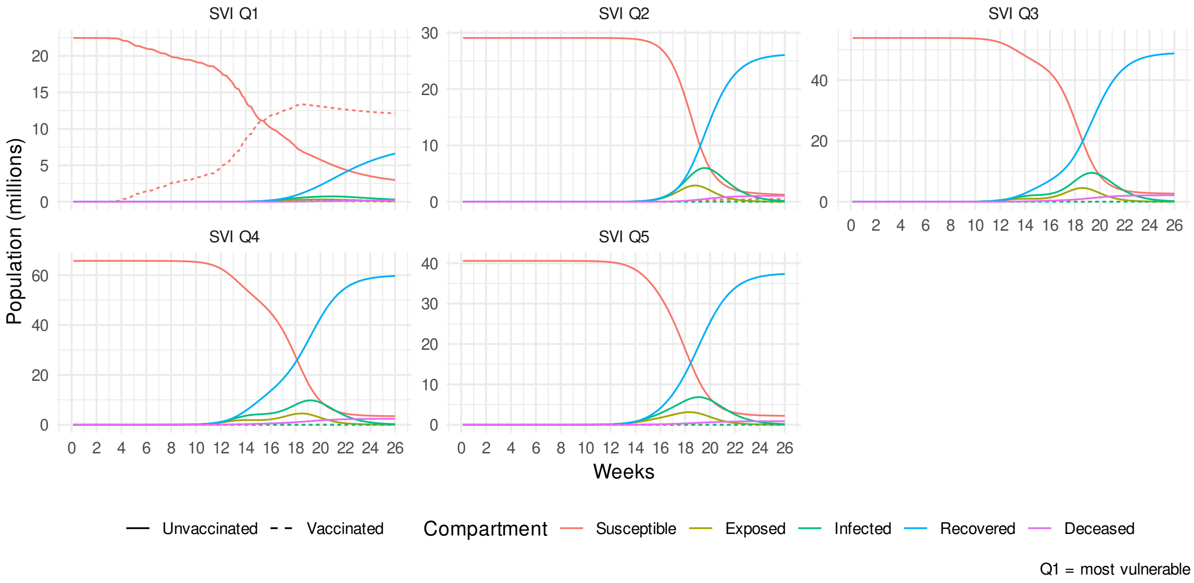


Fig. A.13. SIR dynamics for *Vaccination-equitable* scenario with municipalities fully vaccinated in descending order according to vulnerability and no NPIs (Scenario 7, Table 3), stratified by SES quintile according to inverse SVI.

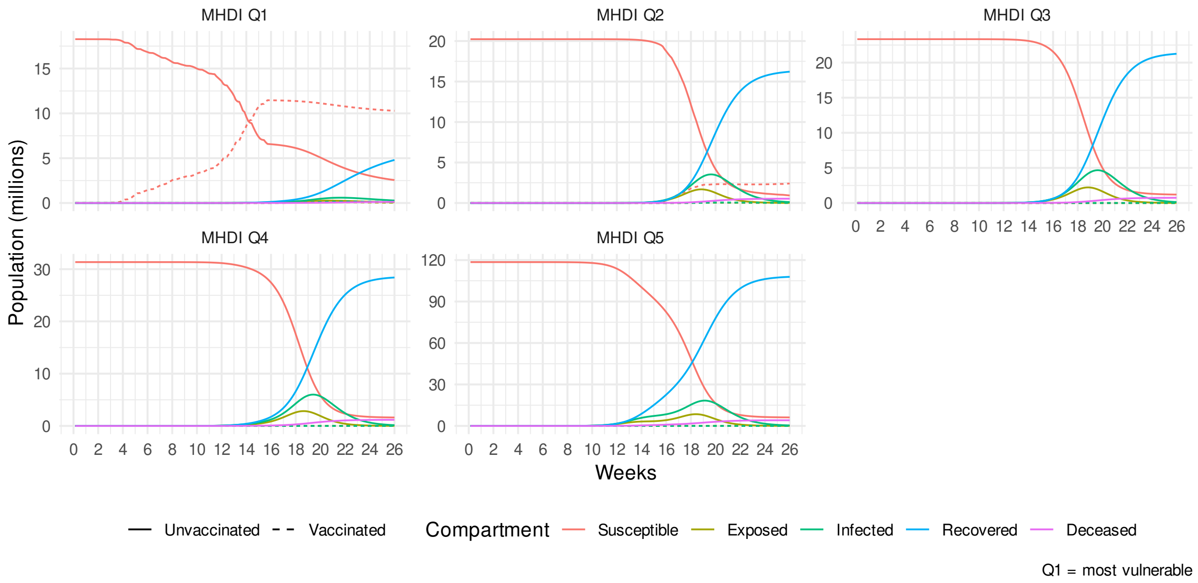


Fig. A.14. SIR dynamics for *Vaccination-equitable* scenario with municipalities fully vaccinated in descending order according to vulnerability and no NPIs (Scenario 7, Table 3), stratified by SES quintile according to MHDI.

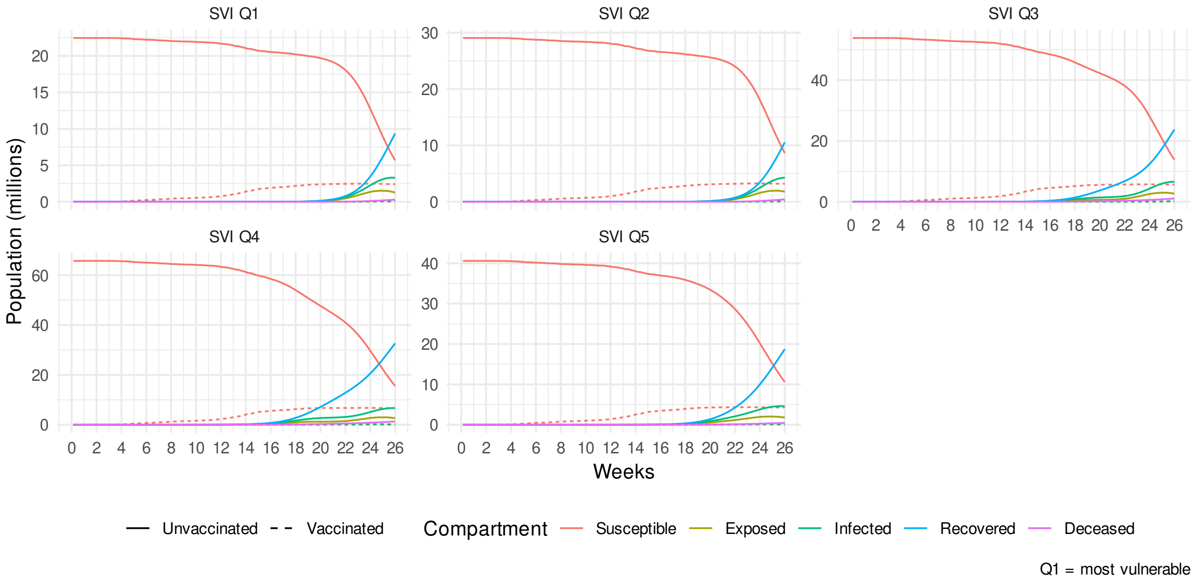


Fig. A.15. SIR dynamics for *NPIs + Vaccination-equal* scenario with all municipalities reducing their mobility by the same amount and vaccinating at the same rate (Scenario 8, Table 3), stratified by SES quintile according to inverse SVI.

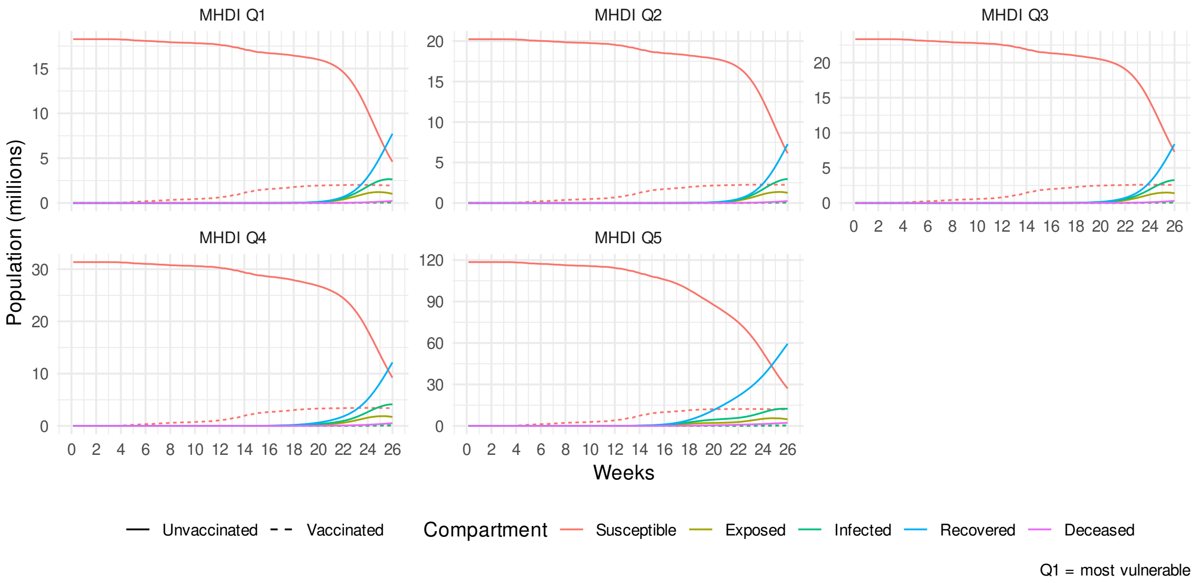


Fig. A.16. SIR dynamics for *NPIs + Vaccination-equal* scenario with all municipalities reducing their mobility by the same amount and vaccinating at the same rate (Scenario 8, Table 3), stratified by SES quintile according to MHDI.

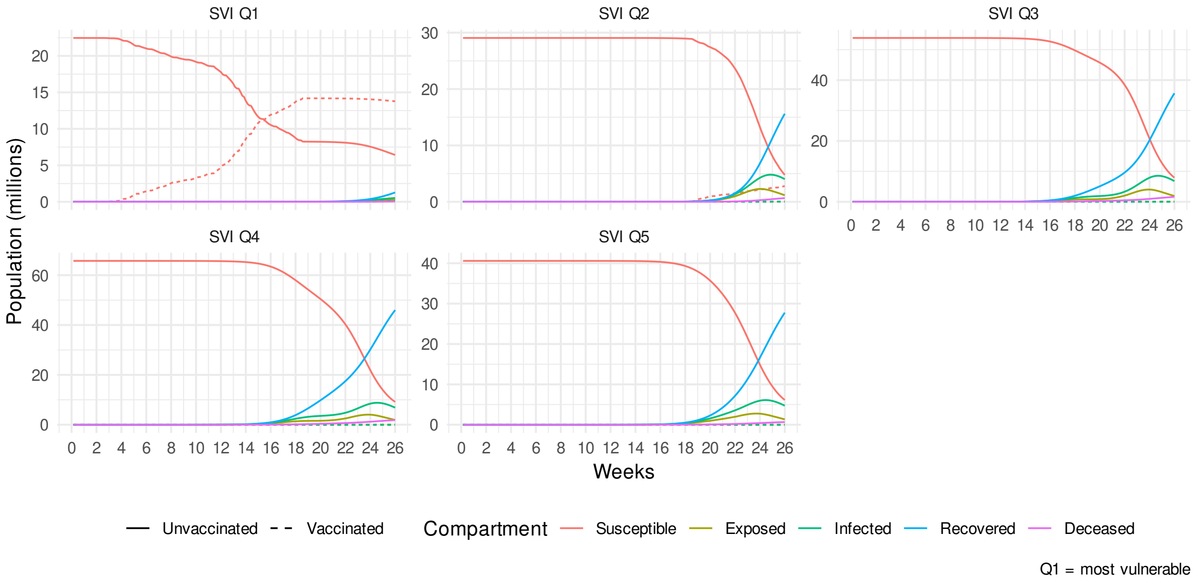


Fig. A.17. SIR dynamics for *NPIs-equal + Vaccination-equitable* scenario with all municipalities reducing their mobility by the same amount and fully vaccinating in descending order according to vulnerability (Scenario 9, Table 3), stratified by SES quintile according to inverse SVI.

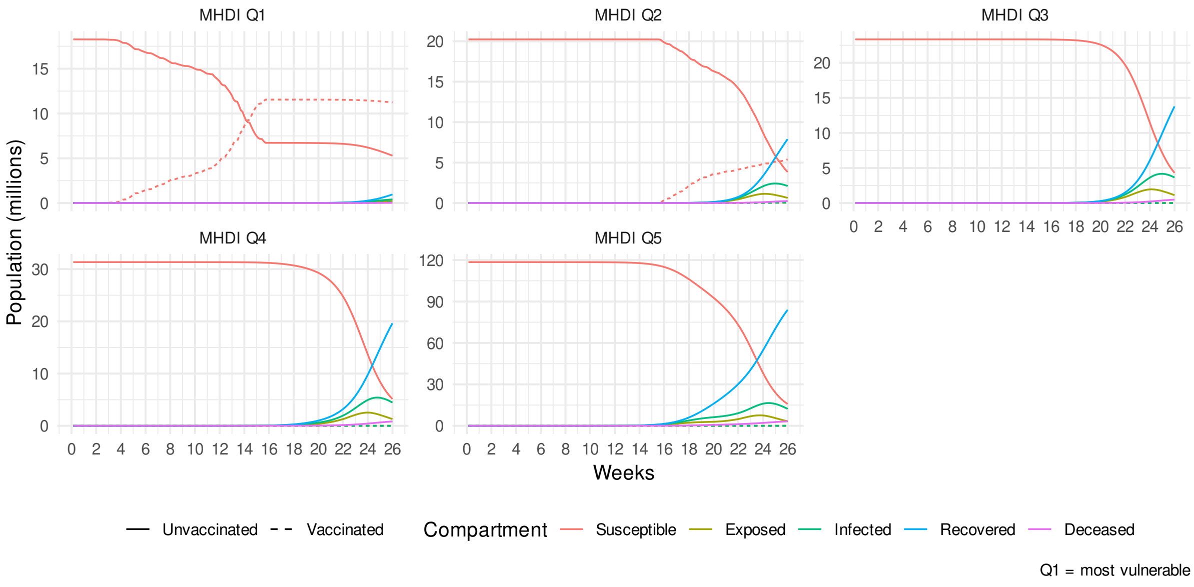


Fig. A.18. SIR dynamics for *NPIs-equal + Vaccination-equitable* scenario with all municipalities reducing their mobility by the same amount and fully vaccinating in descending order according to vulnerability (Scenario 9, Table 3), stratified by SES quintile according to MHDI.

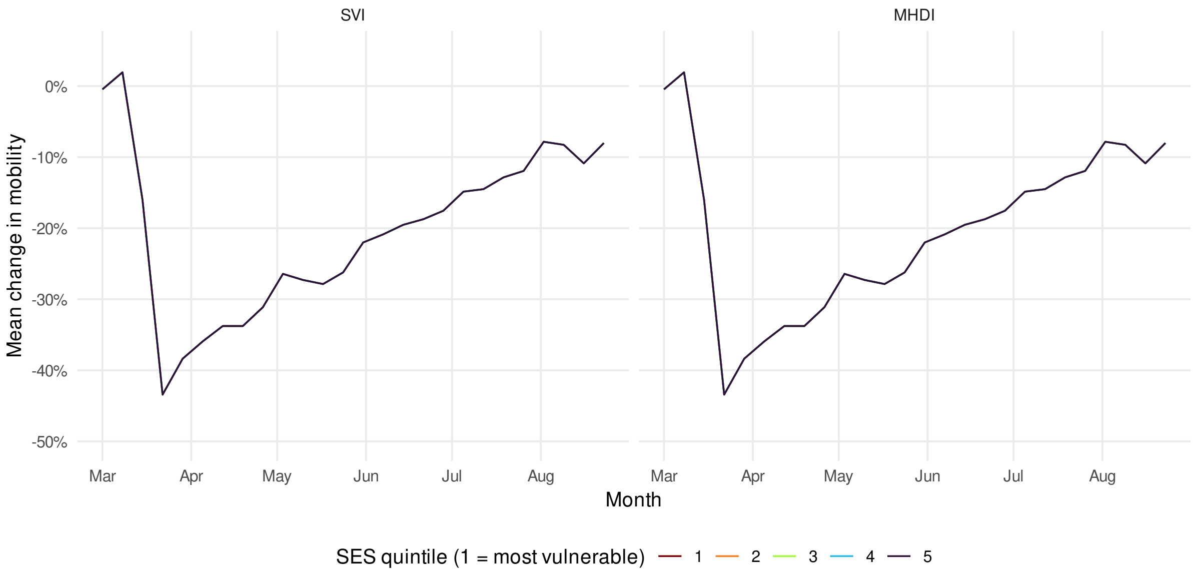


Fig. A.19. Proportional change in mobility over time following the onset of the COVID-19 pandemic by municipality SES quintile, by inverse SVI and MHDI under the counterfactual scenario in which all municipalities reduce their mobility by the same amount.

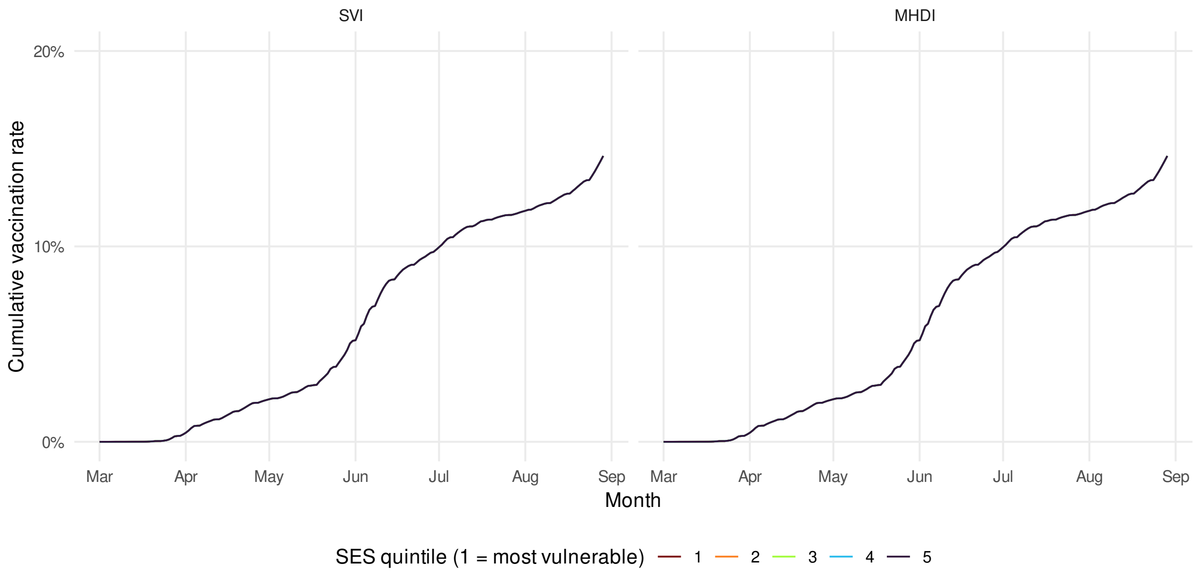


Fig. A.20. Cumulative COVID-19 vaccination uptake by municipality SES quintile, by inverse SVI and MHDI under the counterfactual scenario in which all municipalities have the same vaccination rate. Overall vaccination uptake is temporally transposed to align with a counterfactual vaccination campaign start date of March 1, 2020 instead of January 16, 2021.

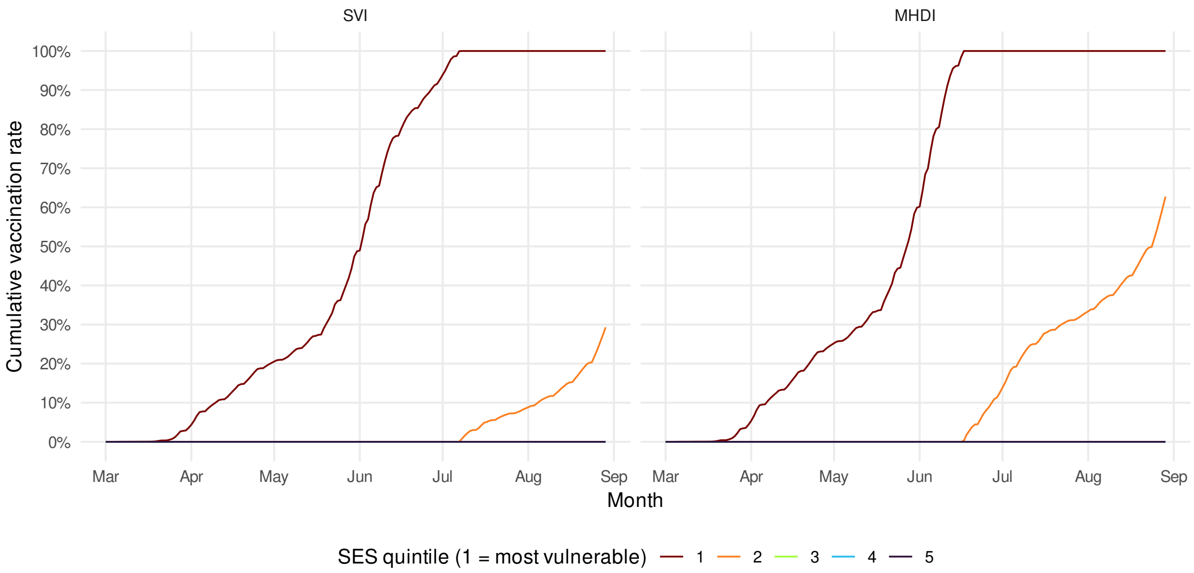


Fig. A.21. Cumulative COVID-19 vaccination uptake by municipality SES quintile, by inverse SVI and MHDI under the counterfactual scenario in which municipalities are fully vaccinated in descending order according to vulnerability. Overall vaccination uptake is temporally transposed to align with a counterfactual vaccination campaign start date of March 1, 2020 instead of January 16, 2021.

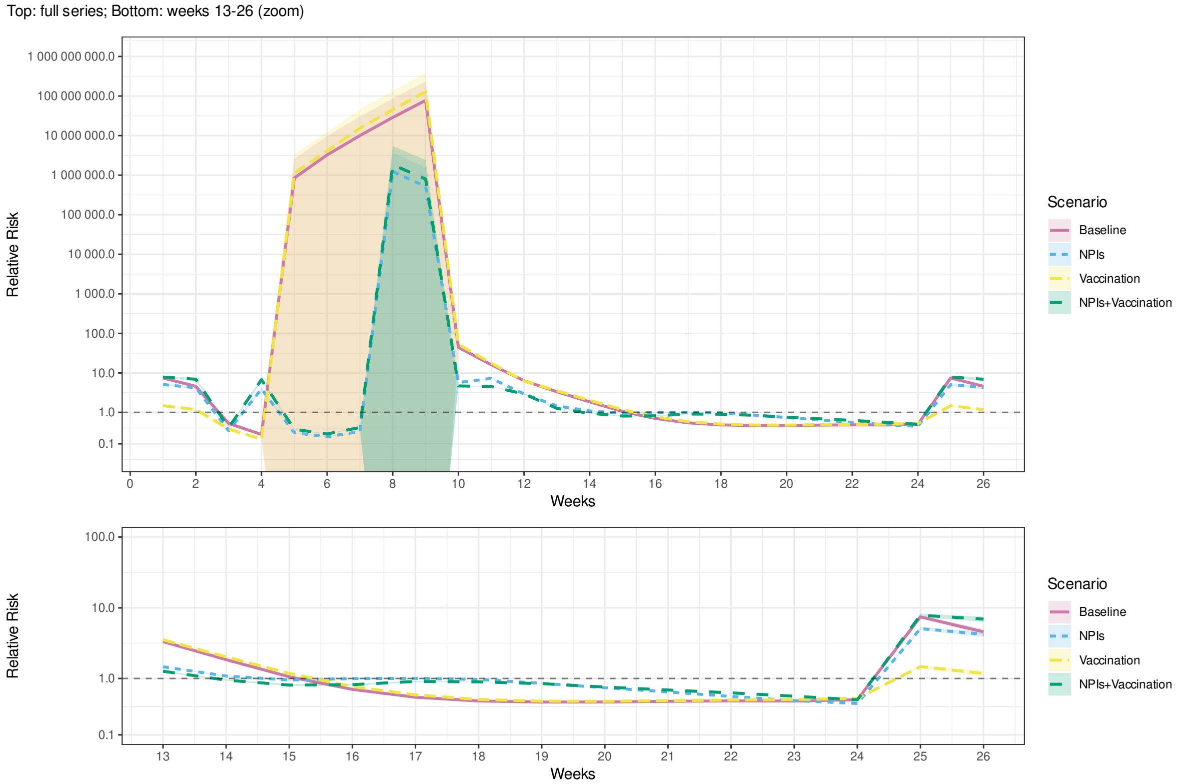


Fig. A.22. Relative risk of mortality in the least vulnerable (Q5) compared to the most vulnerable (Q1) municipalities by SVI during the first 26 weeks of an outbreak in the real world (*Empirical*) and simulations of scenarios 1–4 (Table 3). with infection fatality rate halved ($d_i$, Table 2). Shaded bands indicate 95% confidence intervals.

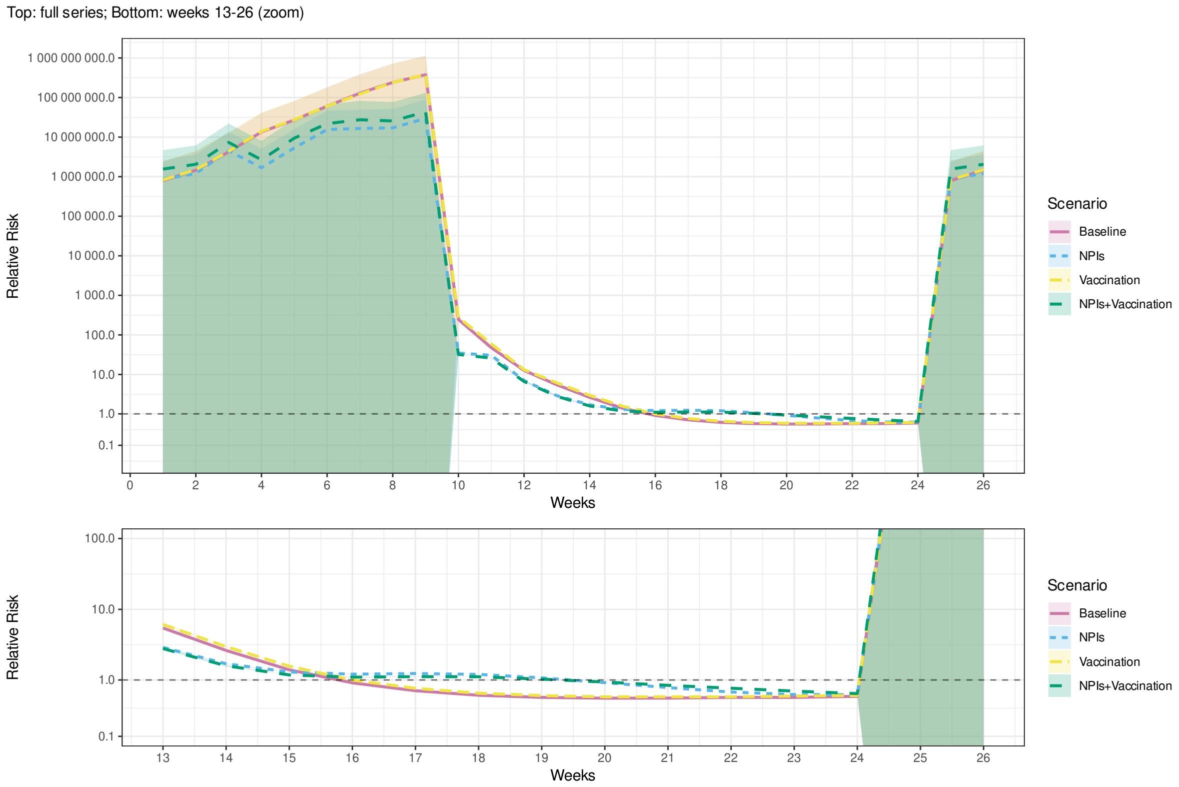


Fig. A.23. Relative risk of mortality in the least vulnerable (Q5) compared to the most vulnerable (Q1) municipalities by MHDI during the first 26 weeks of an outbreak in the real world (*Empirical*) and simulations of scenarios 1–4 (Table 3). with infection fatality rate halved ($d_i$, Table 2). Shaded bands indicate 95% confidence intervals.

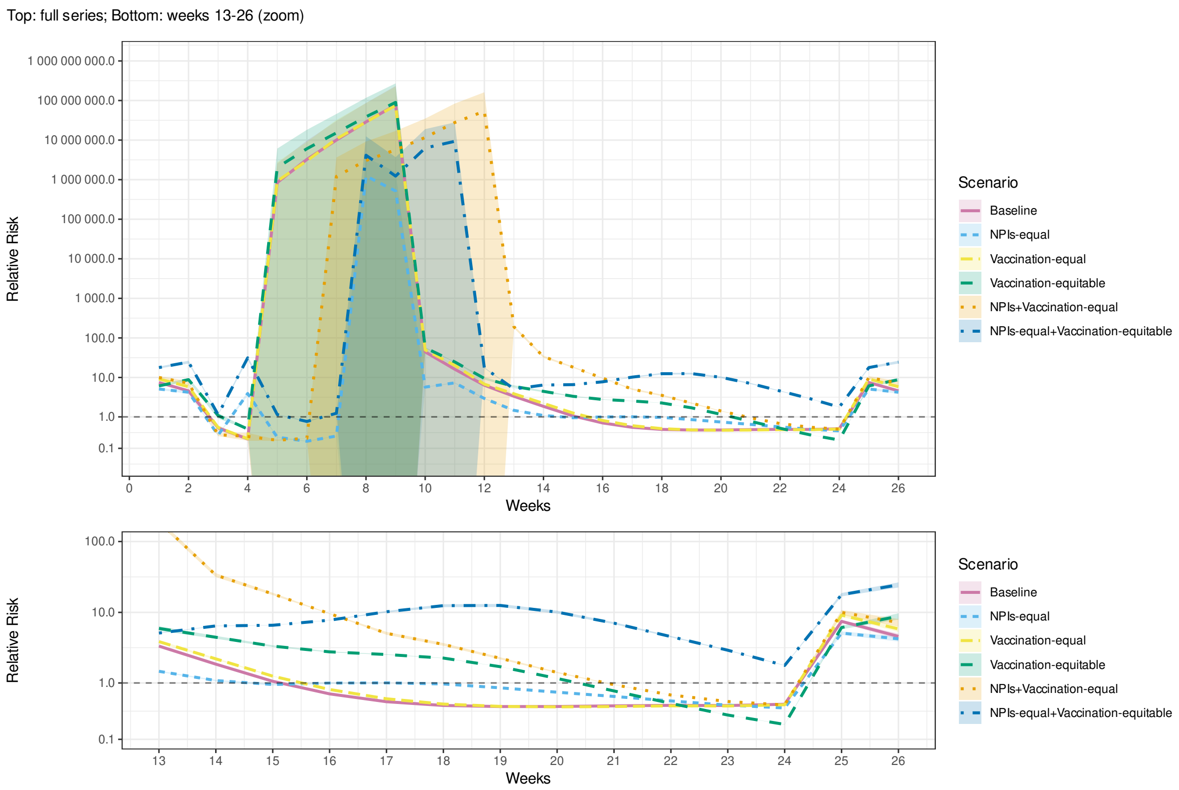


Fig. A.24. Relative risk of mortality in the least vulnerable (Q5) compared to the most vulnerable (Q1) municipalities by SVI during the first 26 weeks of an outbreak in simulations of scenarios 1 and 5–9 (Table 3). with infection fatality rate halved ($d_i$, Table 2). Shaded bands indicate 95% confidence intervals.

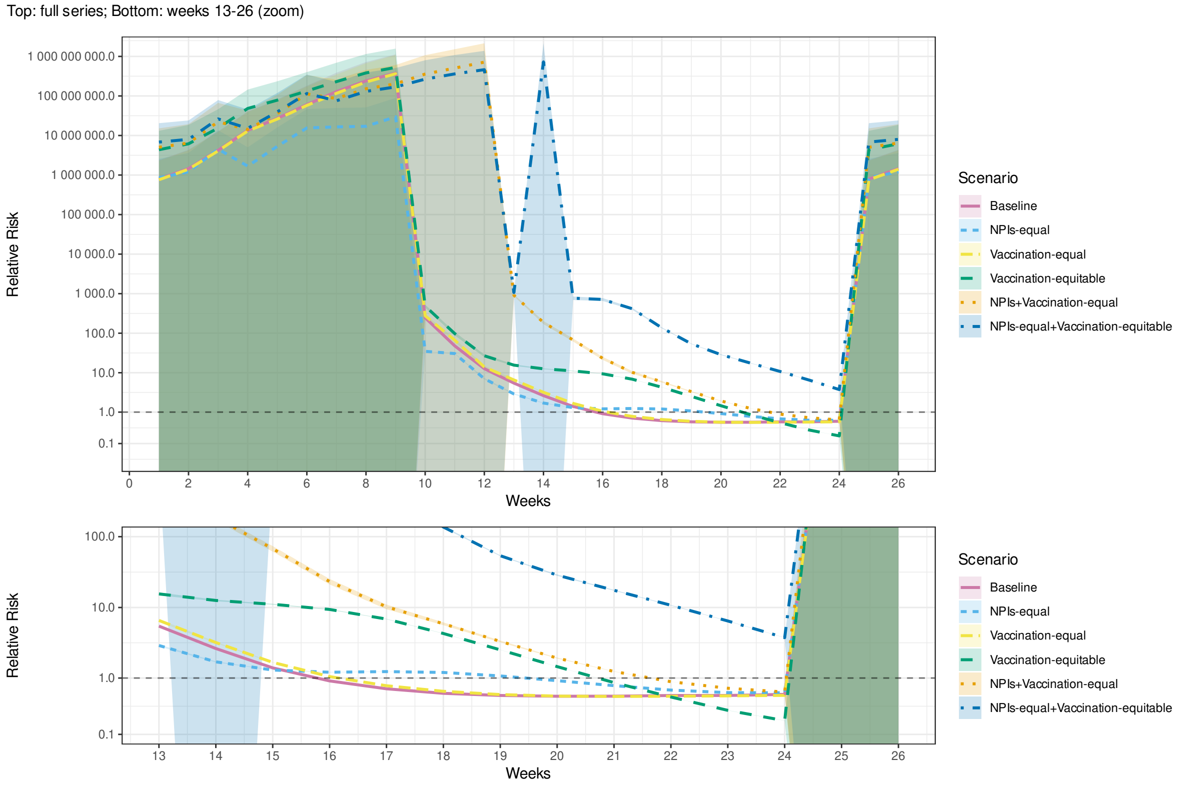


Fig. A.25. Relative risk of mortality in the least vulnerable (Q5) compared to the most vulnerable (Q1) municipalities by MHDI during the first 26 weeks of an outbreak in simulations of scenarios 1 and 5–9 (Table 3). with infection fatality rate halved ($d_i$, Table 2). Shaded bands indicate 95% confidence intervals.

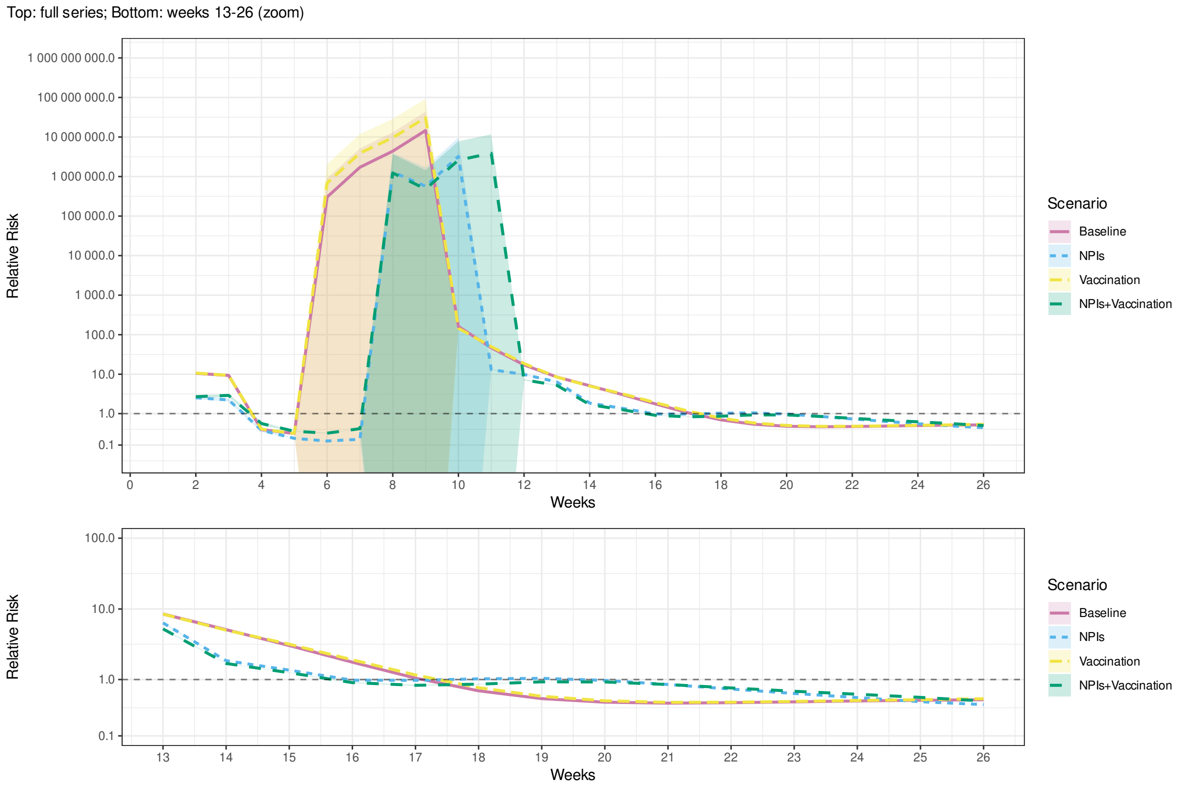


Fig. A.26. Relative risk of mortality in the least vulnerable (Q5) compared to the most vulnerable (Q1) municipalities by SVI during the first 26 weeks of an outbreak in the real world (*Empirical*) and simulations of scenarios 1–4 (Table 3). with infection fatality rate doubled ($d_i$, Table 2). Shaded bands indicate 95% confidence intervals.

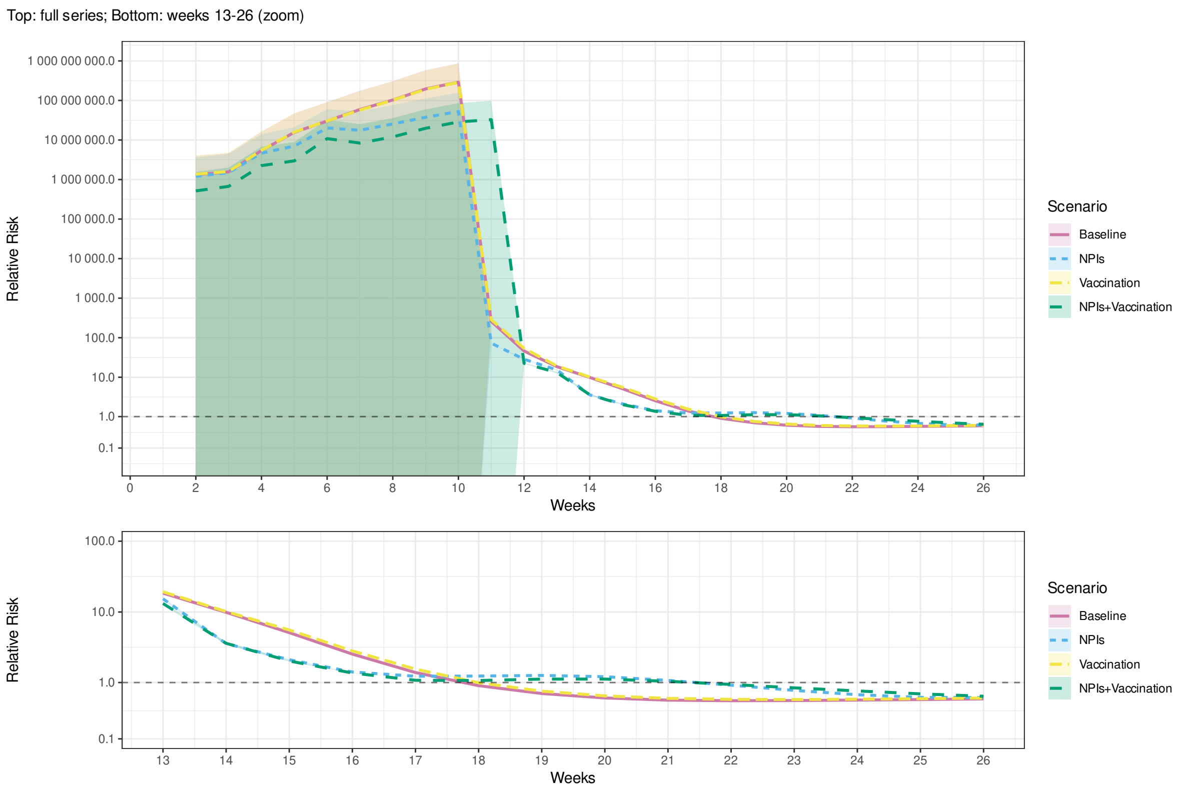


Fig. A.27. Relative risk of mortality in the least vulnerable (Q5) compared to the most vulnerable (Q1) municipalities by MHDI during the first 26 weeks of an outbreak in the real world (*Empirical*) and simulations of scenarios 1–4 (Table 3). with infection fatality rate doubled ($d_i$, Table 2). Shaded bands indicate 95% confidence intervals.

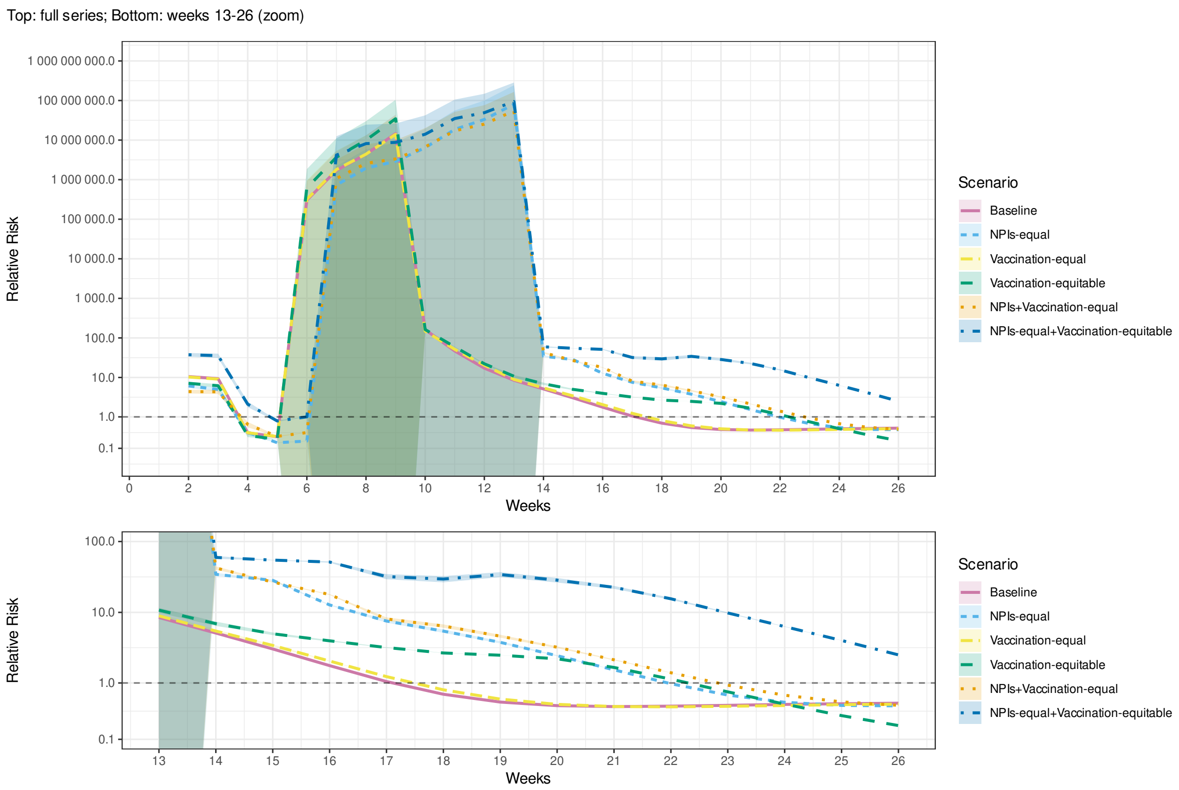


Fig. A.28. Relative risk of mortality in the least vulnerable (Q5) compared to the most vulnerable (Q1) municipalities by SVI during the first 26 weeks of an outbreak in simulations of scenarios 1 and 5–9 (Table 3). with infection fatality rate doubled ($d_i$, Table 2). Shaded bands indicate 95% confidence intervals.

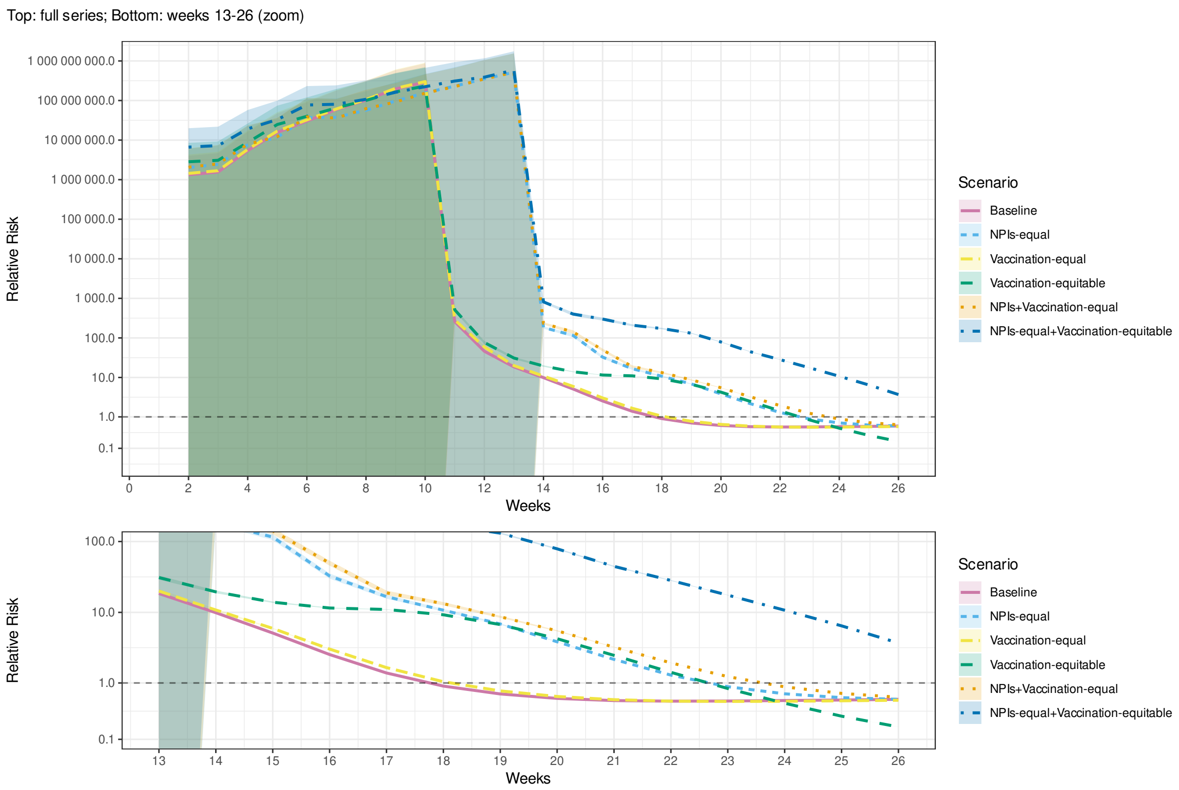


Fig. A.29. Relative risk of mortality in the least vulnerable (Q5) compared to the most vulnerable (Q1) municipalities by MHDI during the first 26 weeks of an outbreak in simulations of scenarios 1 and 5–9 (Table 3). with infection fatality rate doubled ($d_i$, Table 2). Shaded bands indicate 95% confidence intervals.

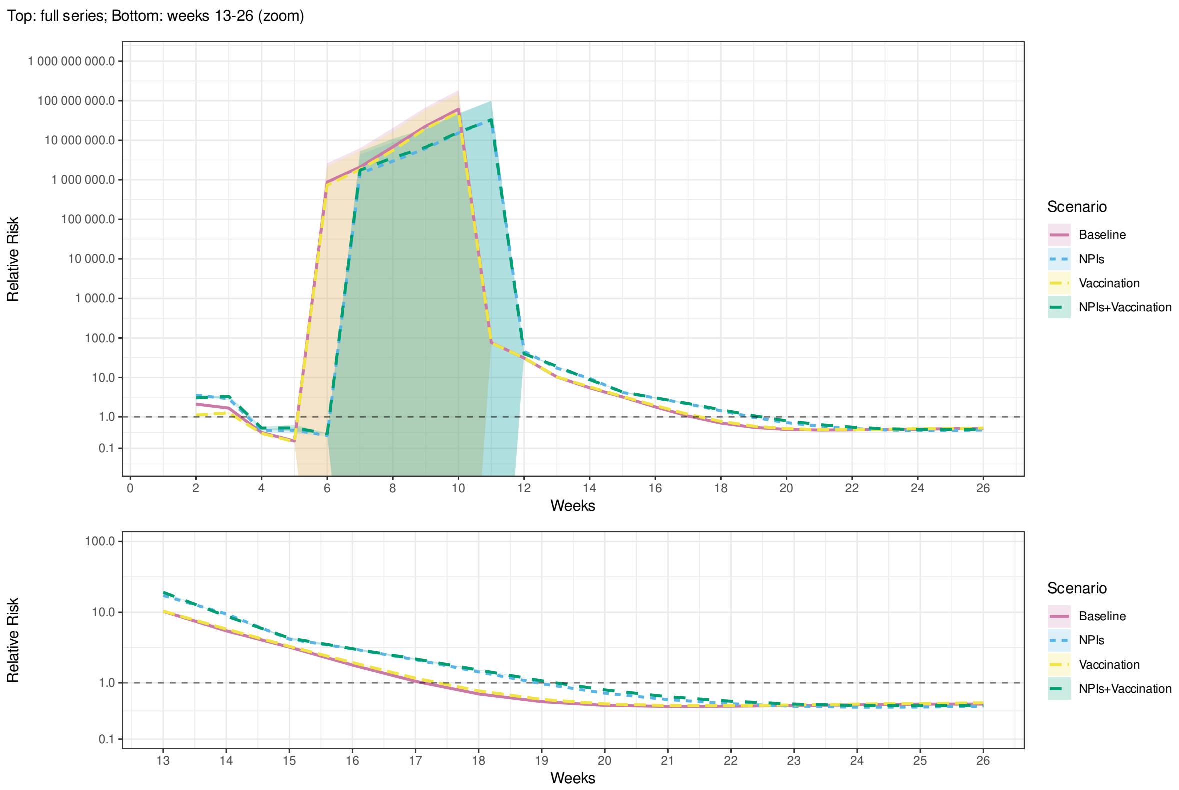


Fig. A.30. Relative risk of mortality in the least vulnerable (Q5) compared to the most vulnerable (Q1) municipalities by SVI during the first 26 weeks of an outbreak in the real world (*Empirical*) and simulations of scenarios 1–4 (Table 3). with effect of mobility changes on transmission halved ($\Delta M_i(t)$, Table 2). Shaded bands indicate 95% confidence intervals.

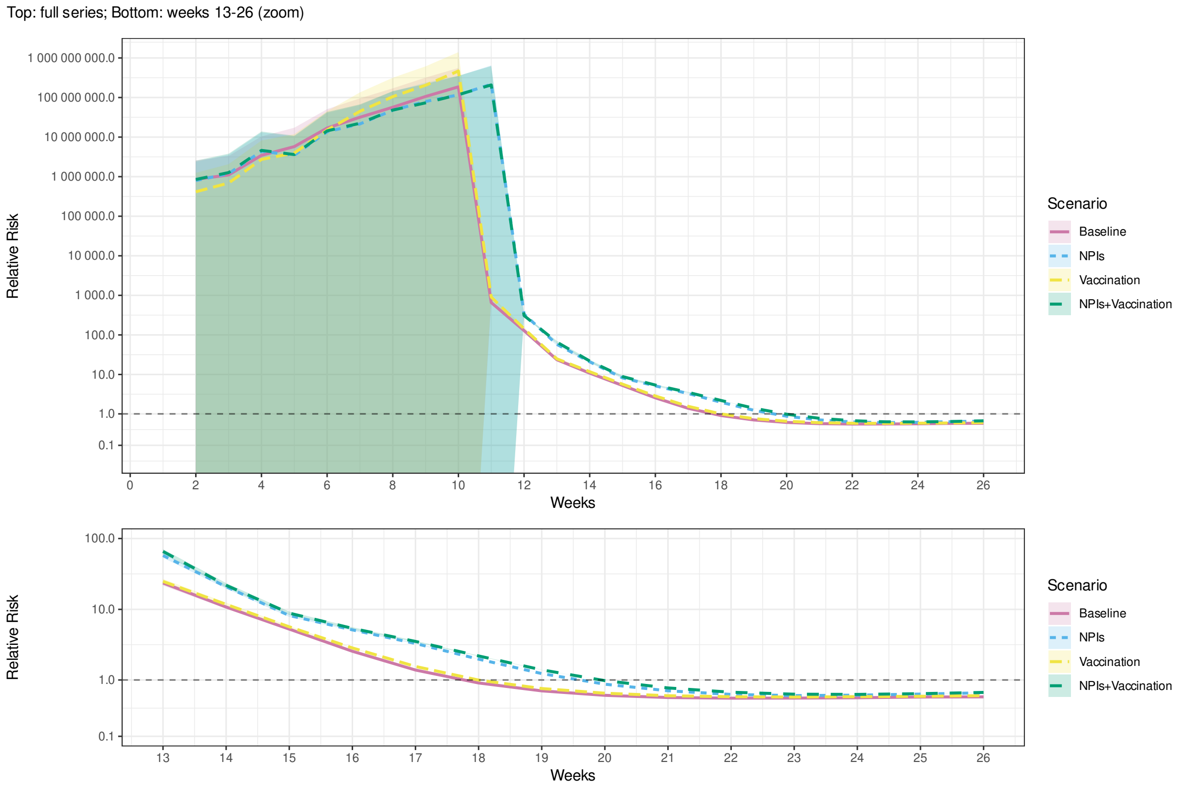


Fig. A.31. Relative risk of mortality in the least vulnerable (Q5) compared to the most vulnerable (Q1) municipalities by MHDI during the first 26 weeks of an outbreak in the real world (*Empirical*) and simulations of scenarios 1–4 (Table 3). with effect of mobility changes on transmission halved ($\Delta M_i(t)$, Table 2). Shaded bands indicate 95% confidence intervals.

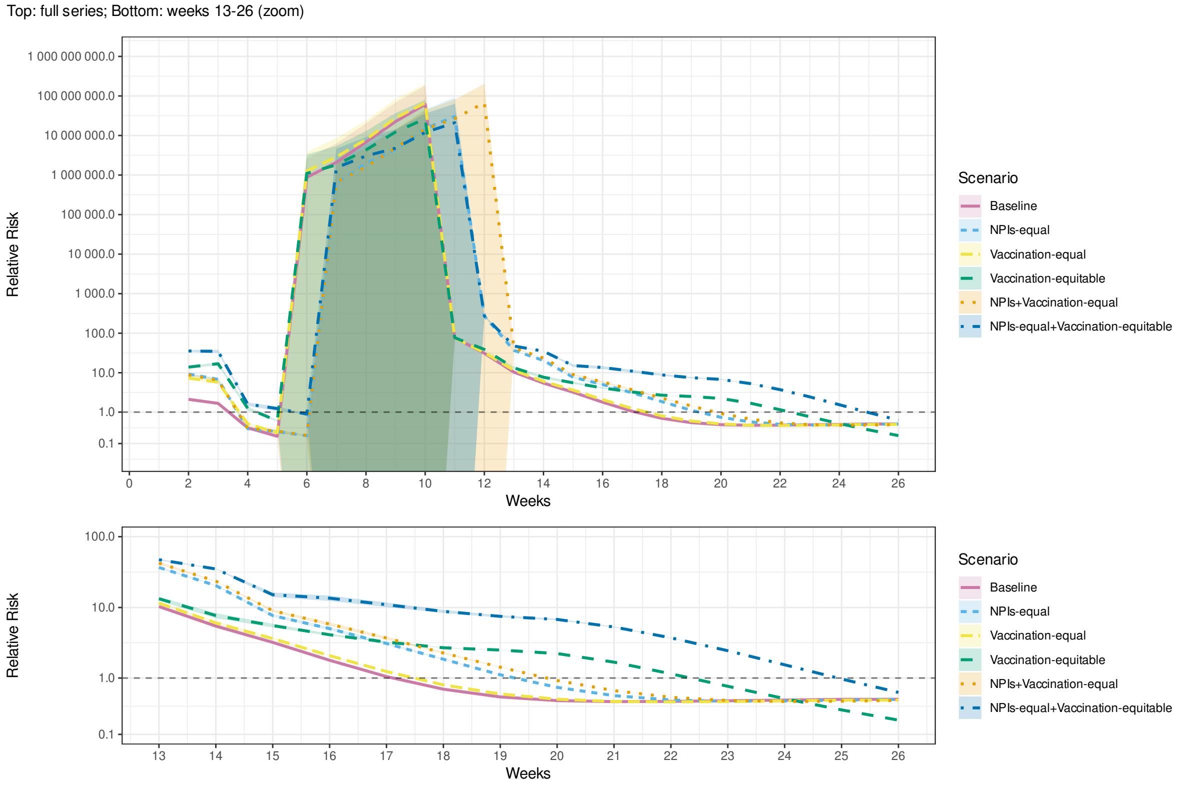


Fig. A.32. Relative risk of mortality in the least vulnerable (Q5) compared to the most vulnerable (Q1) municipalities by SVI during the first 26 weeks of an outbreak in simulations of scenarios 1 and 5–9 (Table 3). with effect of mobility changes on transmission halved ($\Delta M_i(t)$, Table 2). Shaded bands indicate 95% confidence intervals.

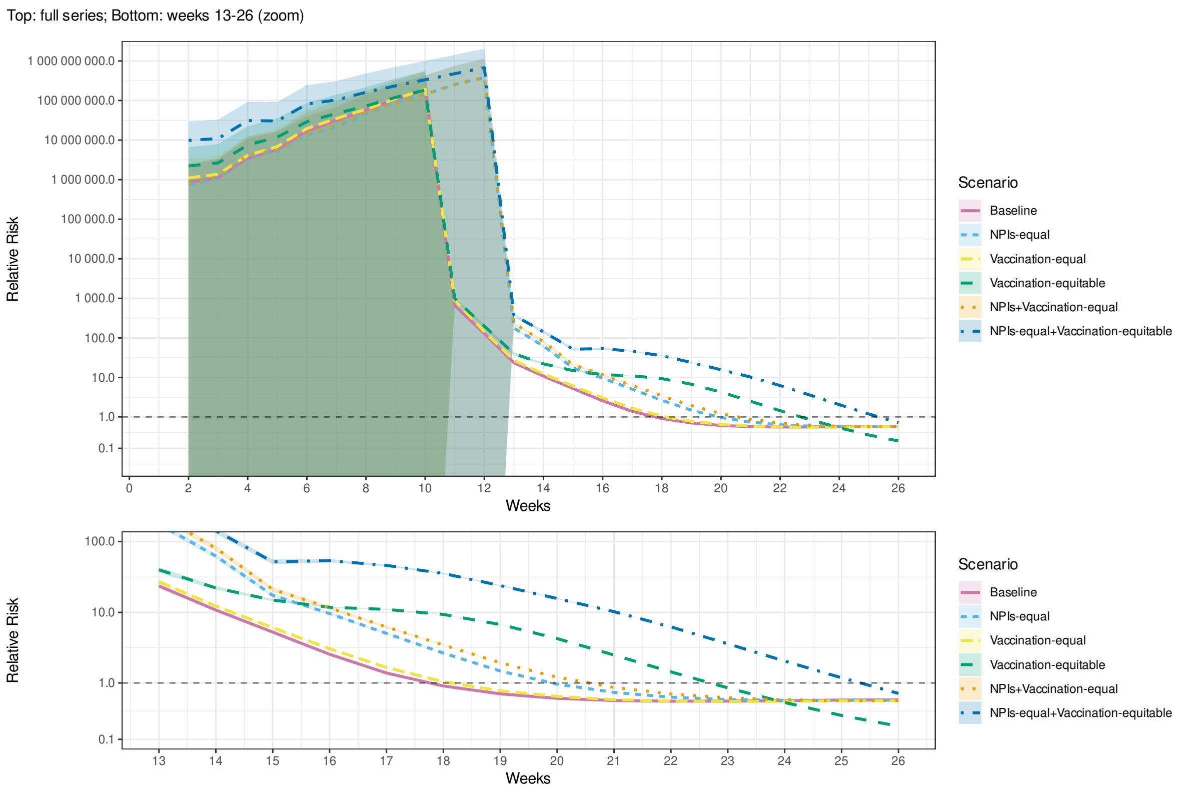


Fig. A.33. Relative risk of mortality in the least vulnerable (Q5) compared to the most vulnerable (Q1) municipalities by MHDI during the first 26 weeks of an outbreak in simulations of scenarios 1 and 5–9 (Table 3). with effect of mobility changes on transmission halved ($\Delta M_i(t)$, Table 2). Shaded bands indicate 95% confidence intervals.

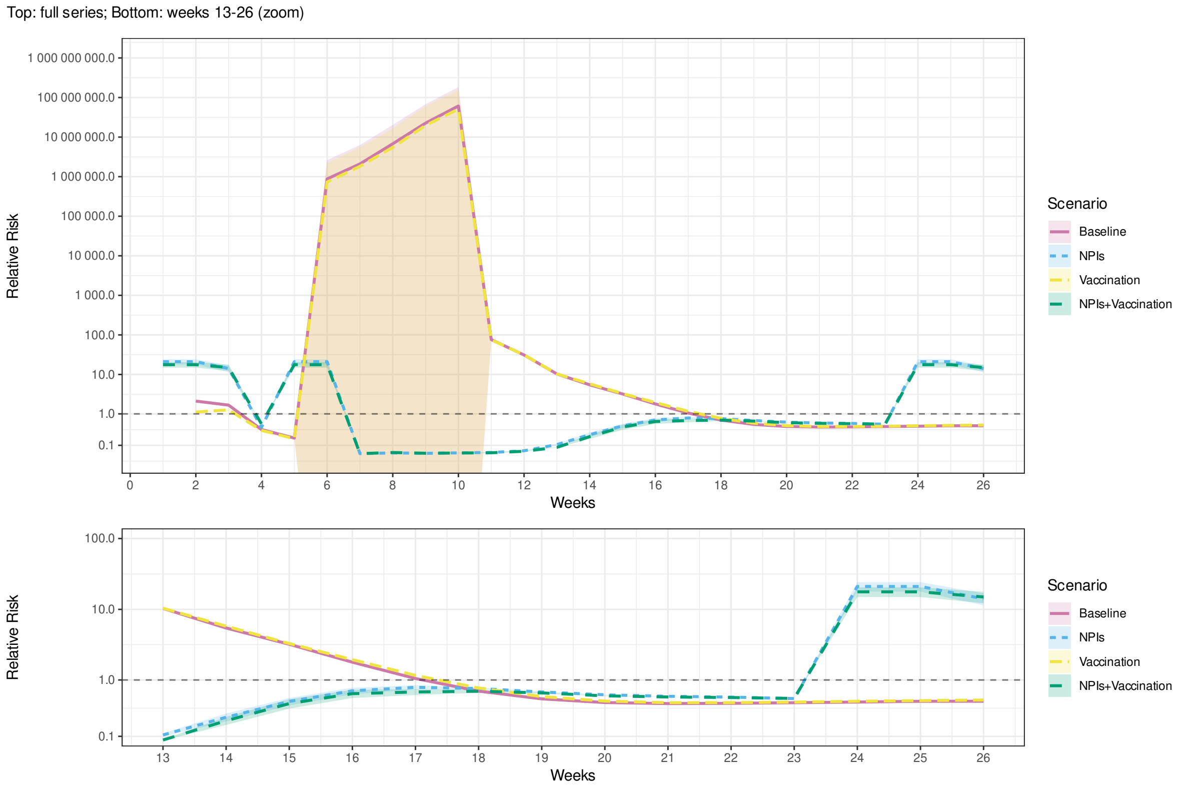


Fig. A.34. Relative risk of mortality in the least vulnerable (Q5) compared to the most vulnerable (Q1) municipalities by SVI during the first 26 weeks of an outbreak in the real world (*Empirical*) and simulations of scenarios 1–4 (Table 3). with effect of mobility changes on transmission doubled ($\Delta M_i(t)$, Table 2). Shaded bands indicate 95% confidence intervals.

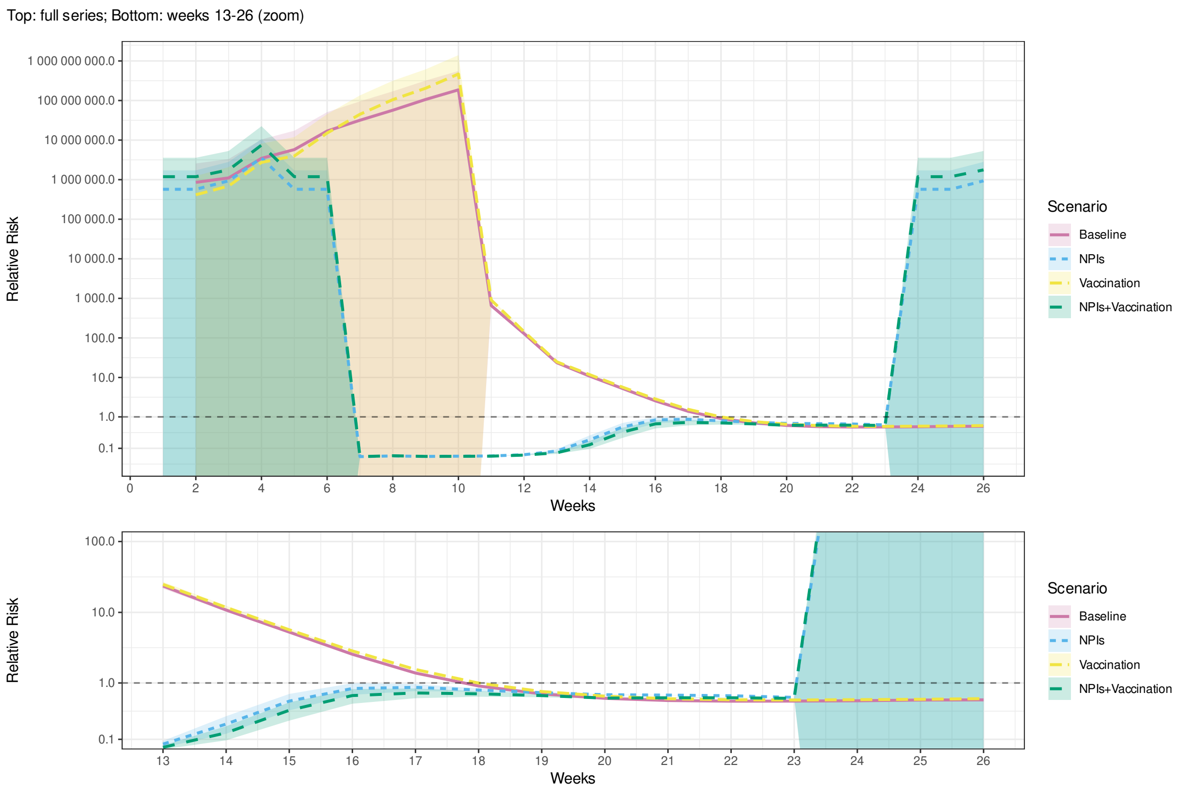


Fig. A.35. Relative risk of mortality in the least vulnerable (Q5) compared to the most vulnerable (Q1) municipalities by MHDI during the first 26 weeks of an outbreak in the real world (*Empirical*) and simulations of scenarios 1–4 (Table 3). with effect of mobility changes on transmission doubled ($\Delta M_i(t)$, Table 2). Shaded bands indicate 95% confidence intervals.

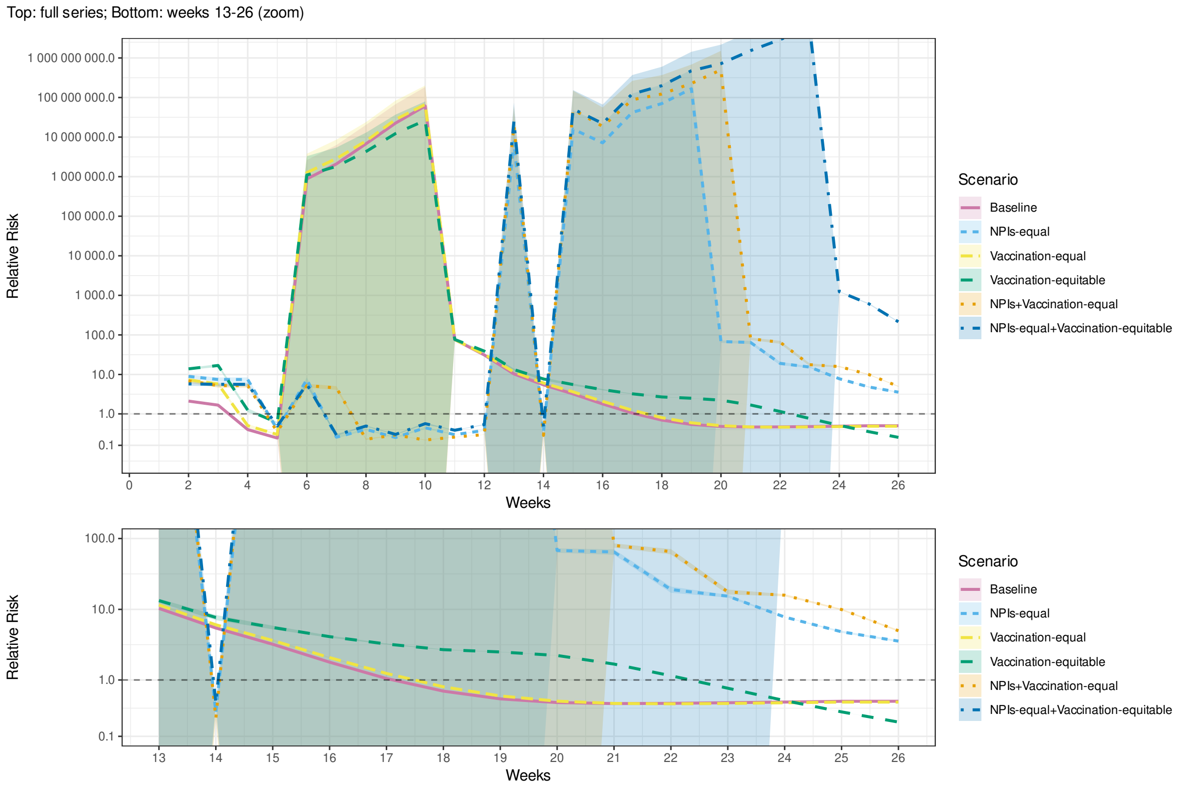


Fig. A.36. Relative risk of mortality in the least vulnerable (Q5) compared to the most vulnerable (Q1) municipalities by SVI during the first 26 weeks of an outbreak in simulations of scenarios 1 and 5–9 (Table 3). with effect of mobility changes on transmission doubled ($\Delta M_i(t)$, Table 2). Shaded bands indicate 95% confidence intervals.

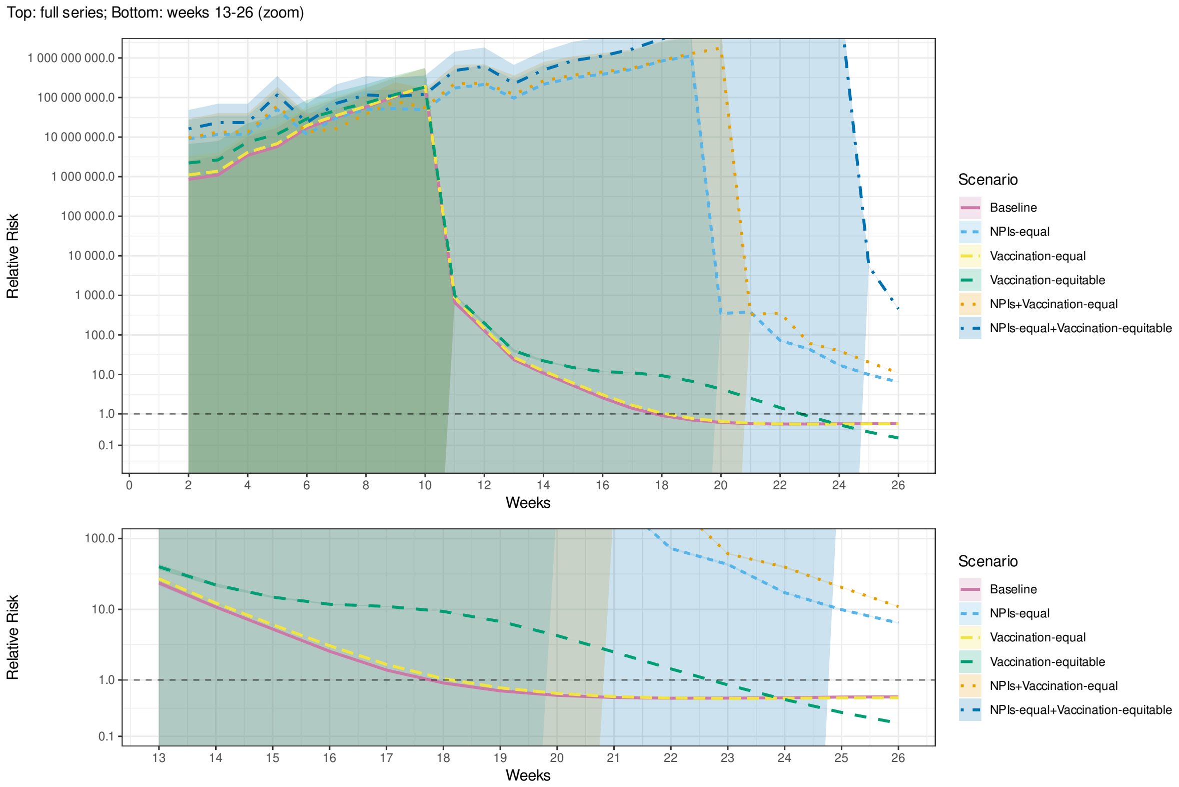


Fig. A.37. Relative risk of mortality in the least vulnerable (Q5) compared to the most vulnerable (Q1) municipalities by MHDI during the first 26 weeks of an outbreak in simulations of scenarios 1 and 5–9 (Table 3). with effect of mobility changes on transmission doubled ($\Delta M_i(t)$, Table 2). Shaded bands indicate 95% confidence intervals.

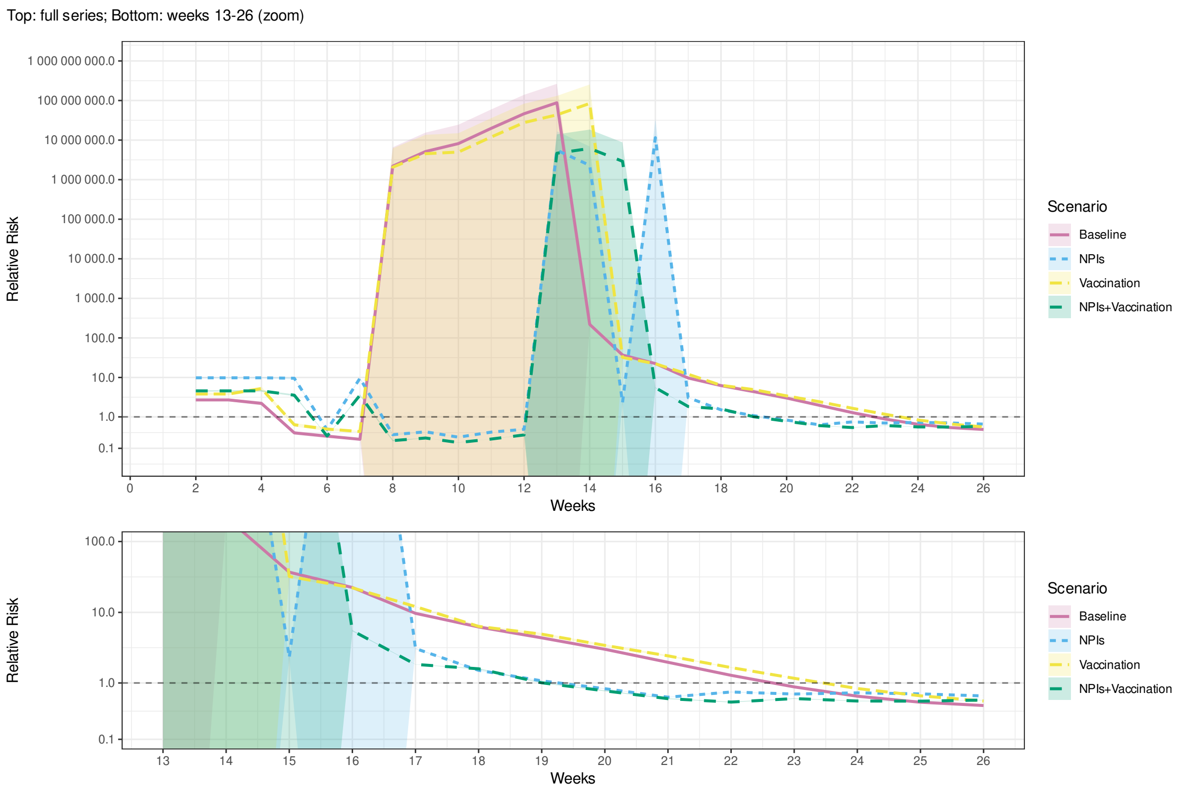


Fig. A.38. Relative risk of mortality in the least vulnerable (Q5) compared to the most vulnerable (Q1) municipalities by SVI during the first 26 weeks of an outbreak in the real world (*Empirical*) and simulations of scenarios 1–4 (Table 3). with lower bound of basic reproduction number ($R_0$, Table 2). Shaded bands indicate 95% confidence intervals.

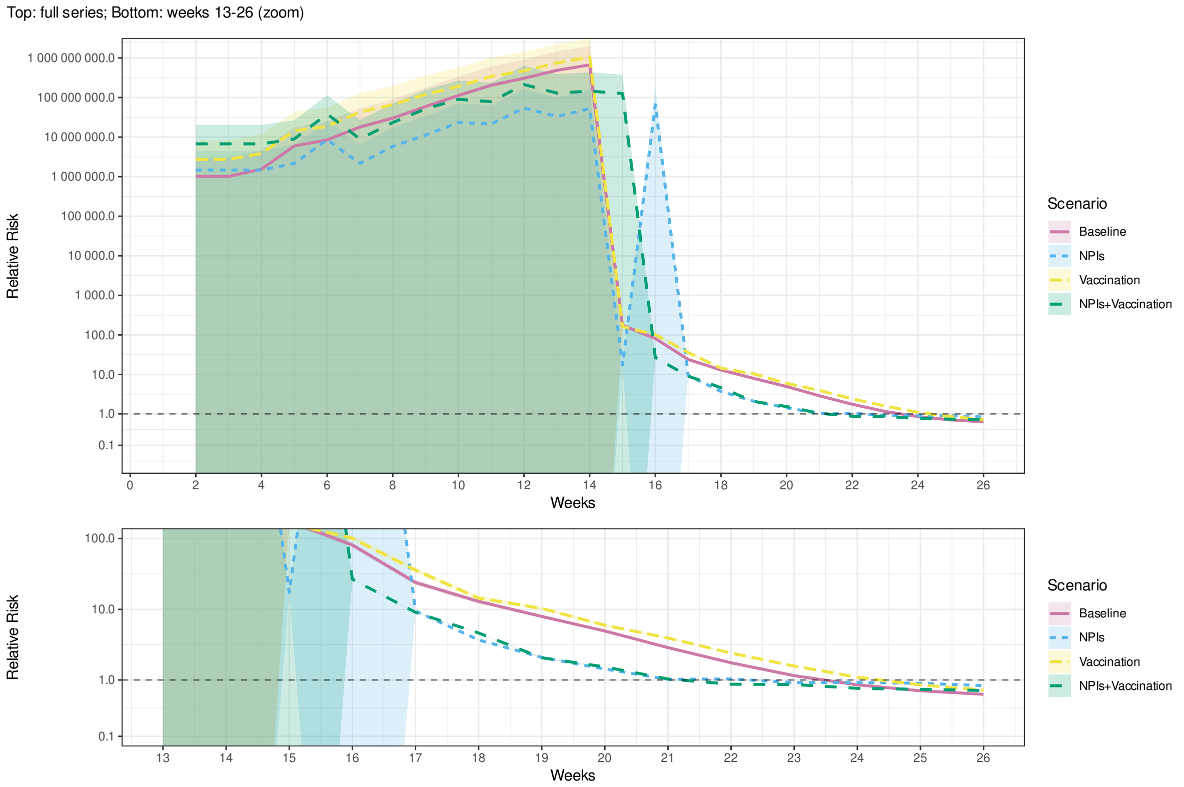


Fig. A.39. Relative risk of mortality in the least vulnerable (Q5) compared to the most vulnerable (Q1) municipalities by MHDI during the first 26 weeks of an outbreak in the real world (*Empirical*) and simulations of scenarios 1–4 (Table 3). with lower bound of basic reproduction number ($R_0$, Table 2). Shaded bands indicate 95% confidence intervals.

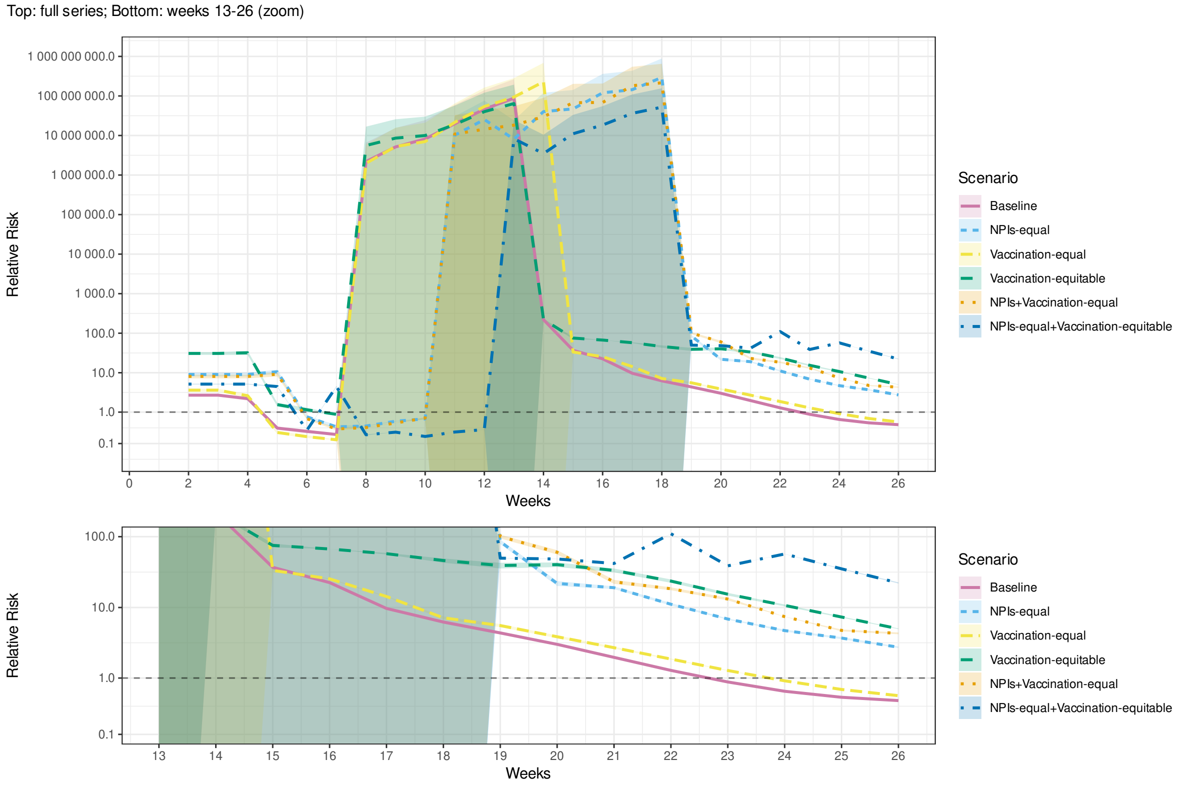


Fig. A.40. Relative risk of mortality in the least vulnerable (Q5) compared to the most vulnerable (Q1) municipalities by SVI during the first 26 weeks of an outbreak in simulations of scenarios 1 and 5–9 (Table 3). with lower bound of basic reproduction number ($R_0$, Table 2). Shaded bands indicate 95% confidence intervals.

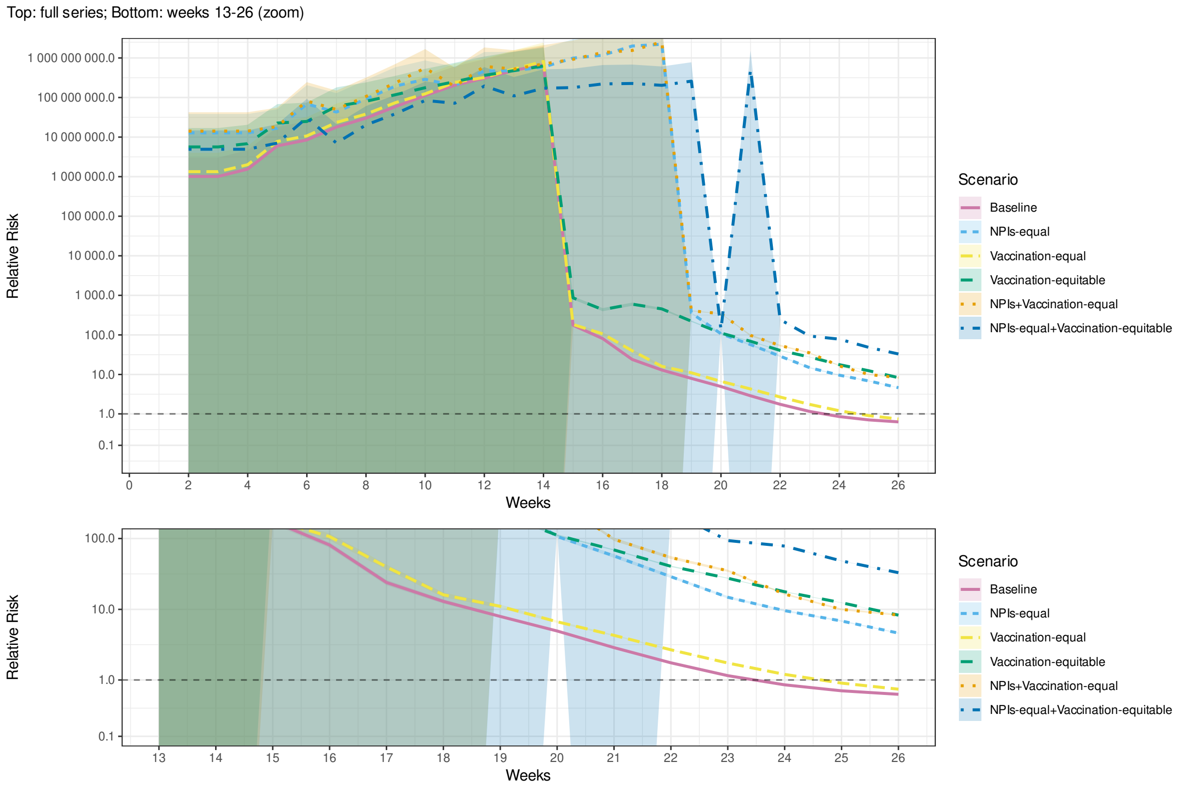


Fig. A.41. Relative risk of mortality in the least vulnerable (Q5) compared to the most vulnerable (Q1) municipalities by MHDI during the first 26 weeks of an outbreak in simulations of scenarios 1 and 5–9 (Table 3). with lower bound of basic reproduction number ($R_0$, Table 2). Shaded bands indicate 95% confidence intervals.

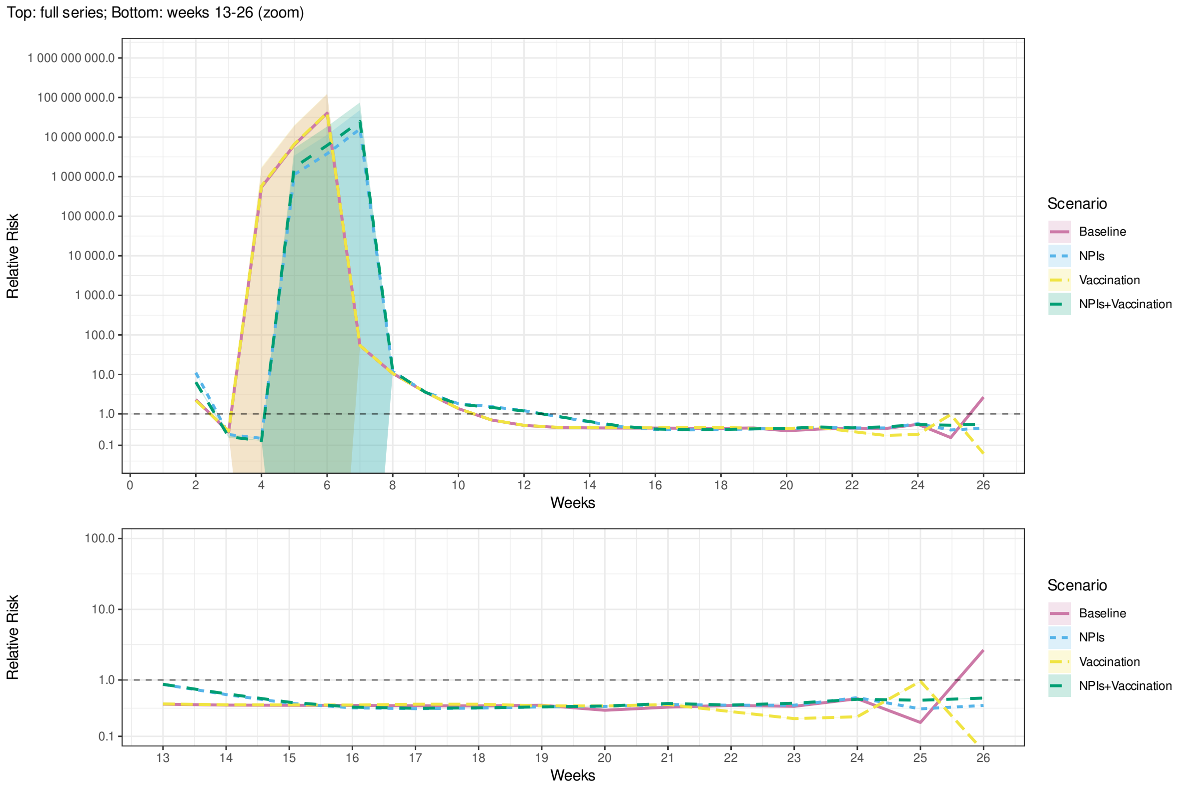


Fig. A.42. Relative risk of mortality in the least vulnerable (Q5) compared to the most vulnerable (Q1) municipalities by SVI during the first 26 weeks of an outbreak in the real world (*Empirical*) and simulations of scenarios 1–4 (Table 3). with upper bound of basic reproduction number ($R_0$, Table 2). Shaded bands indicate 95% confidence intervals.

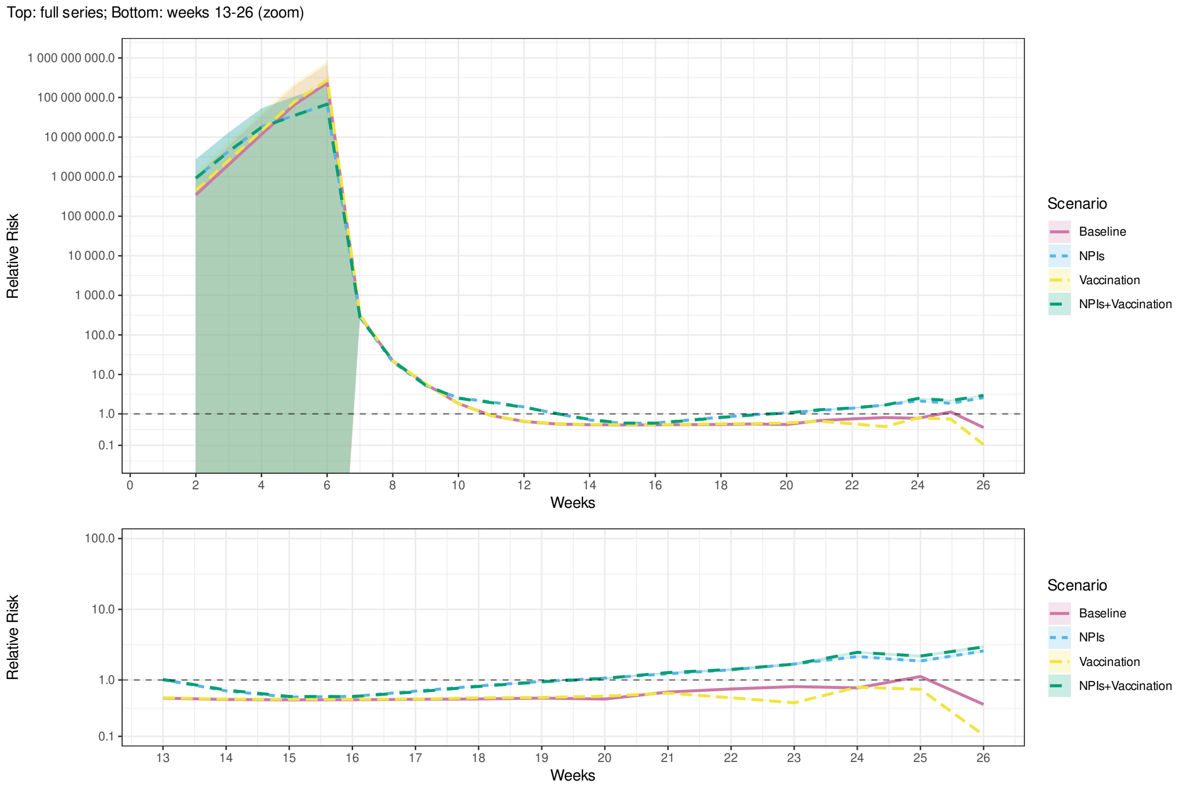


Fig. A.43. Relative risk of mortality in the least vulnerable (Q5) compared to the most vulnerable (Q1) municipalities by MHDI during the first 26 weeks of an outbreak in the real world (*Empirical*) and simulations of scenarios 1–4 (Table 3). with upper bound of basic reproduction number ($R_0$, Table 2). Shaded bands indicate 95% confidence intervals.

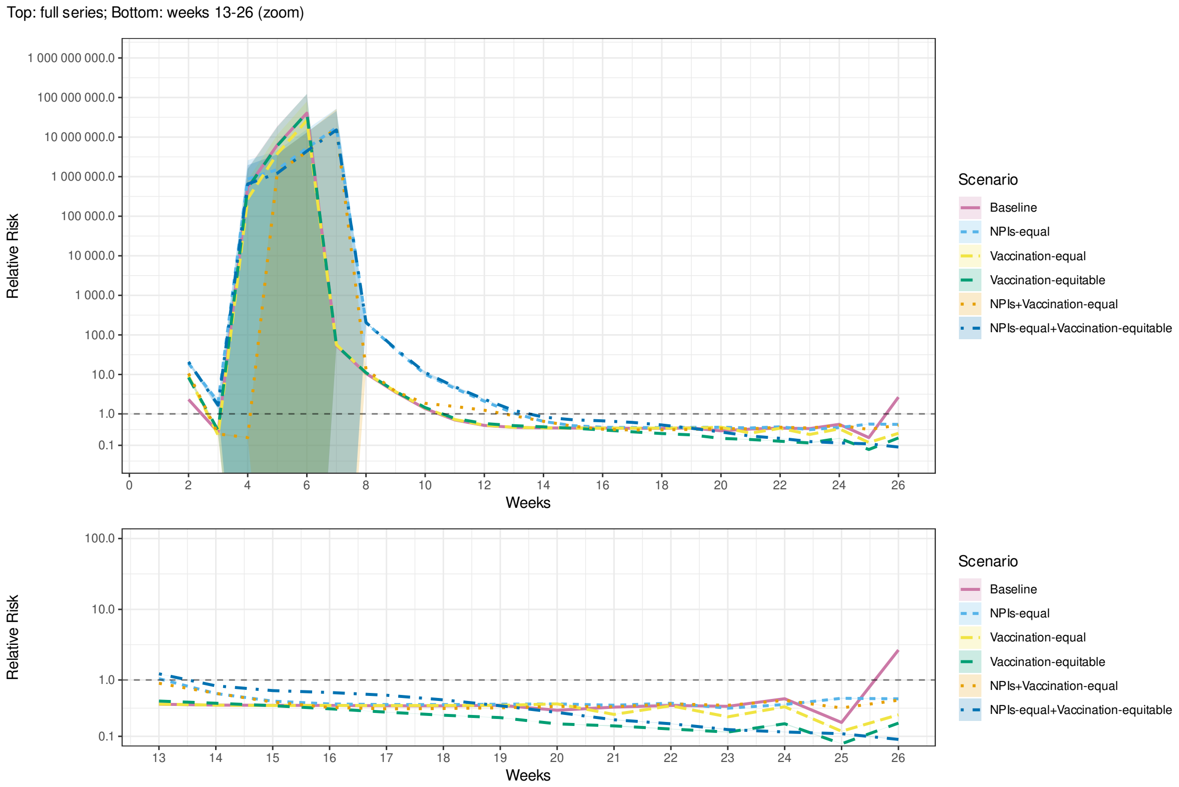


Fig. A.44. Relative risk of mortality in the least vulnerable (Q5) compared to the most vulnerable (Q1) municipalities by SVI during the first 26 weeks of an outbreak in simulations of scenarios 1 and 5–9 (Table 3). with upper bound of basic reproduction number ($R_0$, Table 2). Shaded bands indicate 95% confidence intervals.

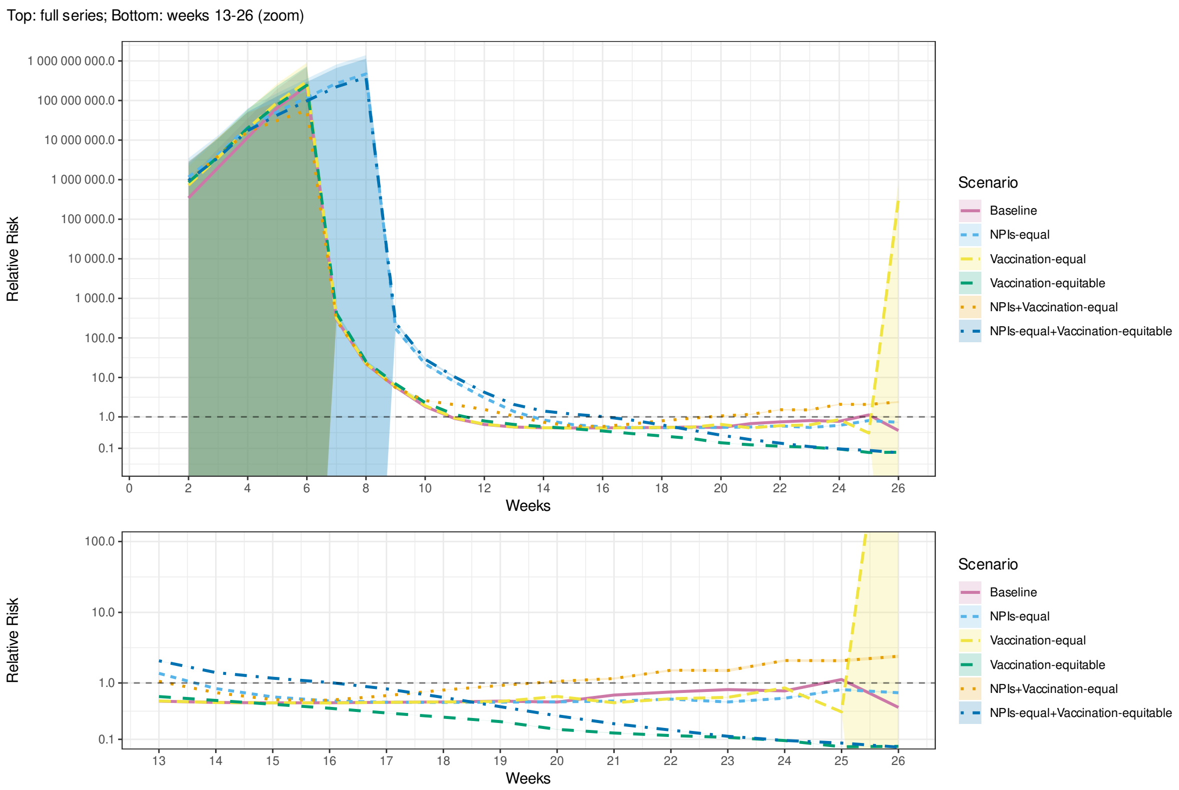


Fig. A.45. Relative risk of mortality in the least vulnerable (Q5) compared to the most vulnerable (Q1) municipalities by MHDI during the first 26 weeks of an outbreak in simulations of scenarios 1 and 5–9 (Table 3). with upper bound of basic reproduction number ($R_0$, Table 2). Shaded bands indicate 95% confidence intervals.

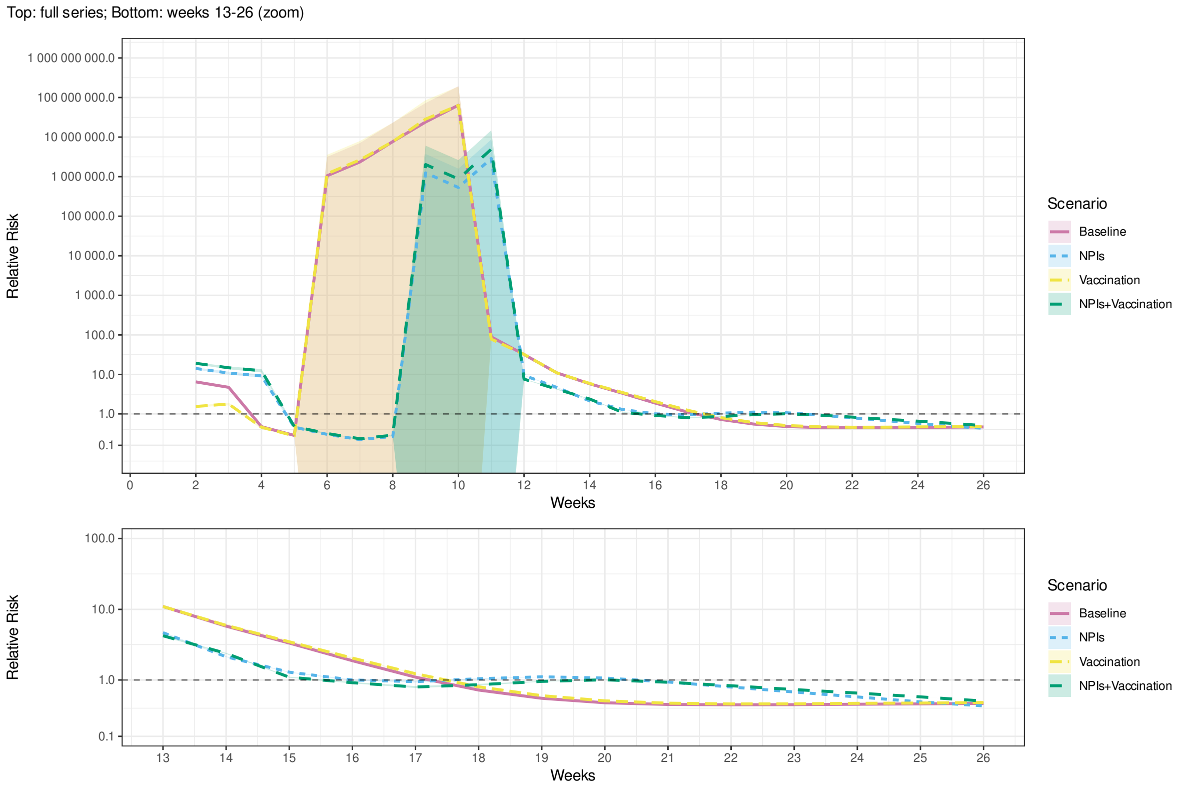


Fig. A.46. Relative risk of mortality in the least vulnerable (Q5) compared to the most vulnerable (Q1) municipalities by SVI during the first 26 weeks of an outbreak in the real world (*Empirical*) and simulations of scenarios 1–4 (Table 3). with lower bound of household secondary attack rate ($SAR$, Table 2). Shaded bands indicate 95% confidence intervals.

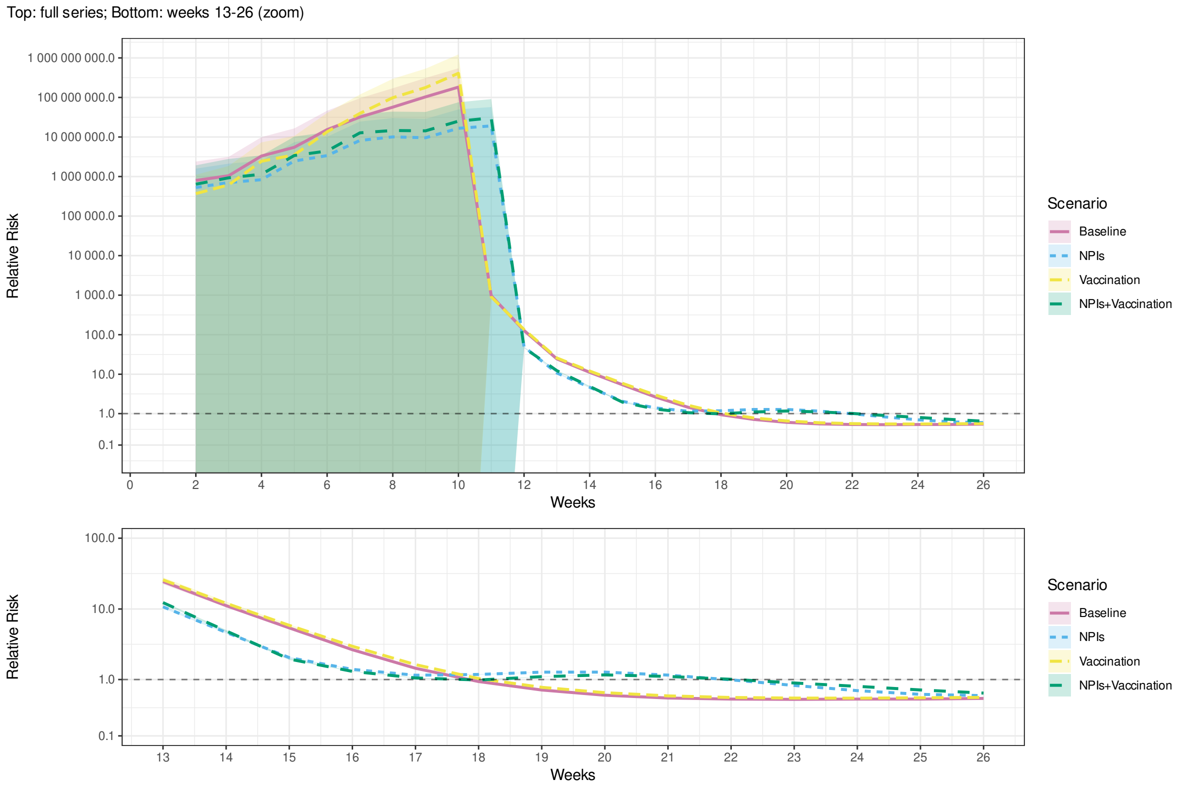


Fig. A.47. Relative risk of mortality in the least vulnerable (Q5) compared to the most vulnerable (Q1) municipalities by MHDI during the first 26 weeks of an outbreak in the real world (*Empirical*) and simulations of scenarios 1–4 (Table 3). with lower bound of household secondary attack rate ($SAR$, Table 2). Shaded bands indicate 95% confidence intervals.

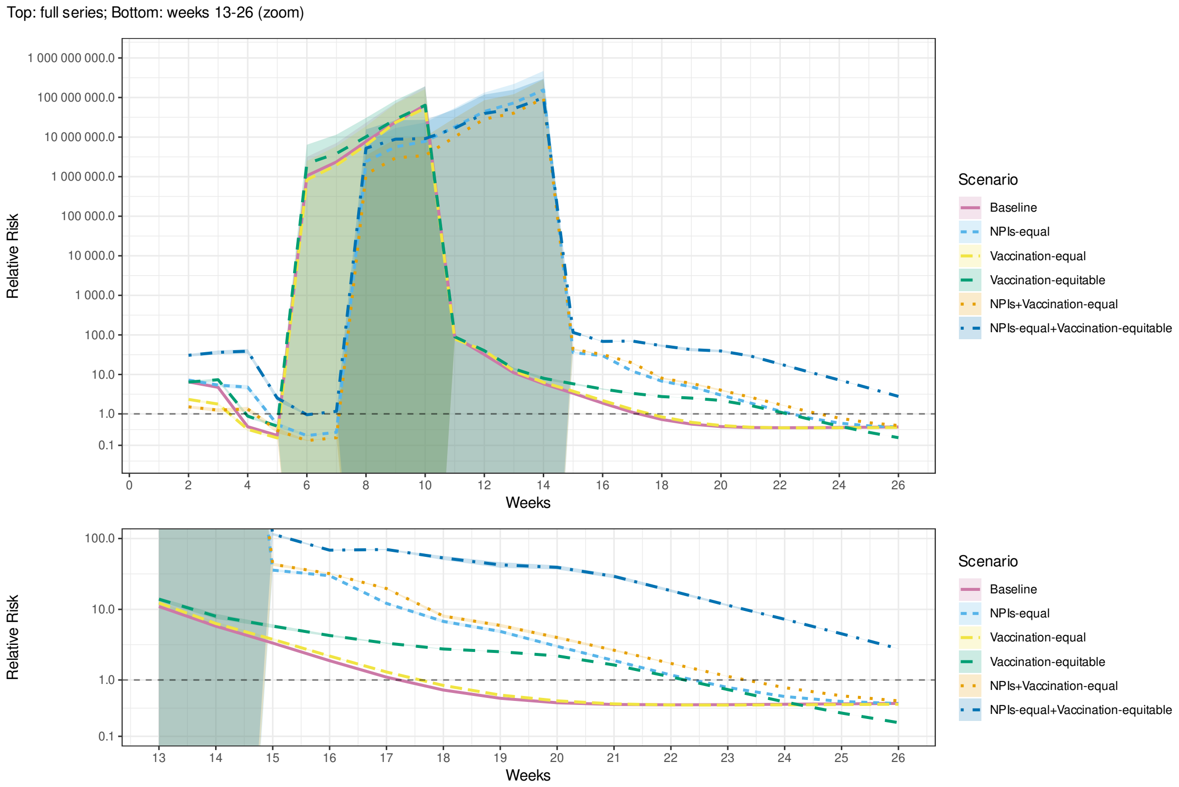


Fig. A.48. Relative risk of mortality in the least vulnerable (Q5) compared to the most vulnerable (Q1) municipalities by SVI during the first 26 weeks of an outbreak in simulations of scenarios 1 and 5–9 (Table 3). with lower bound of household secondary attack rate ($SAR$, Table 2). Shaded bands indicate 95% confidence intervals.

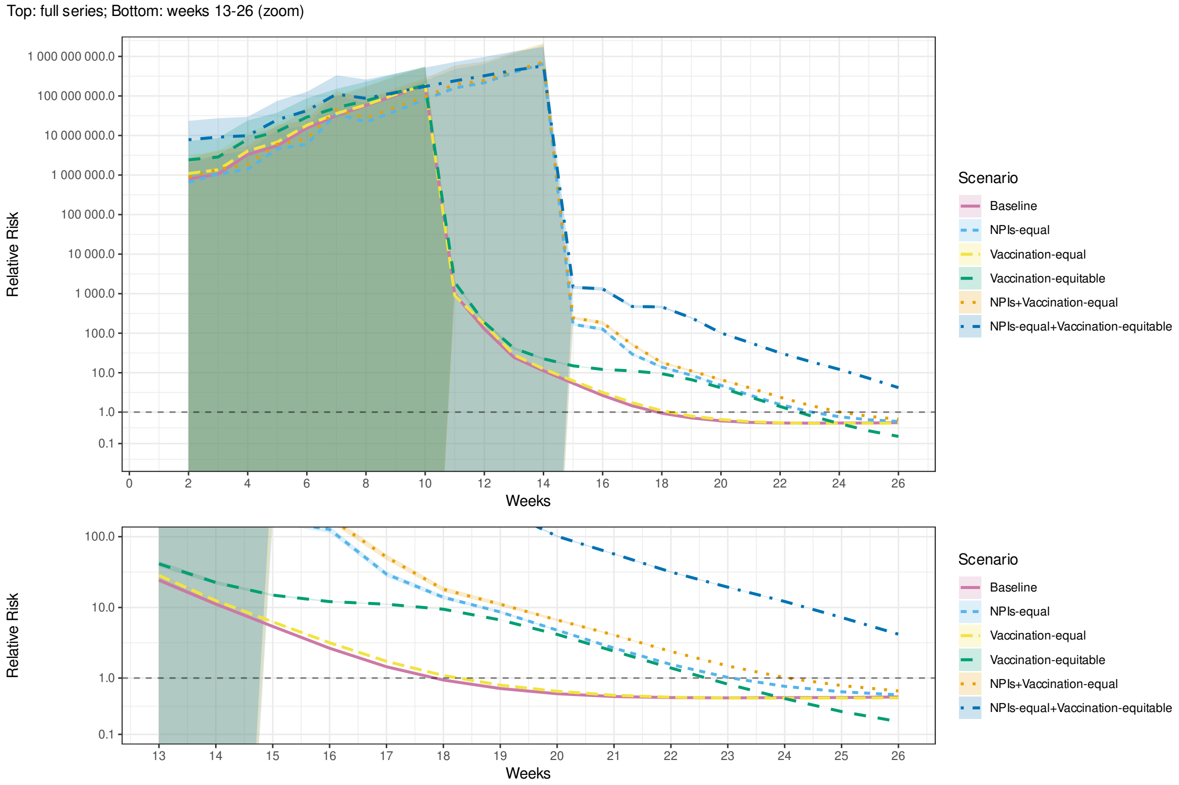


Fig. A.49. Relative risk of mortality in the least vulnerable (Q5) compared to the most vulnerable (Q1) municipalities by MHDI during the first 26 weeks of an outbreak in simulations of scenarios 1 and 5–9 (Table 3). with lower bound of household secondary attack rate ($SAR$, Table 2). Shaded bands indicate 95% confidence intervals.

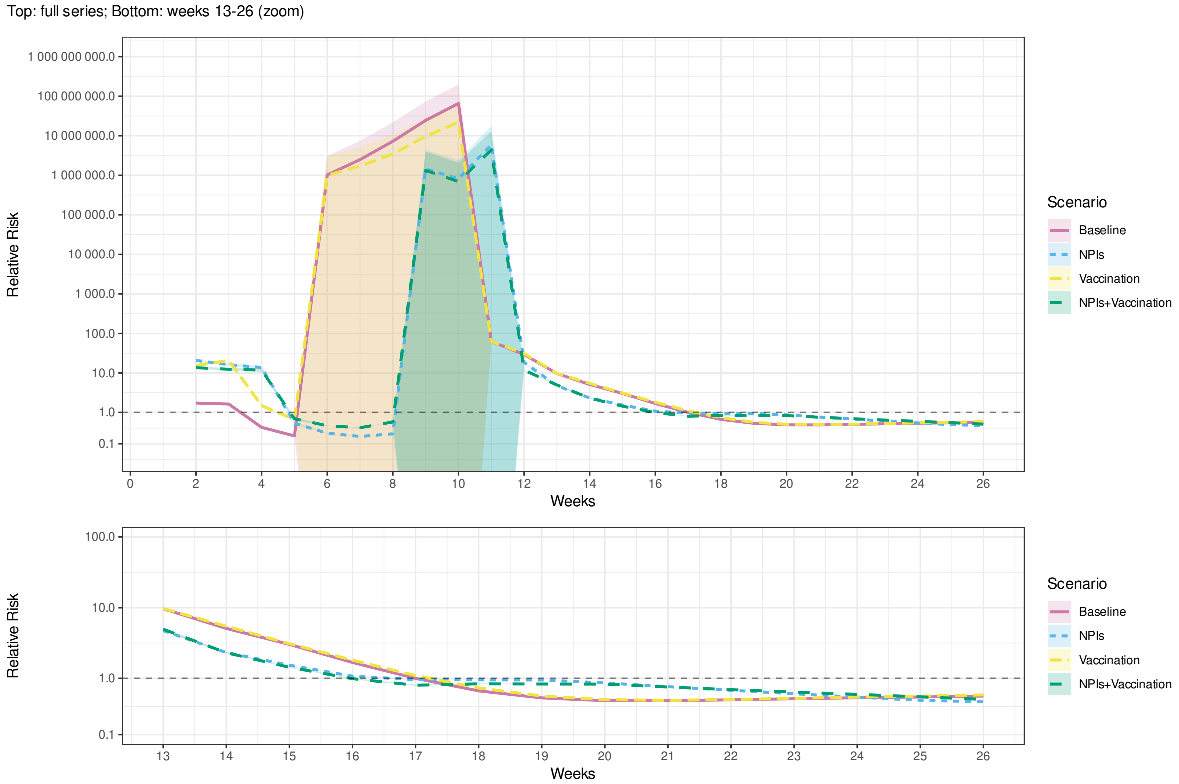


Fig. A.50. Relative risk of mortality in the least vulnerable (Q5) compared to the most vulnerable (Q1) municipalities by SVI during the first 26 weeks of an outbreak in the real world (*Empirical*) and simulations of scenarios 1–4 (Table 3). with upper bound of household secondary attack rate (*SAR*, Table 2). Shaded bands indicate 95% confidence intervals.

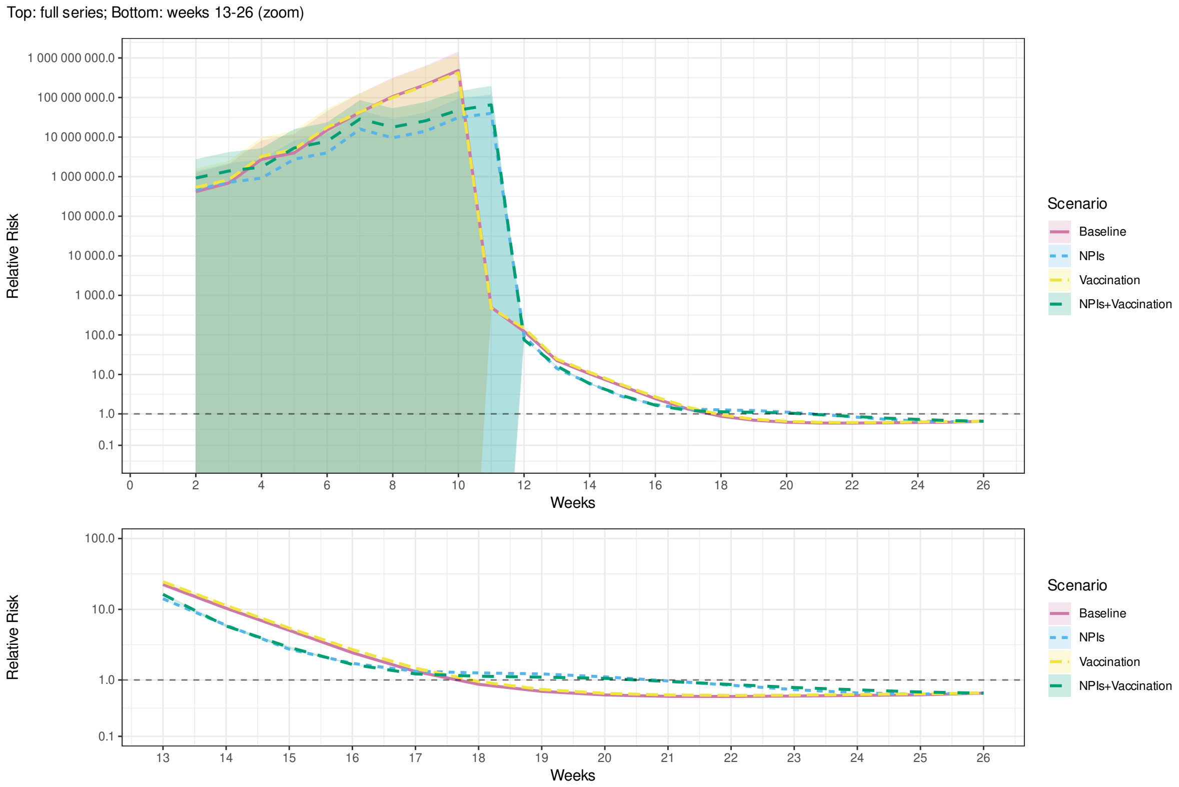


Fig. A.51. Relative risk of mortality in the least vulnerable (Q5) compared to the most vulnerable (Q1) municipalities by MHDI during the first 26 weeks of an outbreak in the real world (*Empirical*) and simulations of scenarios 1–4 (Table 3). with upper bound of household secondary attack rate (*SAR*, Table 2). Shaded bands indicate 95% confidence intervals.

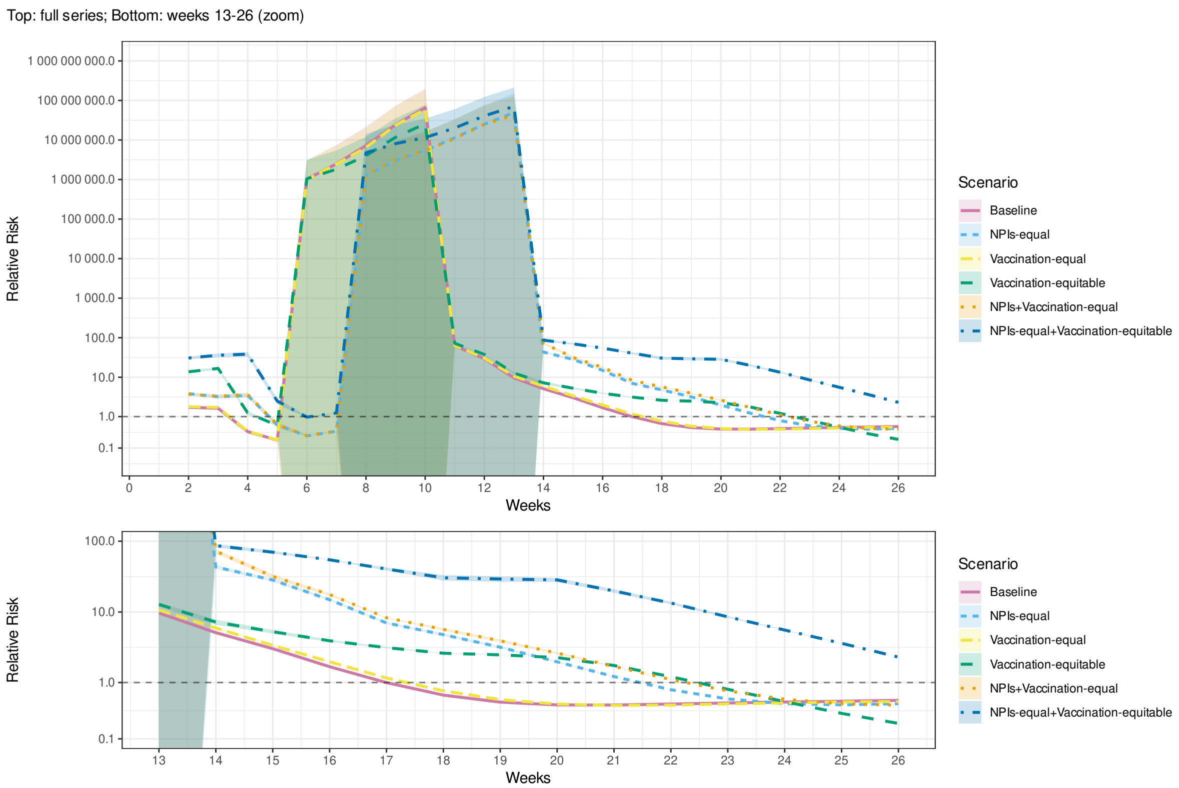


Fig. A.52. Relative risk of mortality in the least vulnerable (Q5) compared to the most vulnerable (Q1) municipalities by SVI during the first 26 weeks of an outbreak in simulations of scenarios 1 and 5–9 (Table 3). with upper bound of household secondary attack rate ($SAR$, Table 2). Shaded bands indicate 95% confidence intervals.

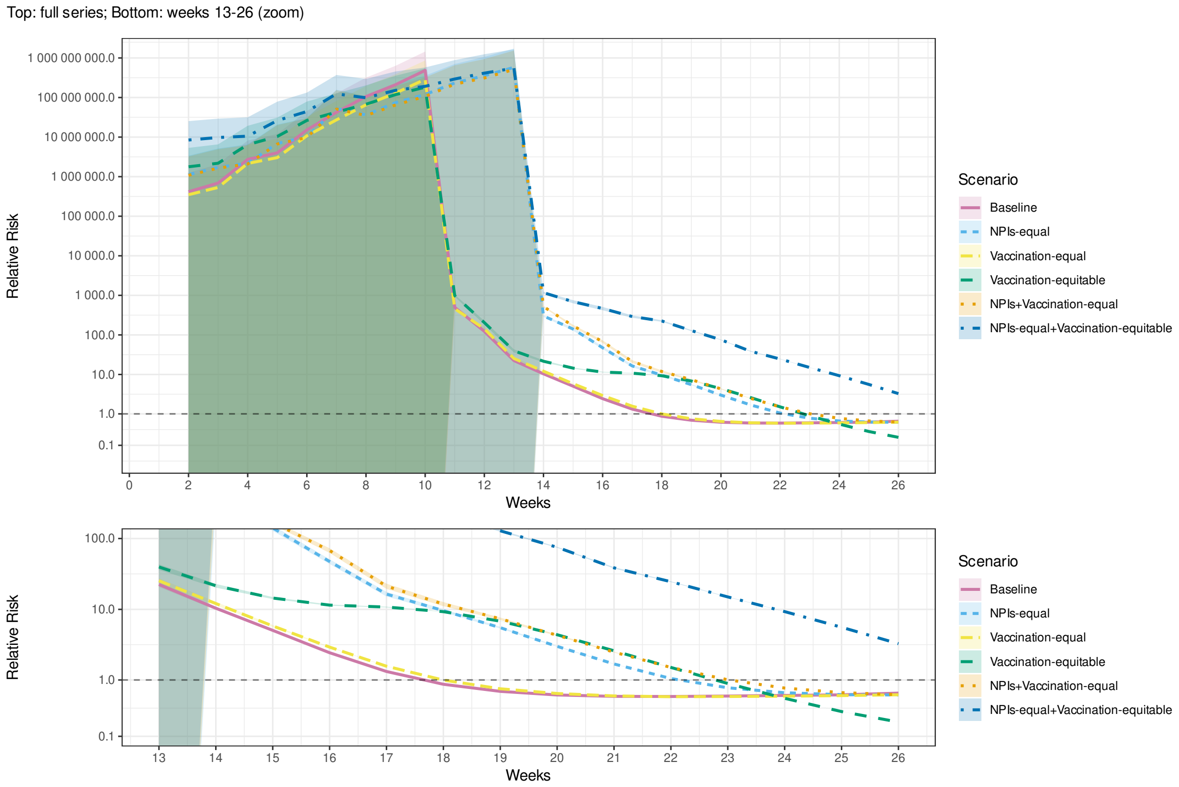


Fig. A.53. Relative risk of mortality in the least vulnerable (Q5) compared to the most vulnerable (Q1) municipalities by MHDI during the first 26 weeks of an outbreak in simulations of scenarios 1 and 5–9 (Table 3). with upper bound of household secondary attack rate ($SAR$, Table 2). Shaded bands indicate 95% confidence intervals.

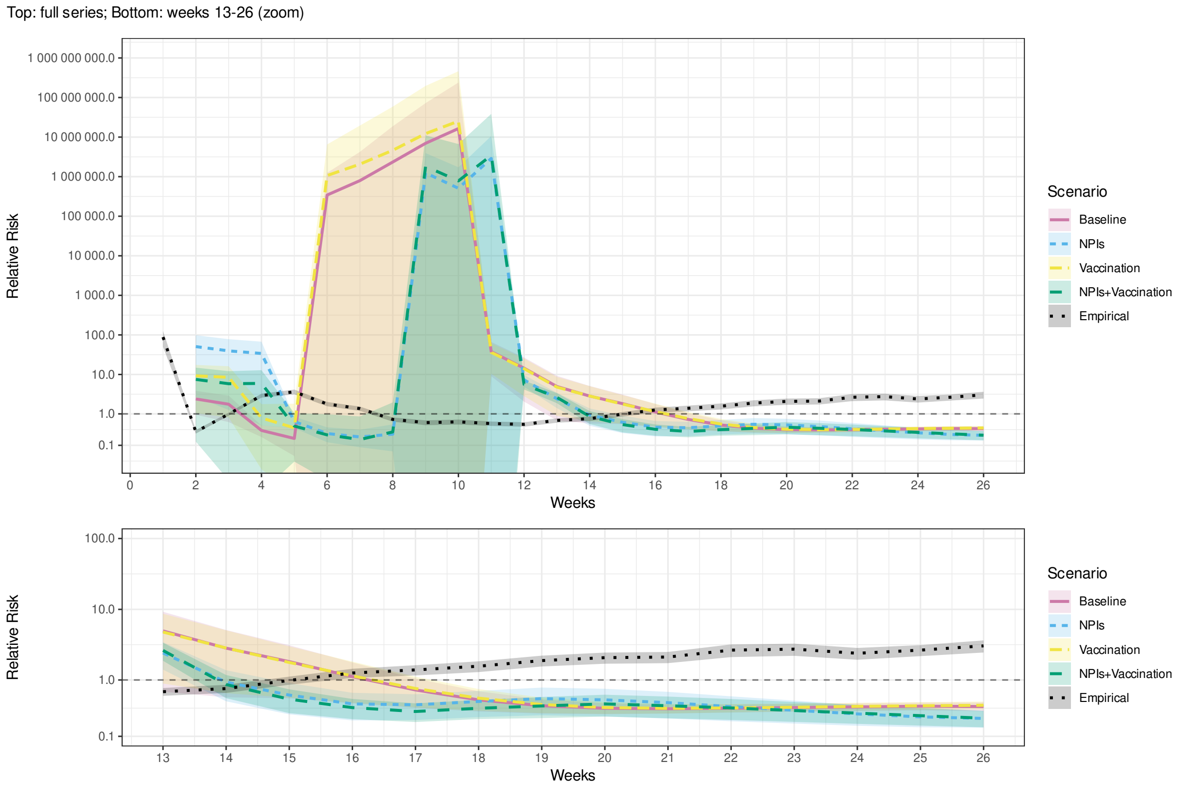


Fig. A.54. Relative risk of mortality in the least vulnerable (Q5) compared to the most vulnerable (Q1) municipalities by SVI during the first 26 weeks of an outbreak in the real world (*Empirical*) and simulations of scenarios 1–4 (Table 3). Additional regression controls: region fixed-effects. Shaded bands indicate 95% confidence intervals.

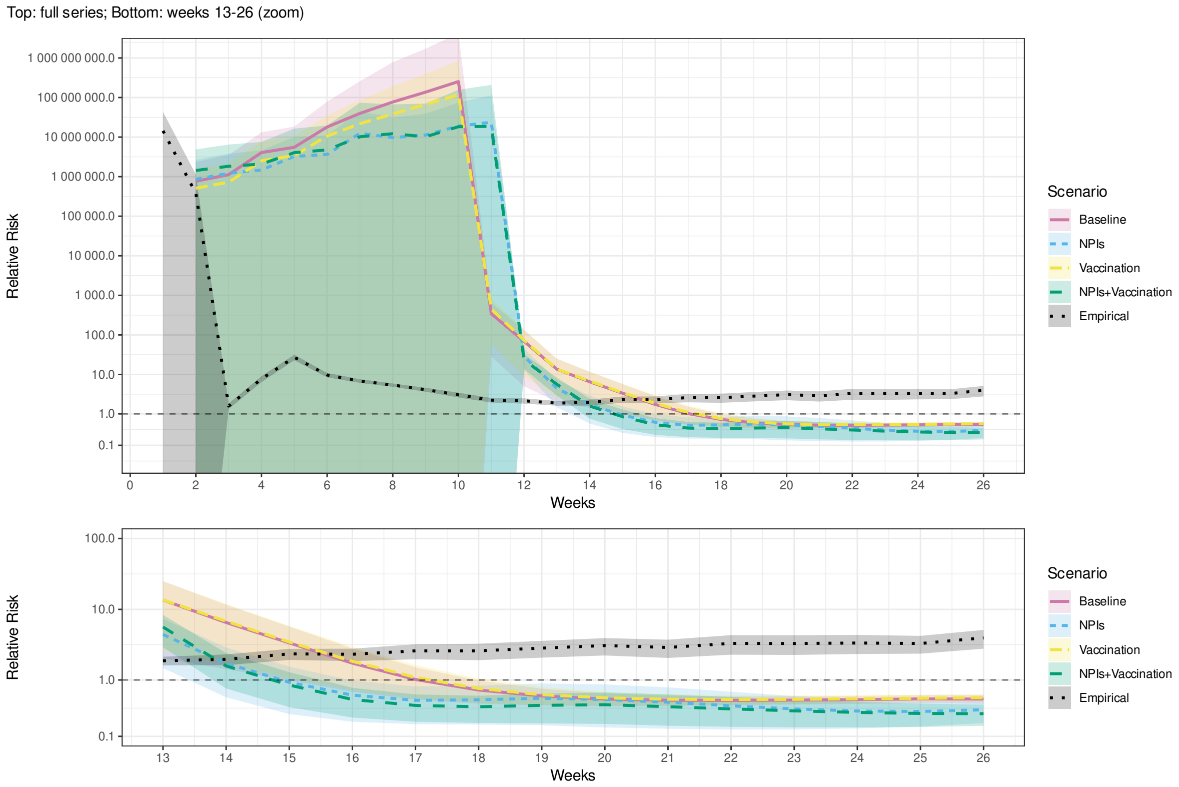


Fig. A.55. Relative risk of mortality in the least vulnerable (Q5) compared to the most vulnerable (Q1) municipalities by MHDI during the first 26 weeks of an outbreak in the real world (*Empirical*) and simulations of scenarios 1–4 (Table 3). Additional regression controls: region fixed-effects. Shaded bands indicate 95% confidence intervals.

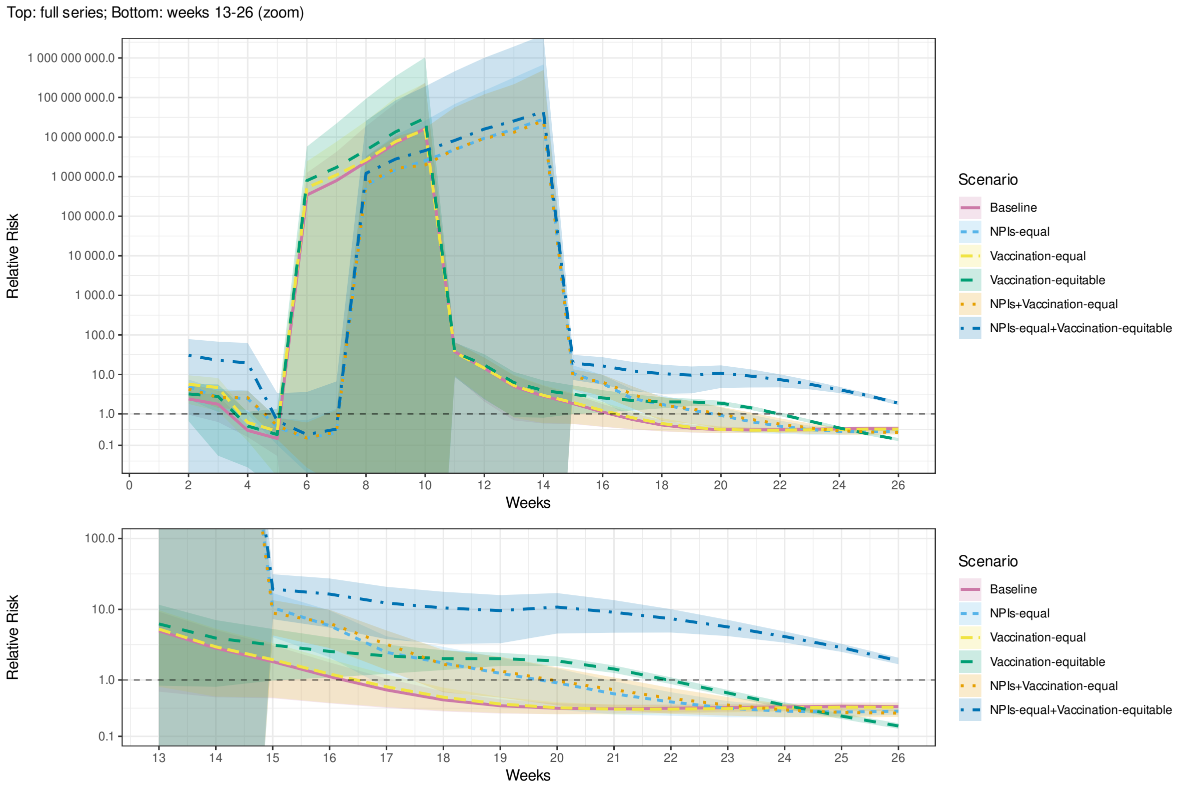


Fig. A.56. Relative risk of mortality in the least vulnerable (Q5) compared to the most vulnerable (Q1) municipalities by SVI during the first 26 weeks of an outbreak in simulations of scenarios 1 and 5–9 (Table 3). Additional regression controls: region fixed-effects. Shaded bands indicate 95% confidence intervals.

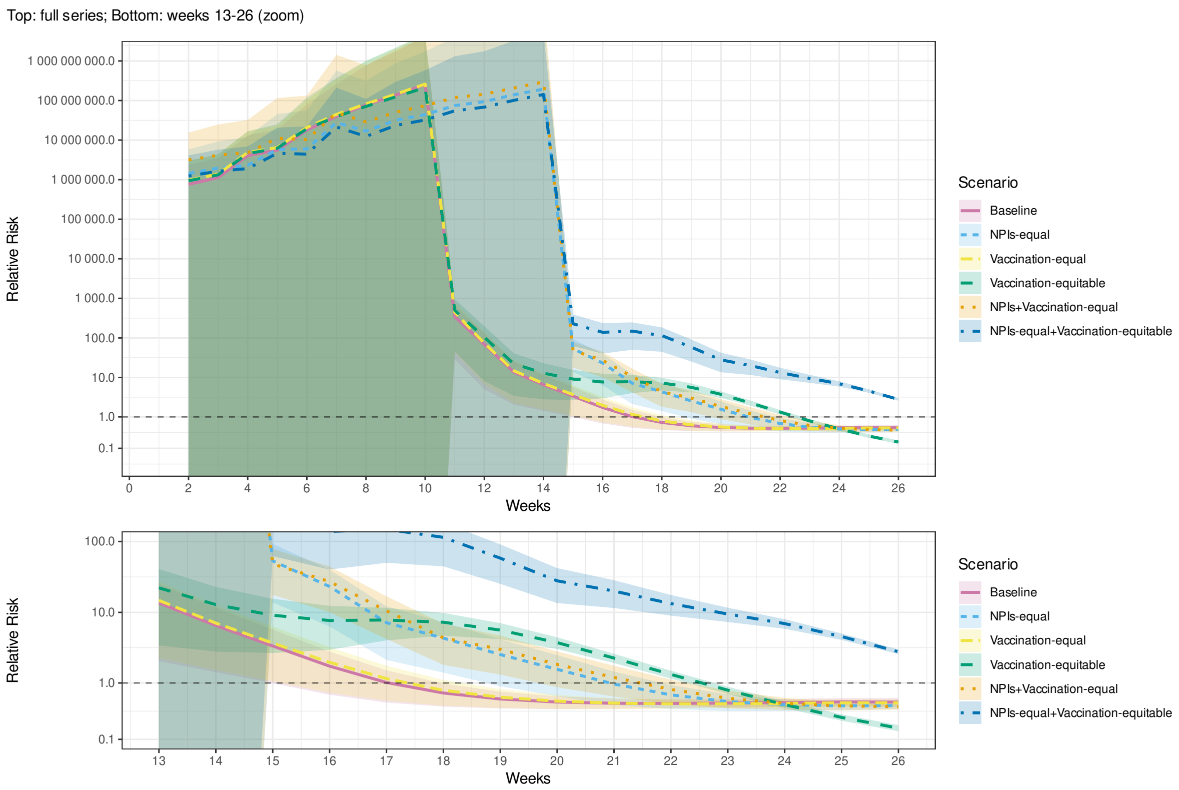


Fig. A.57. Relative risk of mortality in the least vulnerable (Q5) compared to the most vulnerable (Q1) municipalities by MHDI during the first 26 weeks of an outbreak in simulations of scenarios 1 and 5–9 (Table 3). Additional regression controls: region fixed-effects. Shaded bands indicate 95% confidence intervals.

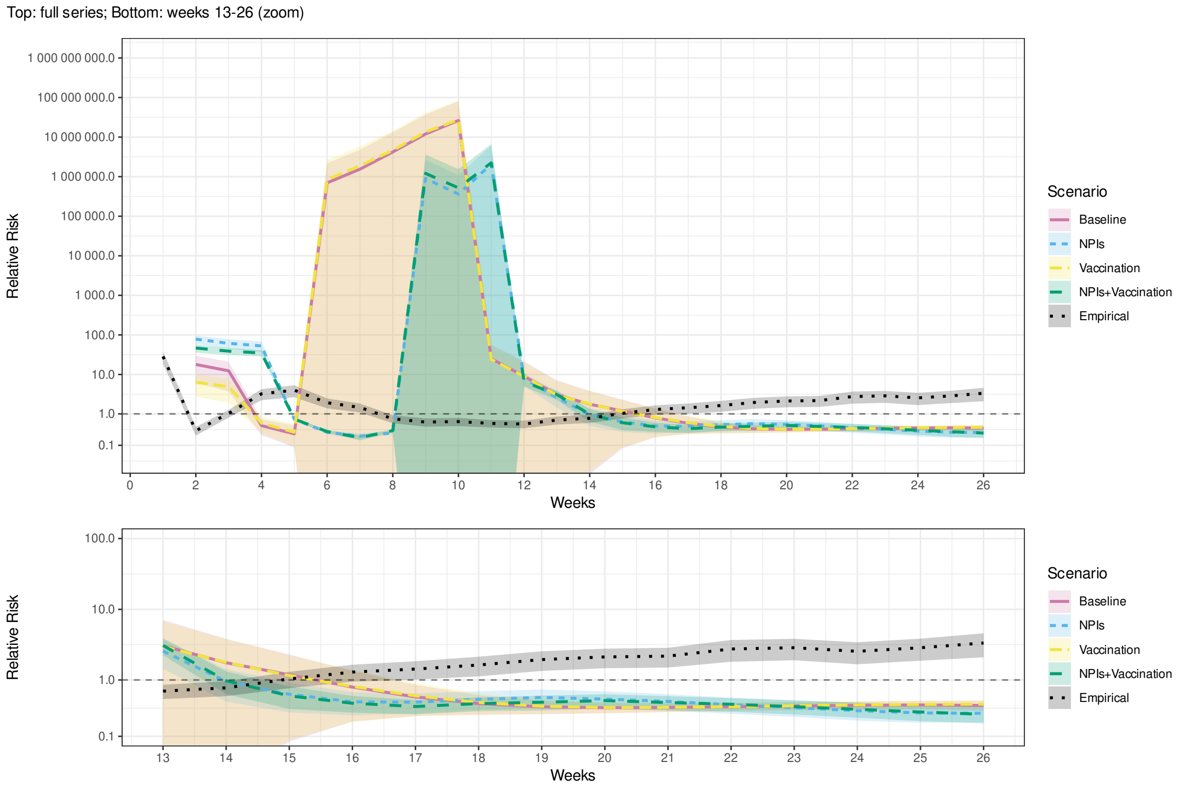


Fig. A.58. Relative risk of mortality in the least vulnerable (Q5) compared to the most vulnerable (Q1) municipalities by SVI during the first 26 weeks of an outbreak in the real world (*Empirical*) and simulations of scenarios 1–4 (Table 3). Additional regression controls: state fixed-effects. Shaded bands indicate 95% confidence intervals.

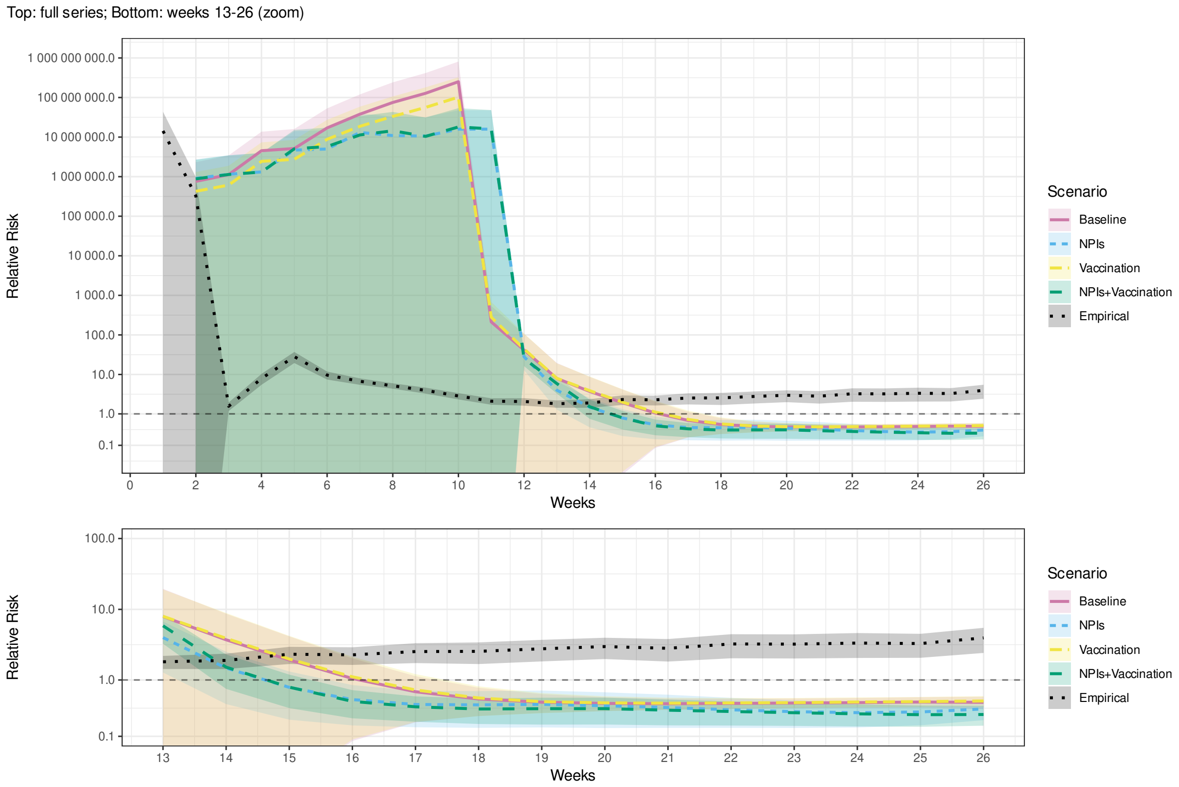


Fig. A.59. Relative risk of mortality in the least vulnerable (Q5) compared to the most vulnerable (Q1) municipalities by MHDI during the first 26 weeks of an outbreak in the real world (*Empirical*) and simulations of scenarios 1–4 (Table 3). Additional regression controls: state fixed-effects. Shaded bands indicate 95% confidence intervals.

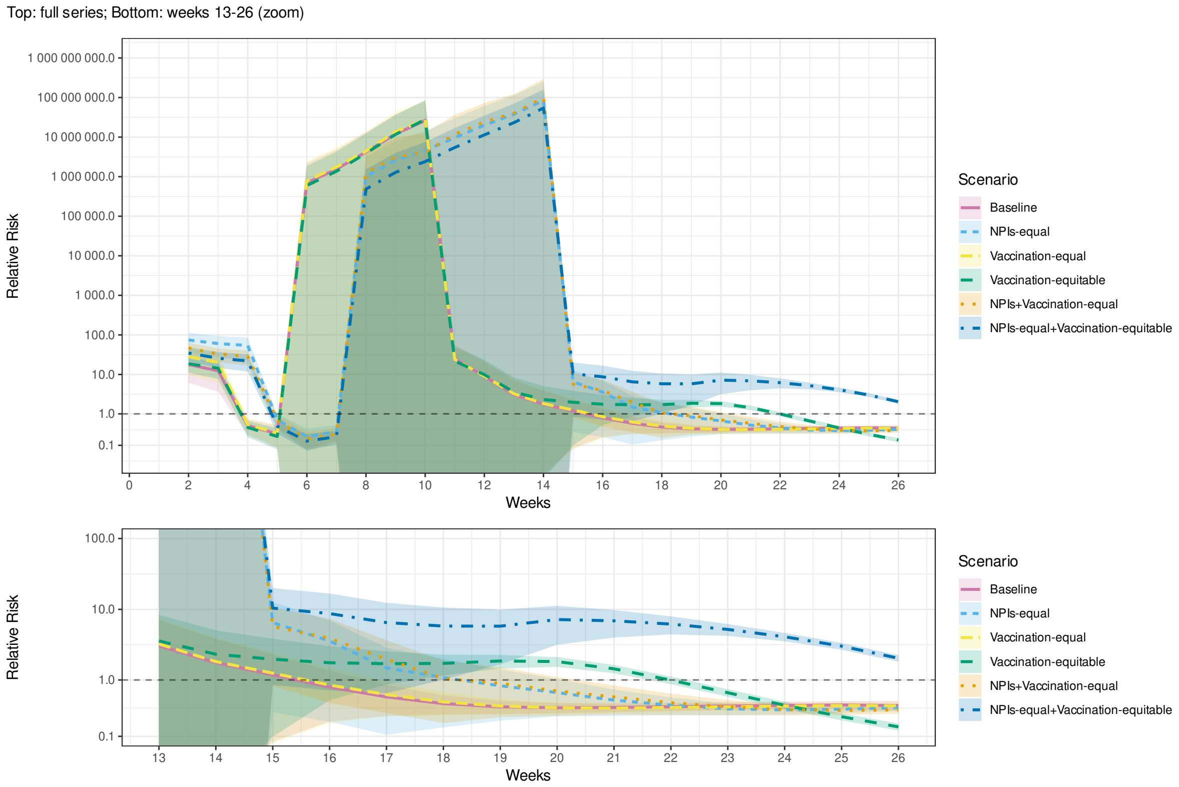


Fig. A.60. Relative risk of mortality in the least vulnerable (Q5) compared to the most vulnerable (Q1) municipalities by SVI during the first 26 weeks of an outbreak in simulations of scenarios 1 and 5–9 (Table 3). Additional regression controls: state fixed-effects. Shaded bands indicate 95% confidence intervals.

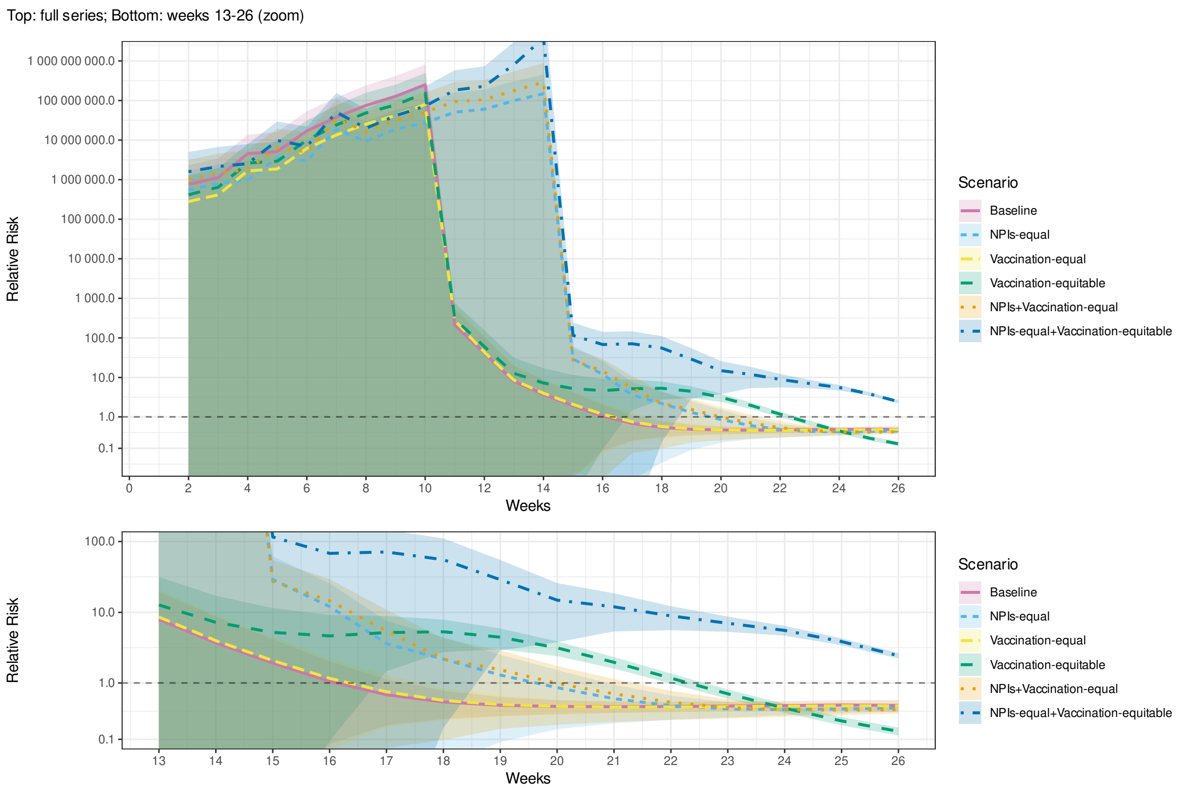


Fig. A.61. Relative risk of mortality in the least vulnerable (Q5) compared to the most vulnerable (Q1) municipalities by MHDI during the first 26 weeks of an outbreak in simulations of scenarios 1 and 5–9 (Table 3). Additional regression controls: state fixed-effects. Shaded bands indicate 95% confidence intervals.

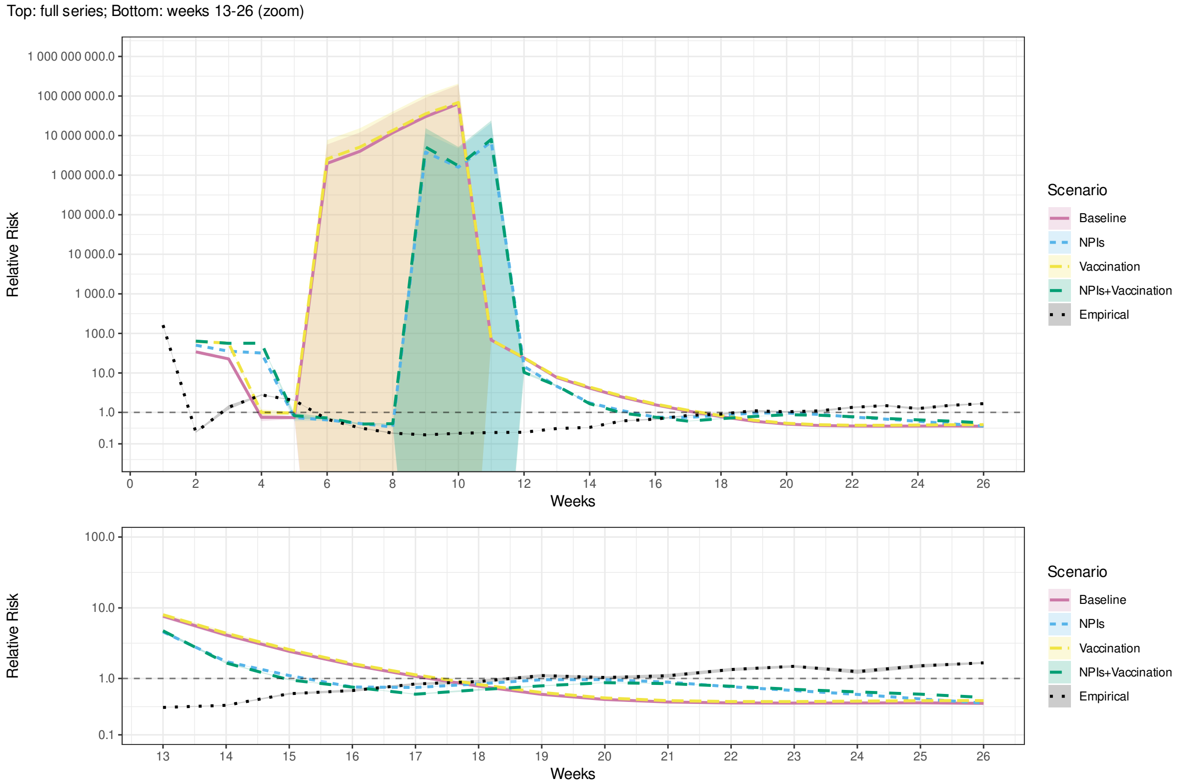


Fig. A.62. Relative risk of mortality in the least vulnerable (Q5) compared to the most vulnerable (Q1) municipalities by SVI during the first 26 weeks of an outbreak in the real world (*Empirical*) and simulations of scenarios 1–4 (Table 3). Additional regression controls: Bolsonaro vote share. Shaded bands indicate 95% confidence intervals.

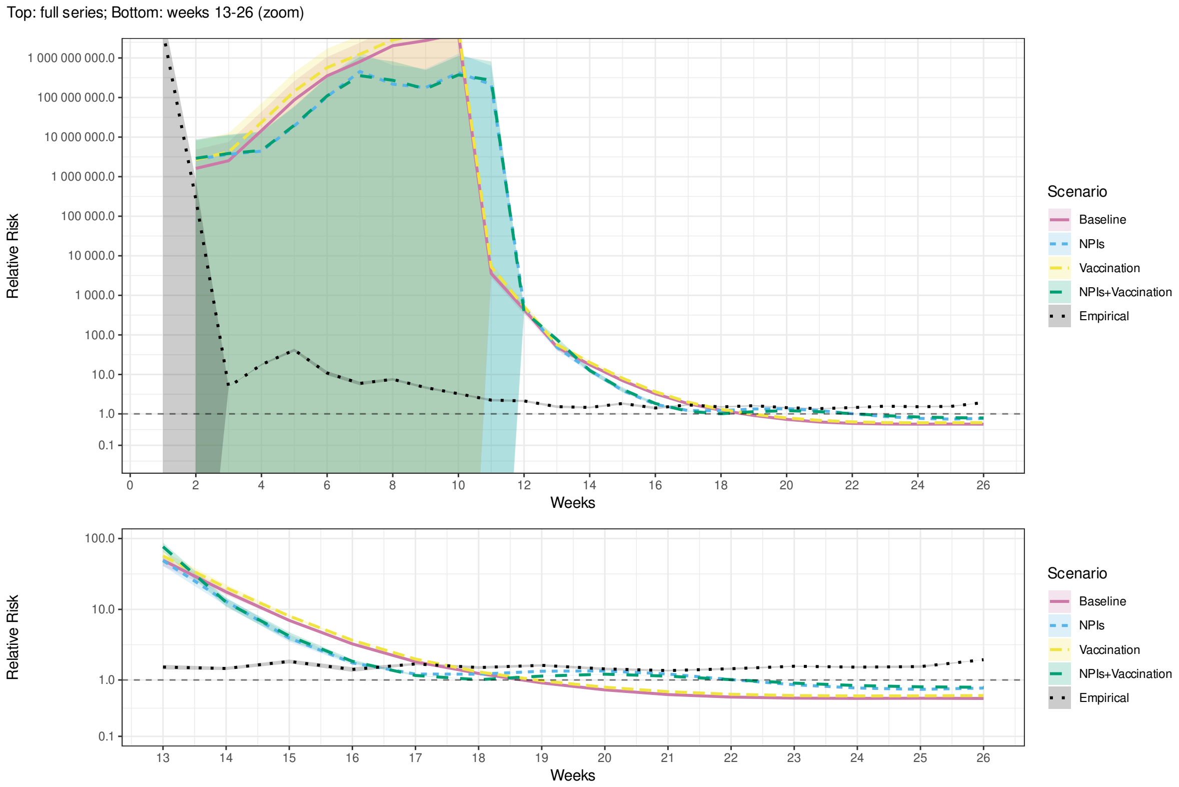


Fig. A.63. Relative risk of mortality in the least vulnerable (Q5) compared to the most vulnerable (Q1) municipalities by MHDI during the first 26 weeks of an outbreak in the real world (*Empirical*) and simulations of scenarios 1–4 (Table 3). Additional regression controls: Bolsonaro vote share. Shaded bands indicate 95% confidence intervals.

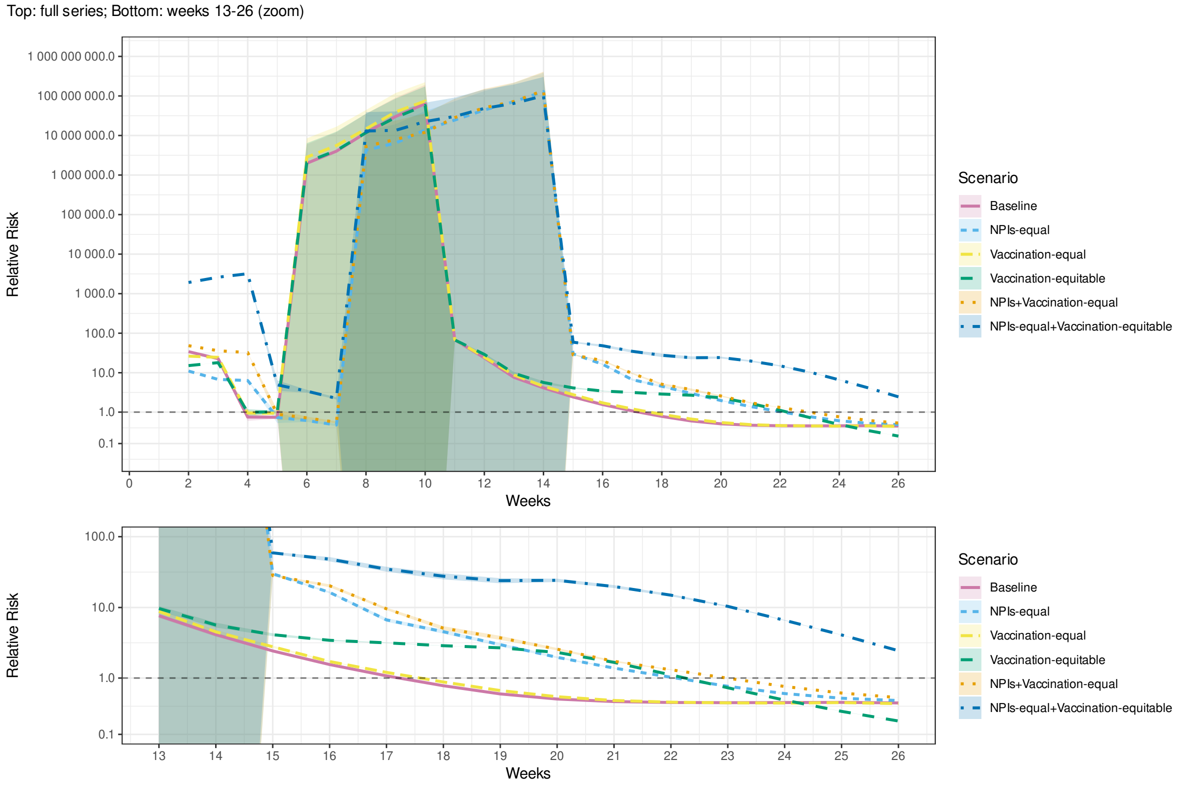


Fig. A.64. Relative risk of mortality in the least vulnerable (Q5) compared to the most vulnerable (Q1) municipalities by SVI during the first 26 weeks of an outbreak in simulations of scenarios 1 and 5–9 (Table 3). Additional regression controls: Bolsonaro vote share. Shaded bands indicate 95% confidence intervals.

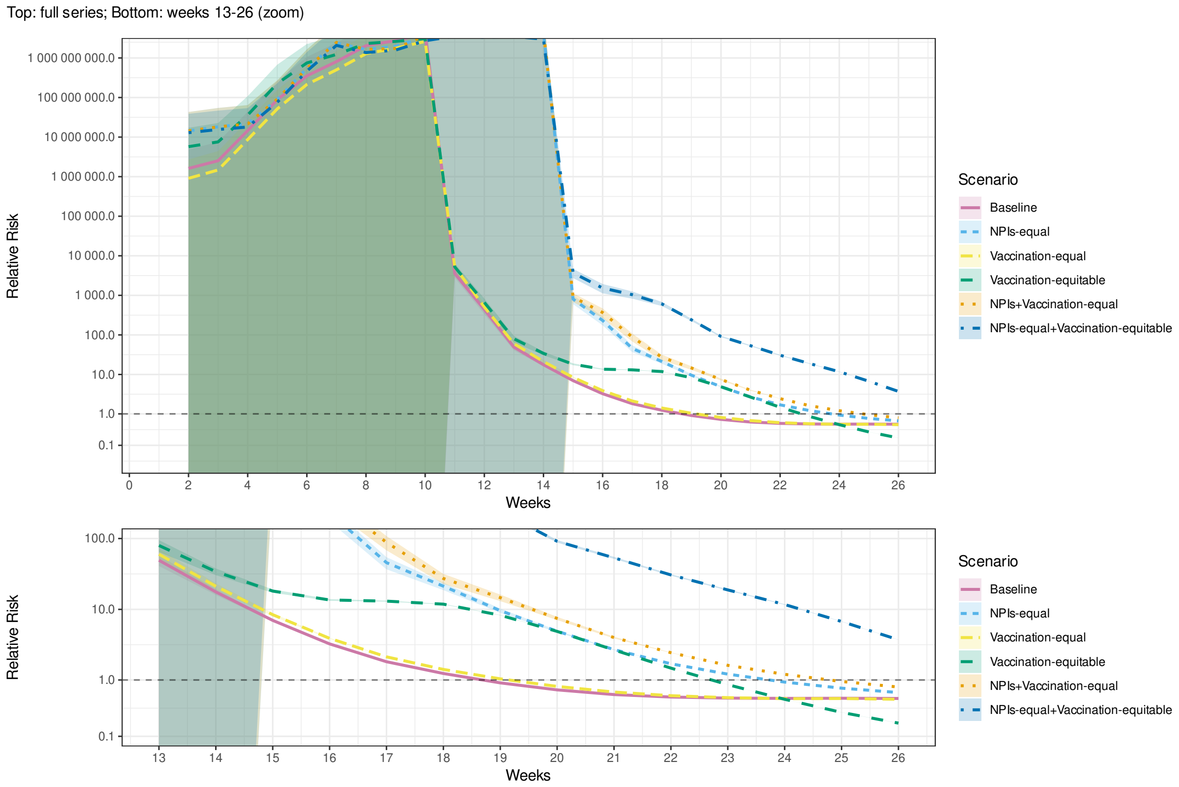


Fig. A.65. Relative risk of mortality in the least vulnerable (Q5) compared to the most vulnerable (Q1) municipalities by MHDI during the first 26 weeks of an outbreak in simulations of scenarios 1 and 5–9 (Table 3). Additional regression controls: Bolsonaro vote share. Shaded bands indicate 95% confidence intervals.

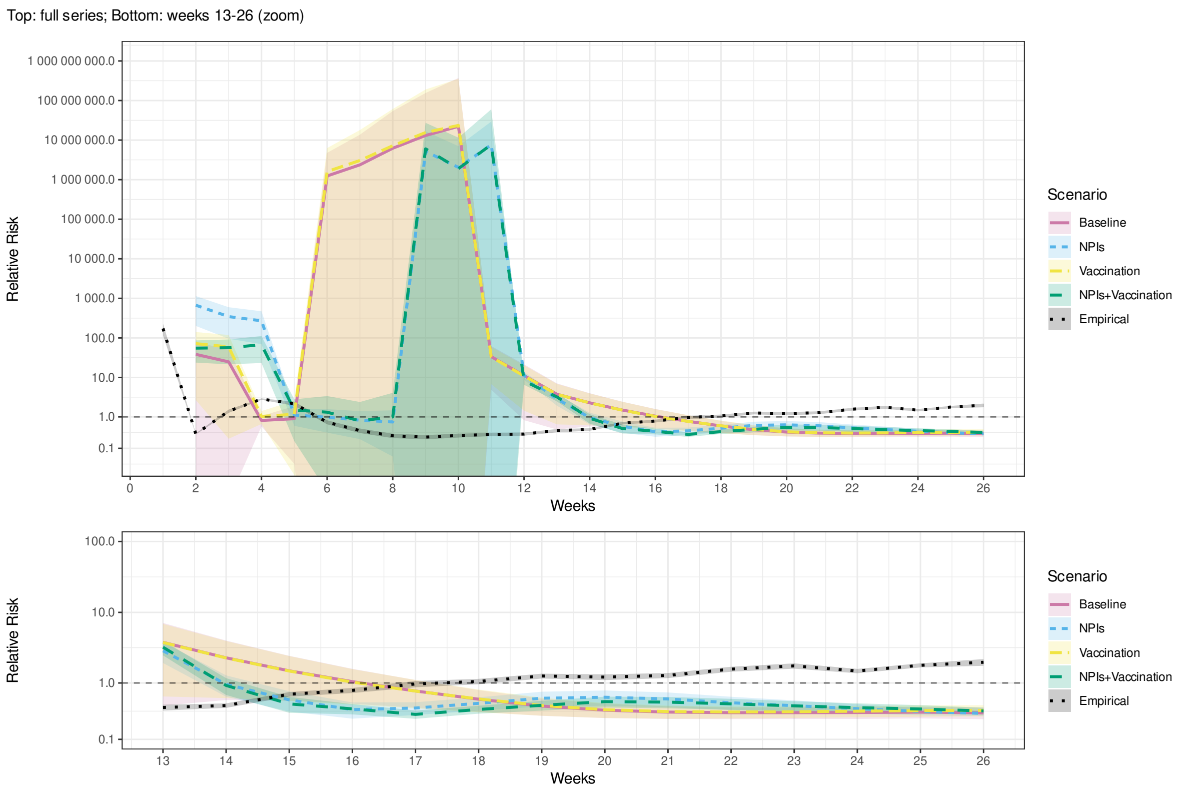


Fig. A.66. Relative risk of mortality in the least vulnerable (Q5) compared to the most vulnerable (Q1) municipalities by SVI during the first 26 weeks of an outbreak in the real world (*Empirical*) and simulations of scenarios 1–4 (Table 3). Additional regression controls: Bolsonaro vote share and region fixed-effects. Shaded bands indicate 95% confidence intervals.

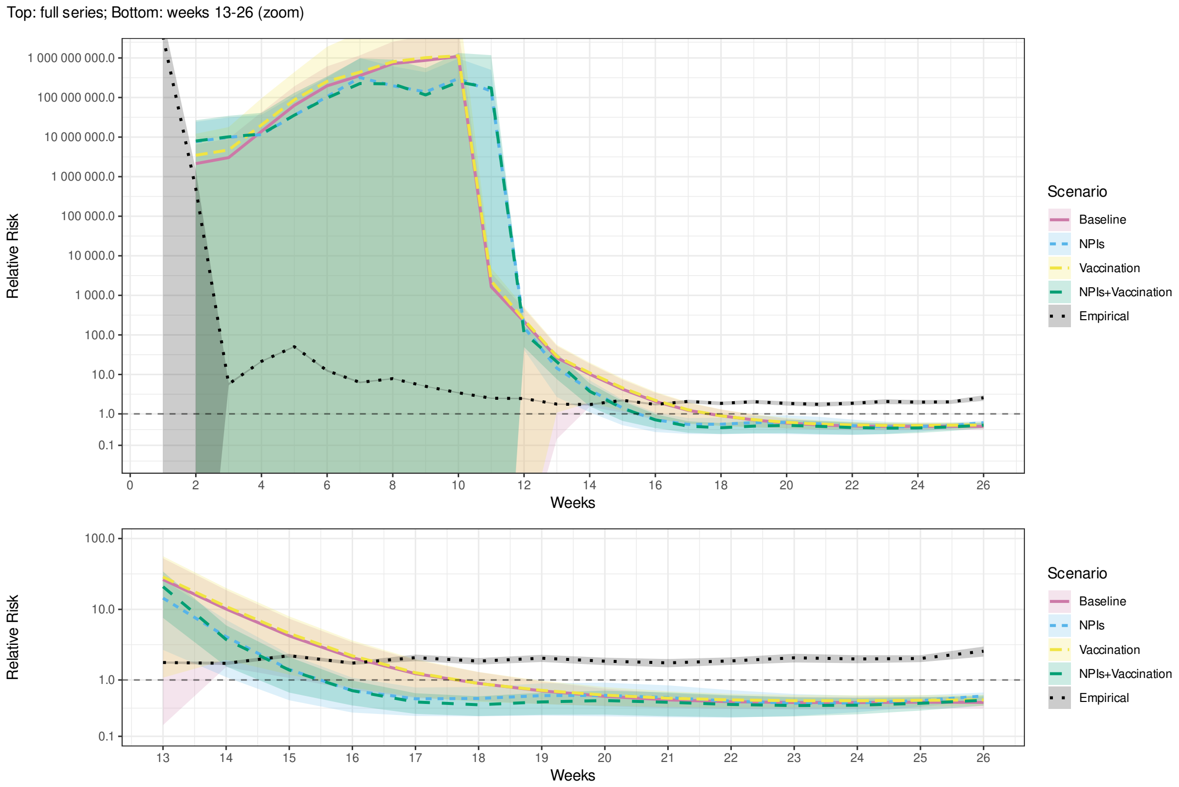


Fig. A.67. Relative risk of mortality in the least vulnerable (Q5) compared to the most vulnerable (Q1) municipalities by MHDI during the first 26 weeks of an outbreak in the real world (*Empirical*) and simulations of scenarios 1–4 (Table 3). Additional regression controls: Bolsonaro vote share and region fixed-effects. Shaded bands indicate 95% confidence intervals.

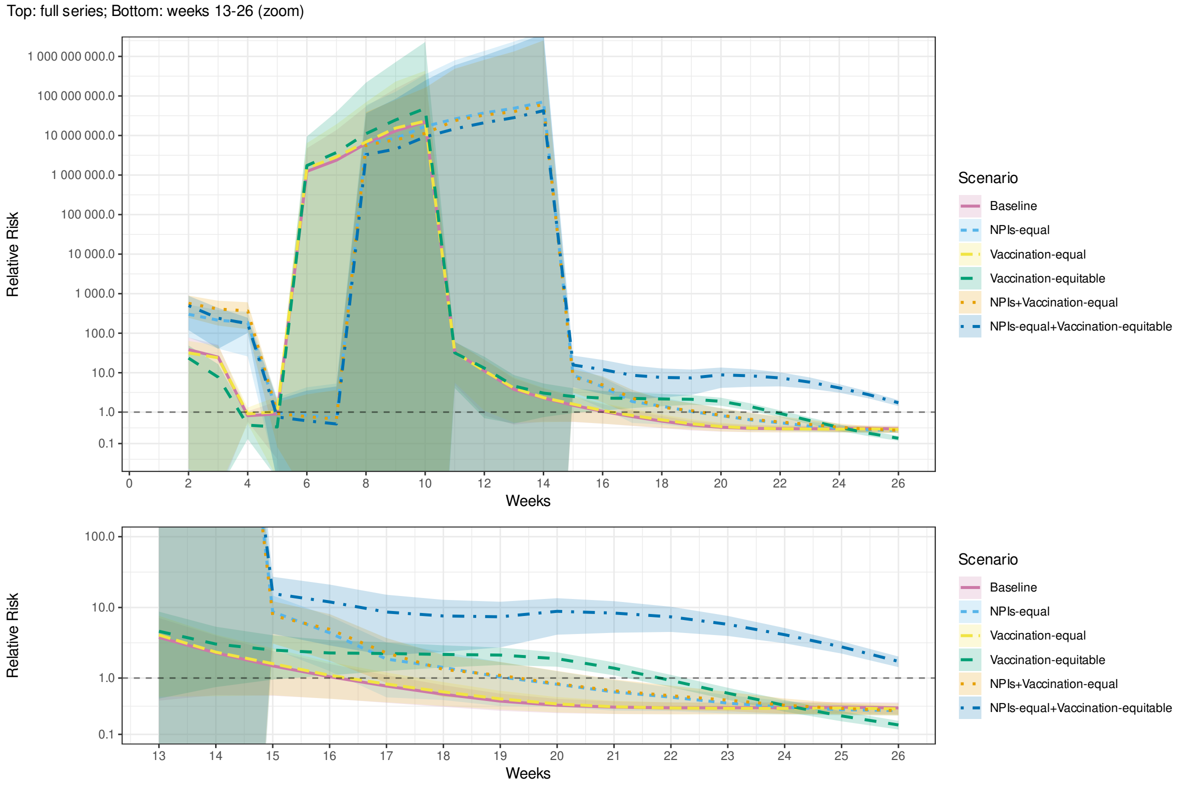


Fig. A.68. Relative risk of mortality in the least vulnerable (Q5) compared to the most vulnerable (Q1) municipalities by SVI during the first 26 weeks of an outbreak in simulations of scenarios 1 and 5–9 (Table 3). Additional regression controls: Bolsonaro vote share and region fixed-effects. Shaded bands indicate 95% confidence intervals.

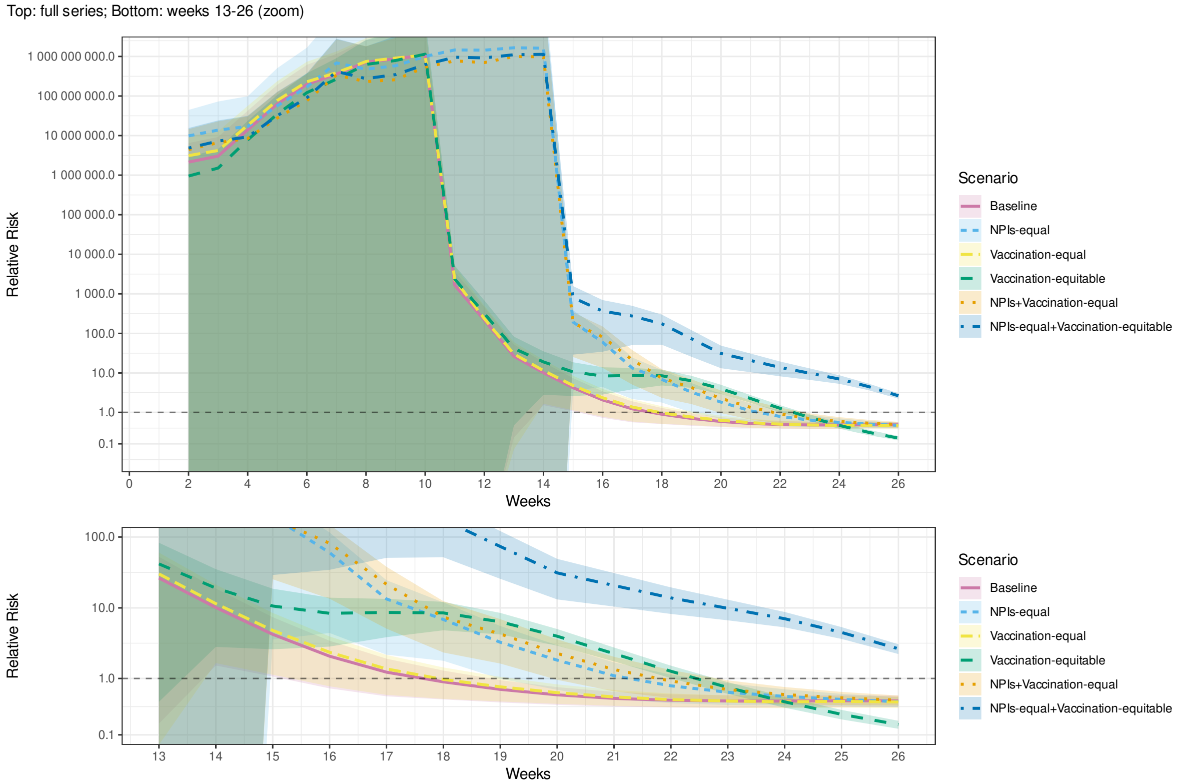


Fig. A.69. Relative risk of mortality in the least vulnerable (Q5) compared to the most vulnerable (Q1) municipalities by MHDI during the first 26 weeks of an outbreak in simulations of scenarios 1 and 5–9 (Table 3). Additional regression controls: Bolsonaro vote share and region fixed-effects. Shaded bands indicate 95% confidence intervals.

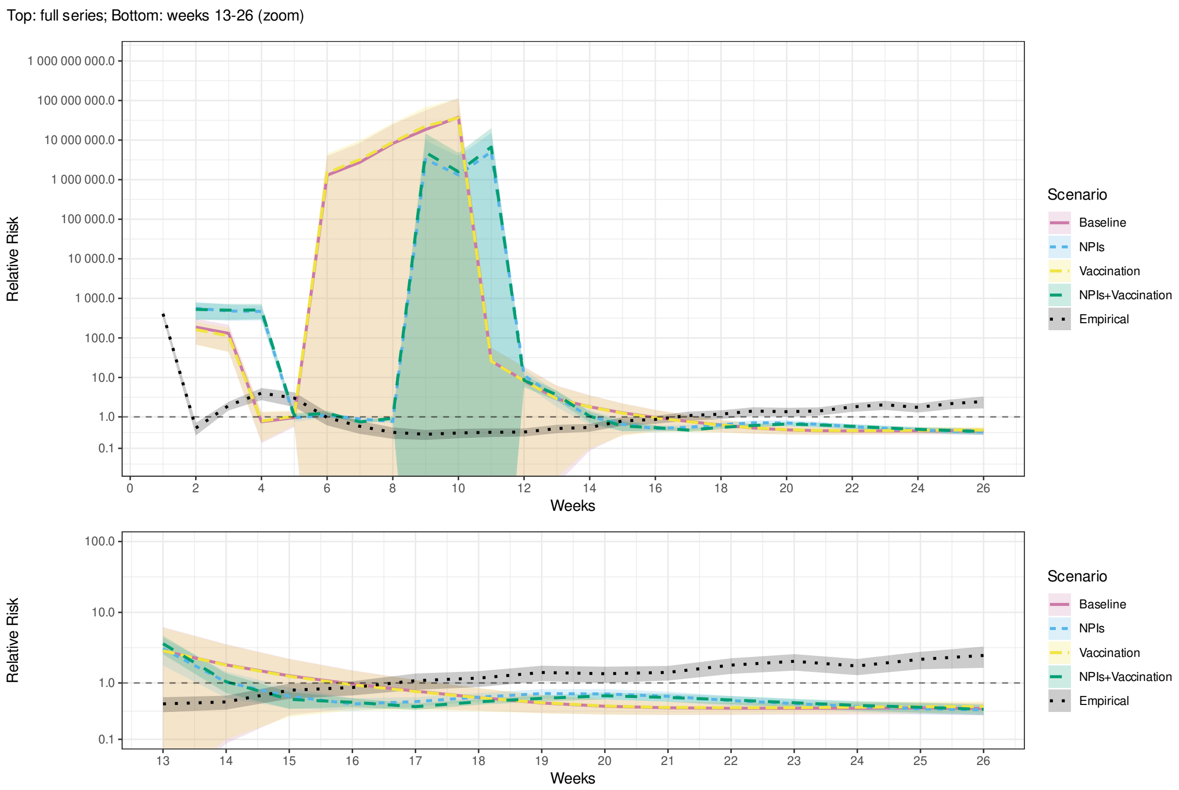


Fig. A.70. Relative risk of mortality in the least vulnerable (Q5) compared to the most vulnerable (Q1) municipalities by SVI during the first 26 weeks of an outbreak in the real world (*Empirical*) and simulations of scenarios 1–4 (Table 3). Additional regression controls: Bolsonaro vote share and state fixed-effects. Shaded bands indicate 95% confidence intervals.

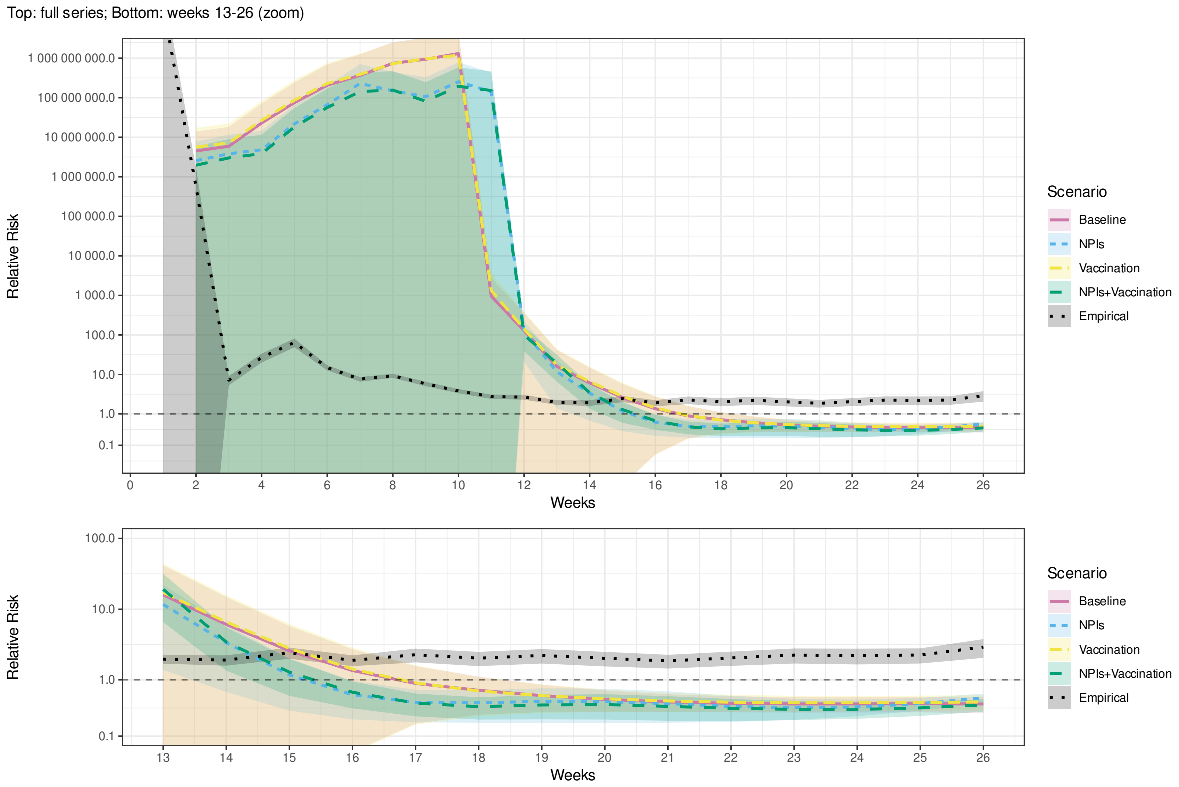


Fig. A.71. Relative risk of mortality in the least vulnerable (Q5) compared to the most vulnerable (Q1) municipalities by MHDI during the first 26 weeks of an outbreak in the real world (*Empirical*) and simulations of scenarios 1–4 (Table 3). Additional regression controls: Bolsonaro vote share and state fixed-effects. Shaded bands indicate 95% confidence intervals.

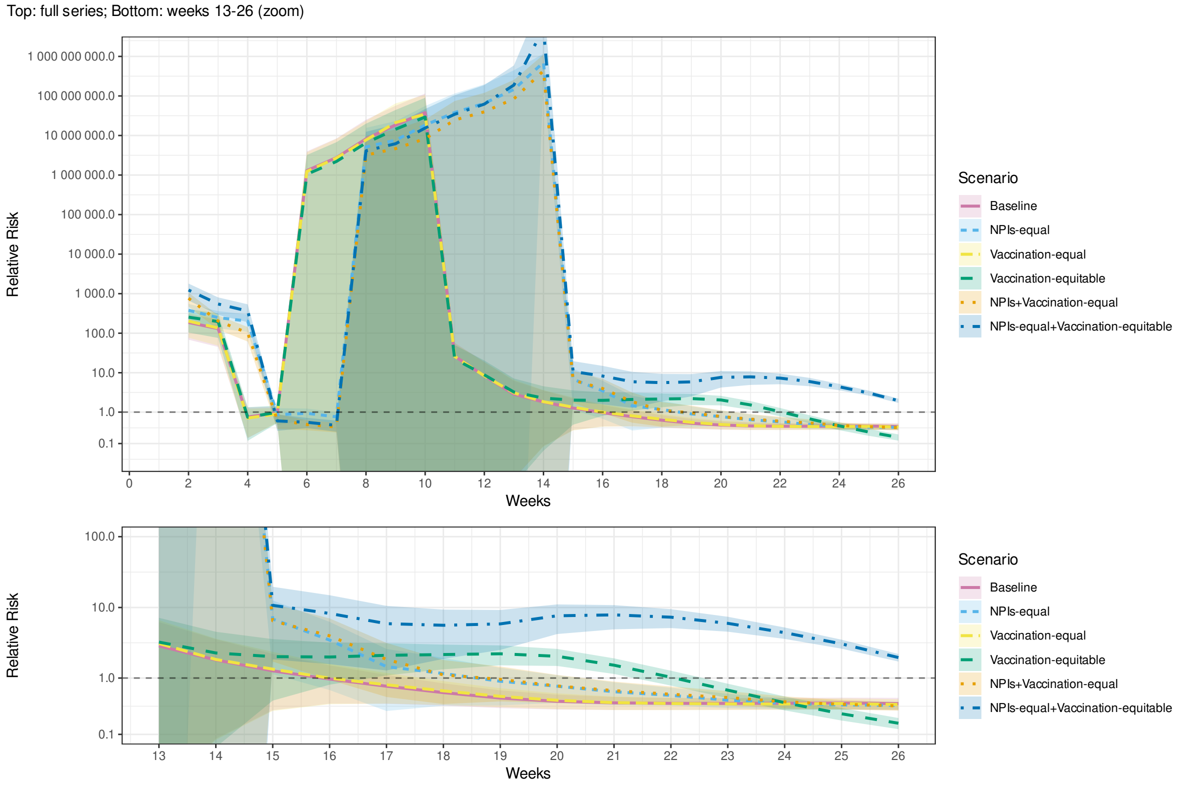


Fig. A.72. Relative risk of mortality in the least vulnerable (Q5) compared to the most vulnerable (Q1) municipalities by SVI during the first 26 weeks of an outbreak in simulations of scenarios 1 and 5–9 (Table 3). Additional regression controls: Bolsonaro vote share and state fixed-effects. Shaded bands indicate 95% confidence intervals.

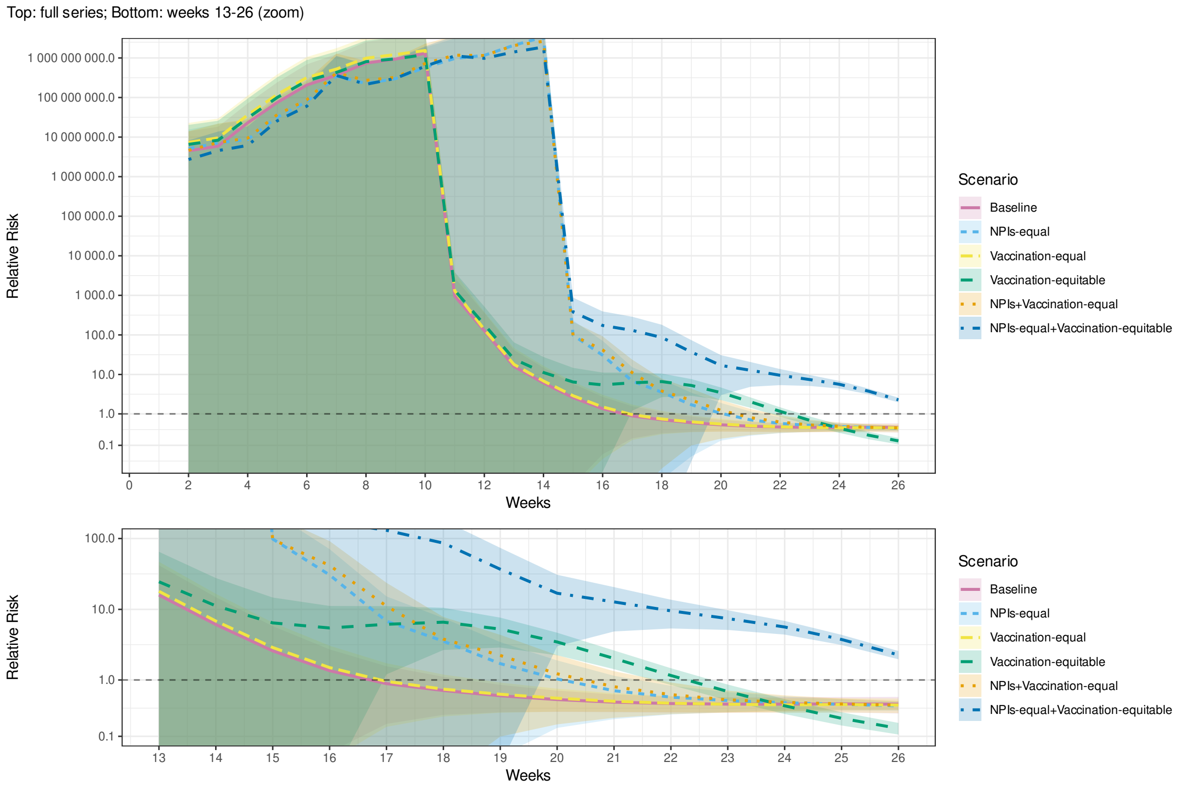


Fig. A.73. Relative risk of mortality in the least vulnerable (Q5) compared to the most vulnerable (Q1) municipalities by MHDI during the first 26 weeks of an outbreak in simulations of scenarios 1 and 5–9 (Table 3). Additional regression controls: Bolsonaro vote share and state fixed-effects. Shaded bands indicate 95% confidence intervals.
